# Genetic prediction and adverse selection

**DOI:** 10.1101/2025.01.20.25320832

**Authors:** Eduardo Azevedo, Jonathan Beauchamp, Richard Karlsson Linnér

## Abstract

The predictive power of genetic data has increased dramatically and is reaching levels of clinical utility for many diseases. Meanwhile, many countries have banned insurers from utilizing genetic information, despite concerns about adverse selection. We make three contributions. First, we develop a method to measure the amount of selection in an insurance market where consumers have access to the current genetic prediction technology. Second, we extend the method to measure selection under expected future prediction technology. Third, using the rich UK Biobank dataset with nearly 500,000 genotyped individuals linked to electronic health records, we apply the method to the critical illness insurance market. We find potentially crippling selection under expected future technology. A robustness analysis using a calibrated equilibrium model of adverse selection yields similar results, with the equilibrium quantity unraveling to zero under future genetic prediction technology.

## 1 Introduction

Technological advancements have led to substantial increases in the predictive power of genetic data (Abdellaoui et al., 2023). Scientists can now create **“polygenic indexes”** (**PGIs**) from a person’s genetic sequence that partially predict the risk of developing various common diseases, such as breast cancer or heart disease. Because common diseases are polygenic, state-of-the-art PGIs sum up the effects of thousands of genetic variants, most of which have a small individual effect (Visscher et al., 2021). Because PGIs work best for common diseases, they are complementary to the long-established use of genetic testing to diagnose rare monogenic disorders.

The predictive power of PGIs has increased dramatically over the last ten years and it recently reached levels of clinical and personal utility (Khera et al., 2018). The first clinical trials are now underway to evaluate whether PGIs can improve routine healthcare or disease screening programs (Klarin and Natarajan, 2022; Saya et al., 2022). Thanks to the continued growth of massive biobanks, experts predict that the predictive power of PGIs will continue to improve and to approach its theoretical upper limits in the coming decades (Abdellaoui et al., 2023). As a result, large healthcare providers, such as the UK’s National Health Service (NHS), are now initiating trials to inform millions of patients about their PGIs (Our Future Health, 2022).

Although PGIs are not yet fully implemented in healthcare, consumers can already purchase prototypes in the consumer genetics market. Tens of millions of people world-wide have already purchased an at-home genetic test, mainly to explore their genealogy (Regalado, 2019). The major consumer genetics companies, such as 23andMe or MyHeritage, have started to bundle their genealogy services with PGI-based disease risk assessments (23andMe, 2023; MyHeritage, 2021). The price of these assessments is declining fast, and they now often cost less than $150—similar to at-home blood or cholesterol tests. Because of these and similar developments, it is likely that both patients and ordinary consumers will soon be routinely informed of their PGI-based disease risk predictions (Zeggini et al., 2019; Meyer et al., 2024).

Meanwhile, in response to concerns about genetic discrimination, many jurisdictions have banned the use of genetic information by insurance companies (Golinghorst et al., 2022).^1^ Such bans typically prohibit insurers from requesting genetic information known to applicants (e.g., from a consumer genetic test) or from utilizing any genetic information at all—creating a distinct information asymmetry. Scholars, policymakers, and insurance industry stakeholders have long voiced concerns that these developments could lead to high levels of adverse selection (Joly et al., 2014; Rechfeld et al., 2019; Maxwell et al., 2021). High levels of selection could then lead to spiraling premiums and to the unraveling of market segments, thus leaving swathes of consumers—including the most vulnerable ones—uninsured (Akerlof, 1970; Einav, Finkelstein, and Fisman, 2023). However, despite decades of research on the potential impact of genetic testing on insurance markets, there is still considerable debate over whether PGIs could eventually create much selection (Golinghorst et al., 2022; Yanes et al., 2024; Dixon et al., 2024).

This study combines large-scale genetic data, economic theory, and econometrics to inform the debate on genetics and adverse selection in insurance markets. We focus on the impact of PGIs on the market for **critical illness insurance** (**CII**), which is estimated to be worth more than $100 billion globally and is growing rapidly (Allied Market Research, 2022). The standard CII contract pays out a lump sum in the event that the insured person is diagnosed with any of a number of medical conditions listed on the policy.^2^

This study makes three key contributions. First, we develop a method to measure the opportunity for adverse selection in an insurance market where consumers have adopted the current genetic prediction technology. The method uses large-scale data on genetics, non-genetic risk factors, and insured events. Second, we extend the method to estimate the future amount of selection given the expected improvements in the predictive power of PGIs, which we derive by combining the estimates from heritability studies with the current PGIs’ correlations with standard non-genetic covariates used in underwriting.

Third, we apply the method to measure selection with both current and future PGIs. We use data from the UK Biobank (UKB) (Bycroft et al., 2018), a large dataset of 500,000 people with genotypic and rich health-related data, including linked data from the UK’s National Health Service (NHS). To illustrate our method in a simple setting, we first study seven single-disease CII contracts—for Alzheimer’s disease, breast cancer, coronary artery disease, colorectal cancer, prostate cancer, schizophrenia, and type 2 diabetes—that each pay a lump sum upon the onset of the disease. We next apply our method to a multiple-disease contract that bundles these diseases.

We report three main findings. First, with current genetic prediction technology, but assuming that all consumers observe their genetic predictions, we find noticeable levels of selection. Current PGIs are meaningfully predictive, and this is reflected in moderate to high levels of selection. For most contracts, the level of selection is non-negligible, but lower than in previously studied market segments that have unraveled. The present situation of low takeup is consistent with the fact that critical illness insurance markets do not yet appear to be plagued by selection from PGIs (Golinghorst et al., 2022).

Second, with the expected future predictive power, the degree of selection becomes extremely high. For all contracts, we find degrees of selection that are within or above the range observed in market segments that had unraveled. Even in a scenario where only 50% of consumers have private knowledge about their genetic predictions, we still find high levels of selection. These results suggest that the increasingly common policy of simply banning insurers from using genetic data may become problematic. Some market segments may collapse, leaving consumers uninsured. In this case, alternative policy options that better promote access to insurance may have to be considered.

Third, we find that there is subtle variation in the amount of selection for different contracts. The amount of selection is mostly determined by the expected accuracy of future prediction technology, by how well observable non-genetic risk factors can predict the diseases, and by the disease prevalence.

We test the robustness of these results in several ways. We reproduce some of the main findings using data from the Health and Retirement Study (HRS). We also calibrate an equilibrium model of adverse selection using the HRS data on risk preferences and industry facts about current CII market conditions. In the calibrated model, availability of the current PGI reduces equilibrium quantity from 30% to 21.4% of potential buyers. Availability of the future PGI reduces the equilibrium quantity to zero, in an adverse selection death spiral. We consider other robustness checks, limitations of our approach, and policy implications.

A key driver of these findings is that we consider genetic predictions based on PGIs, which aggregate information from over one million genetic variants to predict a substantial share of the variation in disease risk. For instance, the current PGI of prostate cancer accounts for 9.9% of the variation and—based on heritability studies—we expect future PGIs to account for 18.0 to 57.0% of the variation. By contrast, popular discussion of genetic prediction often focuses on rare mutations that individually have large effects on the risk of certain diseases, but that explain only a small proportion of the disease cases. Salient examples include mutations in the *BRCA* genes for breast cancer and in the *PKD* genes for polycystic kidney disease. Interestingly, these mutations are unlikely to create problematic selection because they are rare and can be predicted from family history.

Our findings complement a literature that finds that people respond to new genetic information when they make decisions about insurance. Oster et al. (2010) find that carriers of the genetic variant that causes Huntington’s disease (a rare single-gene disorder affecting less than 0.01% of the population) were 5 times more likely to own long-term care insurance compared to the general population. Taylor Jr et al. (2010) find that carriers of the *APOE4* variant, which increases the risk of Alzheimer’s by a factor of 2 to 3, were 2.3 times more likely to buy (or plan to buy) long-term care insurance relative to carriers of the more common *APOE3* variant.

Existing research on the potential impact of genetic testing on insurance markets has focused almost exclusively on tests for rare mutations and has obtained mixed results. Angus S. Macdonald and McIvor (2009) and A. Macdonald (2011) find little impact, while Howard (2016) shows that the impact could be substantial. Our study is the first (to our knowledge) to address this question with large-scale genetic and health data together with an econometric model on multiple-disease contracts that accounts for the predictive power of future PGIs over and above that of observable risk factors.

### 1.1 Illustrative example: CII for prostate cancer

It is possible to understand the gist of our contribution with a simple example. Consider our single-disease contract for prostate cancer. This one-time contract pays $100,000 if the buyer ever incurs prostate cancer by age 65. This contract is a simplification of real CII contracts but, as we will discuss later, it captures several key features. The contract is purchased at age 35, many years before the usual age of onset of prostate cancer. The insurer’s cost of selling this contract is roughly proportional to the buyer’s risk of prostate cancer by age 65. Our methodology allows us to calculate the distribution of risk in a population conditional on various subsets of information. We will focus on information subsets that comprise observable covariates used by insurers in underwriting, as well as on combinations of these covariates with a current or future PGI.

The left panel of Figure 1 shows results based on UKB data. Scenario 1 shows the distribution of the risk of prostate cancer by age 65 in an all-male population of typical consumers, conditional on non-genetic covariates only. This information is observed by both consumers and insurers and is used by insurers to set prices. There would be a non-trivial amount of selection if insurers were not allowed to use this information. But, in practice, insurers do underwrite and define various consumer risk classes which they price differently. Following textbook practice in the insurance industry, we define the **standard risk class** as the set of consumers whose risk of contracting prostate cancer is between 0.75 and 1.25 times the average population risk 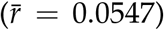 (Brackenridge, Croxson, and Mackenzie, 2006). These consumers are shown in blue in the figure and face the same price of insurance. By definition, these consumers have almost the same risk conditional on the non-genetic covariates, and so there is almost no selection within the standard risk class in Scenario 1.

**Figure 1:**
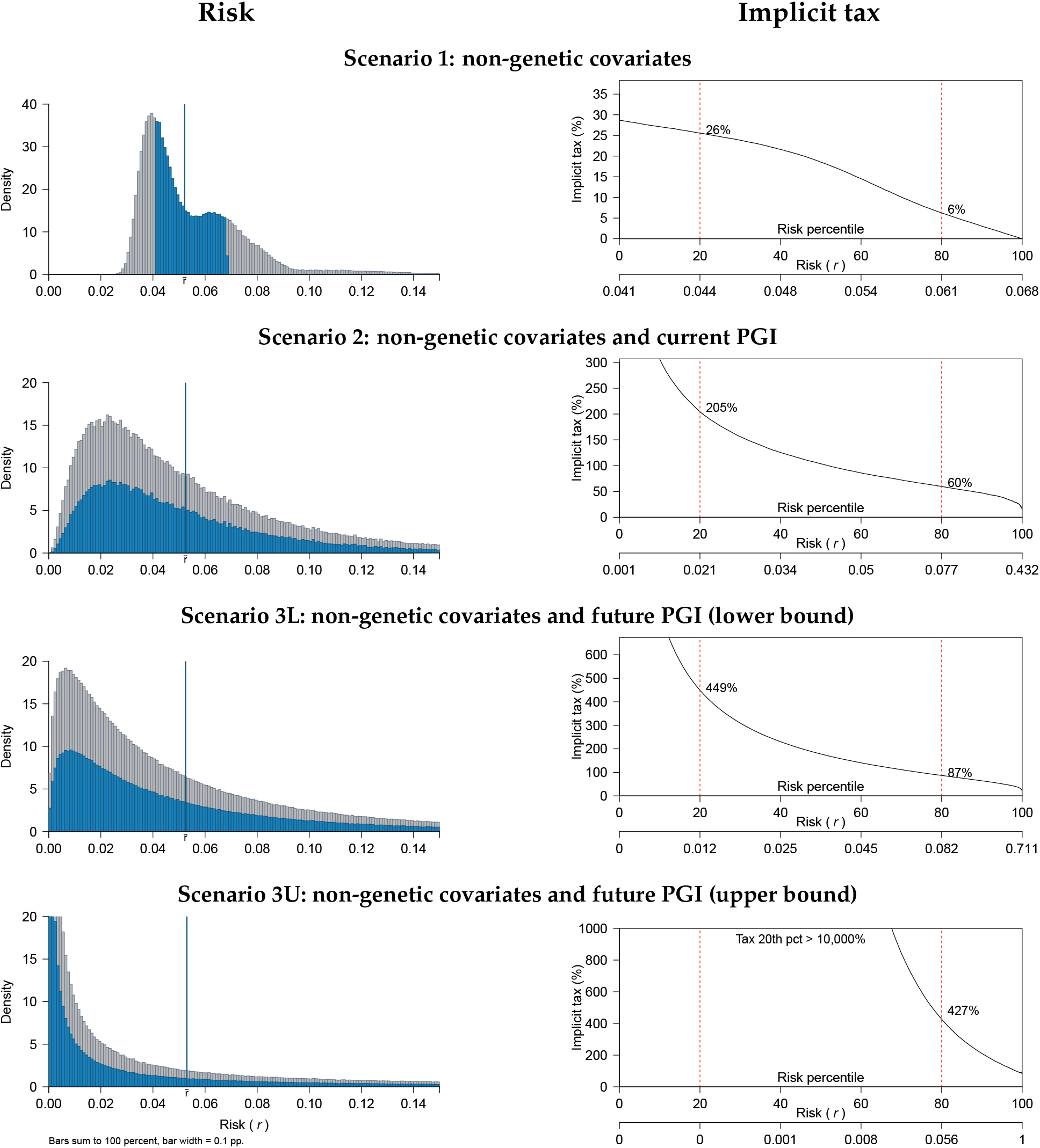
Single-disease CII contract for prostate: risk and implicit tax. *Notes*: Left panel: distribution of the risk of contracting prostate cancer by age 65, conditional on various information scenarios and computed in the UKB data (*N* = 181, 902). The vertical blue line marks the average risk in the standard risk class 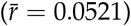. The standard risk class individuals are shown in blue; insurers treat these individuals identically, so the blue distribution corresponds to the distribution of private risk for these individuals. Right panel: implicit tax for consumers in the standard risk class as a function of their percentile private risk of prostate cancer for each scenario.

Scenario 2 shows the distribution of risk conditional on both the non-genetic covariates and the current PGI of prostate cancer. The risk distribution for the standard risk class now has a wider spread. Because insurers are not allowed to observe genetic information, these consumers look like identical risks to insurers and are therefore charged the same price. However, because consumers can observe their PGIs, their privately known risk varies widely.

Scenarios 3L and 3U show the distribution of risk with two future PGIs with different assumed accuracies. These assumed accuracies are informed by existing estimates of the heritability of prostate cancer and correspond to what we consider reasonable lower (Scenario 3L) and upper (Scenario 3U) bounds. In Scenario 3L, the standard risk class now has an even wider spread, with consumers at the 5th and 95th percentiles facing risks of 0.4% and 16.5% of contracting prostate cancer, respectively. And in Scenario 3U, consumers at the 5th percentile face a negligible risk (as can be seen from the spike at 0) vs. a 31.4% risk for consumers at the 95th percentile. Thus, consumers who face the same price of insurance vary widely with respect to their privately known risk. This suggests that adverse selection could be a serious issue.

To quantify the degree of asymmetric information and adverse selection that would result from this situation, we employ the **implicit tax**, a metric proposed by Hendren (2013) and Hendren (2017). The right panel of Figure 1 shows for each scenario the estimated implicit tax as a function of a given consumer’s risk percentile for prostate cancer, for the consumers in the standard risk class. In Scenarios 2, 3L, and 3U, the implicit taxes imply elevated levels of selection that exceed those that have been estimated for market segments that have unraveled. This, along with the results we present below for the other CII contracts we study, suggests that selection due to future genetic prediction technology will be a first-order concern and may make some market segments non-viable.

The paper is structured as follows. Section 2 provides background on genetics and the CII market. Section 3 describes the theory. Section 4 describes the data and empirical specification. Section 5 presents the main results. Section 6 discusses limitations, robustness, and extensions. Section 7 concludes.

## 2 Background

### 2.1 Genetics background

Some conditions, such as Huntington’s disease, are caused by single genetic variants. Such conditions are called monogenic. Some traits, like eye color, are caused by variation in a relatively small number of genes (Simcoe et al., 2021). However, the vast majority of traits are polygenic: they are influenced by a very large number of causal genetic variants (as well as by environmental factors), with each causal genetic variant having a tiny effect (Visscher et al., 2021). For example, height, BMI, and cognitive ability are quintessential polygenic traits. The seven diseases we study here (e.g., Alzheimer’s disease, or coronary artery disease) are all good examples of polygenic diseases.^3^

Consider first how genetic data are used to predict a continuous polygenic trait *Y*, such as height. The entire genome contains more than 3 billion base pairs (labeled with the letters “A”, “C”, “G”, and “T”). But most of the variation across human individuals is concentrated on <5% of base pairs (Walsh and Lynch, 2018). Thus, typical genotyping datasets contain data on millions rather than billions of variants.^4^

*γ* is assumed to follow the **additive genetic model** (Falconer and Mackay, 1996): genetic effects are linear with no interactions. Accordingly, *Y* is the weighted sum of an individual’s genetic variants plus a disturbance term *ψ*:

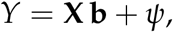

where **X** is a row vector that contains the measured variants, and **b** is a column vector that contains their true effect sizes on *Y. G*^*\**^ = **X b** is the individual’s **true additive genetic factor** for *Y*. (Throughout, we denote random variables with uppercase and their realizations and constants with lowercase letters, and vectors with bold font.) Despite its simplicity, the additive genetic model is commonly used in genetics, and both theory and evidence suggest that additive effects account for the bulk of the genetic variation for most polygenic diseases (Hill, Goddard, and Visscher, 2008).

In practice, neither **b** nor *G*^*\**^ are observed. Instead, one can obtain estimates 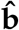 from a previously published **genome-wide association study** (**GWAS**)^5^ of the trait and use them to construct a **polygenic index** (**PGI**) of *Y*:

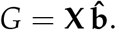

Thus, the observed index *G* is a noisy measure of the true additive genetic factor *G*^*\**^.^6^ Let *ϵ* be the error: *G* = *G*^*\**^ + *ϵ. ϵ* stems from measurement error in 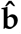, which is due to sampling variation in the GWAS. To avoid overfitting, the GWAS is conducted in a sample that is independent of the analysis dataset in which the PGI is constructed, so *ϵ* and *G*^*\**^ are independent. Supplementary Information 1 provides further background on the genetics of polygenic traits, on GWASs, and on PGIs.

As mentioned, a polygenic trait is influenced by a large number of genetic variants, each with a tiny effect. This implies, by the central limit theorem, that *G*^*\**^ = **X b** is approximately normally distributed. By the same reasoning, so is *ϵ*. As a result, so is *G*—i.e., PGIs also are approximately normally distributed.

This model for a continuous trait can easily be adapted to a **disease** *D*. It is common practice to use a liability threshold model such as probit, which we do in our analysis.

As GWASs have rapidly grown in sample size over the past decade (Tam et al., 2019), estimates of **b** have become much more precise; as a result, the predictive power of PGIs has also increased dramatically (Visscher et al., 2021). As GWAS sample sizes keep increasing, and as the set of genetic variants they analyze is augmented to include rarer variants, PGI predictive power should increase further (Daetwyler, Villanueva, and Woolliams, 2008; Chatterjee, Shi, and García-Closas, 2016). In theory, the upper bound for a PGI’s predictive power for a given trait is the trait’s narrow-sense heritability (So and Sham, 2010; Dudbridge, 2013), defined as the share of the trait variation that is attributable to additive genetic factors.

Table 1 shows our in-sample estimates of the current PGI *R*^2^’s for the seven diseases we study, together with previous studies’ estimates of their **SNP heritabilities** and of their **twin heritabilities**. Since the diseases are binary, we report their *R*^2^ **on the liability scale**, also known as the McKelvey & Zavoina pseudo-*R*^2^ (Lee et al., 2012; McKelvey and Zavoina, 1975). A disease’s **SNP heritability** is the share of the variance in its liability that is attributable to the set of common genetic variants used in the SNP heritability estimation.^7^ Because not all variants are used (as most were not measured, and for computational reasons), SNP heritability is necessarily lower than the true heritability. A trait’s **twin heritability** is computed by comparing the resemblance of monozygotic twins—who share 100% of their genomes—to that of dizygotic twins—who share 50% on average. Under some assumptions, it is a consistent estimate of the trait’s true heritability.

**Table 1:** Descriptive statistics for selected diseases.

| Disease | Prevalence | Lifetime risk | Current PGI $R^2$ | SNP $h^2$ | Twin $h^2$ |
| --- | --- | --- | --- | --- | --- |
| Alzheimer’s disease | 7.1% | 6.3% (M), 12% (F) | 7.1% | 33.1% | 58% |
| Breast cancer | 8.1% | 12.3% | 6.7% | 14% | 31% |
| Coronary artery disease | 3.0% | 51.7% (M), 39.2% (F) | 2.5% | 22% | 40% |
| Colorectal cancer | 2.9% | 4.6% | 2.2% | 9% | 40.2% |
| Prostate cancer | 0.9% | 12.7% | 9.9% | 18% | 57% |
| Schizophrenia | 0.46% | 0.7% | 4.9% | 24% | 79% |
| Type 2 diabetes | 3.9% | 27% (M), 18.6% (F) | 7.4% | 19.6% | 72% |

Notes: Current PGI $R^2$ ’s are on the liability scale and were estimated in the UKB. All other statistics come from earlier, separate studies of the diseases and were not produced with a uniform methodology across diseases (for references, see Supplementary Table 1). Prevalence and lifetime risk include various age-corrected measures (e.g., the prevalence of Alzheimer’s disease is reported for ages above 65).

While it is impossible to know with certainty how predictive a disease’s future PGI will be, it is likely that it will explain at least as much as the disease’s SNP heritability estimated with common variants. This is because future PGIs, in addition to being constructed with more precise estimates of common variants’ effect sizes, will also account for the effect sizes of less common variants. And, as mentioned, the theoretical upper limit on a PGI’s predictive power for a disease is the disease’s heritability, which we estimate with the twin heritability. In our analyses, we report results under the assumptions both that future PGIs’ accuracies will be equal to their SNP heritabilities (as lower bounds) and their twin heritabilities (as upper bounds).

We make no attempt to pinpoint the time it would take for PGI predictive power to reach the heritability. There is debate in the literature about the relationship bewteen GWAS sample size and PGI predictive power. There is even greater uncertainty about what methodological improvements will take place, and what GWAS sample sizes will be available in any given year. Thus, our approach is to consider different possiblities for *R*^2^ in the future, such as the SNP heritability and the twin heritability, and to analyze the implications of each of these possibilities.

As is currently standard practice in applied genetic research, our analysis only uses data from respondents of European-like ancestry, for two main reasons. First, available genetic data for other ancestries are still scarce (Mills and Rahal, 2020); as a result, most large-scale GWASs whose estimates we can use to construct our PGIs were conducted among European ancestry individuals. Second, PGIs based on European ancestry GWAS estimates tend to perform poorly among other ancestries (Martin et al., 2017).

### 2.2 Critical illness insurance

**Critical illness insurance** (**CII**) pays out a lump sum in the event that the insured person gets diagnosed with any of the medical conditions listed on the policy. The lump sum can be used as the policyholder wishes. The policy pays out once and is thereafter terminated. The typical policy is renewable term at guaranteed rates, which are determined at the outset by medical underwriting (Brackenridge, Croxson, and Mackenzie, 2006).

Typical contracts cover some 20 to 50 core illnesses or medical conditions, including different types of cancers (e.g., breast, prostate, colorectal, and lung cancer), coronary artery disease, multiple sclerosis, stroke, as well as serious conditions like paralysis, blindness, and unspecified terminal illness (Gatzert and Maegebier, 2015). Enhanced plans exist that cover additional conditions, such as dementia, diabetes, or severe psychiatric illness. Though CII contracts typically cover a menu of conditions, cancers and cardiovascular diseases (including in particular heart attacks, heart surgery, and strokes) account for over 80% of all CII claims, with cancers accounting for the bulk of these.^8^

Major CII markets include Canada, the United Kingdom, Japan, Australia, India, China, and Germany. It is estimated that 20% of British workers were covered by a CII policy in 2009 (Gatzert and Maegebier, 2015). The global CII market has been valued at over $100 billion in 2021 and was projected to grow to over $350 billion by 2031 (Allied Market Research, 2022).

Contract details vary across markets. One dimension of variation is the set of covered illnesses, which has tended to increase over time and is a point of differentiation between competing firms’ offerings. Most contracts cover a suite of diseases, though some—such as cancer insurance in the US and Japan (Gatzert and Maegebier, 2015)—only cover one disease or a narrow group of diseases. Another dimension of variation is whether policies include other benefits, such as paying back a fraction of the premium in case of death of the policyholder. Some contracts are standalone, whereas others are riders to another insurance policy such as life insurance; some contracts are purchased via one’s employer while others are purchased directly from the insurer.

Underwriting for individual policies is similar to life insurance underwriting. For individual policies, insurers request detailed information, and may ask for medical records, blood or other medical tests, and medical examinations (Canadian Institute of Actuaries, 2012). Industry reports often discuss concerns about adverse selection, and some contract features like waiting periods are used as a safeguard (Gatzert and Maegebier, 2015).

CII and genetics have been studied in the actuarial literature (Angus S Macdonald, Waters, and Wekwete, 2005; Angus S. Macdonald and McIvor, 2009; A. Macdonald and Tapadar, 2010; A. Macdonald and Yu, 2011; Adams, Donnelly, and A. Macdonald, 2015). They investigate the impact genetic information will have on CII and generally find that adverse selection due to private genetic information is unlikely to be a problem. However, most of these studies are from more than a decade ago and, consistent with the genetic prediction technology at the time, mainly consider single-gene mutations that account for only small shares of the population variation in the studied disorders.

More recently, Maxwell et al. (2021) found that current PGIs predict breast cancer and coronary artery disease above and beyond normal underwriting factors, and discuss the possibility of adverse selection. In a study for the Canadian Institute of Actuaries, Howard (2016) considers rare mutations associated with six diseases and finds that banning insurers from using genetic data would have a marked impact on CII claims rates.

## 3 Theory

We now introduce our economic and econometric models. We focus on single-disease contracts to facilitate exposition. Section 5.3 and Supplementary Information 3 generalize these models to multiple-disease contracts.

### 3.1 Economic model

#### The model

Consider a population of consumers who may incur a binary loss *L* and can purchase an insurance contract for it. In a single-disease CII contract, the loss corresponds to contracting the disease: *L* = *D*.

Throughout this section, we consider a population that is treated equally by insurers, such as consumers in the same risk class. We assume that consumers have additional private information, so that there may be selection. This additional private information can include both genetic and non-genetic information.

Each consumer is characterized by the realization of four random variables (*L, G*_*c*_, *G*_*f*_, **W**). The **loss** (or **disease**) *L* is a random variable equal to zero or one depending on whether a consumer contracts a disease. *G*_*c*_, the **current polygenic index** (**PGI**) of the disease, is a sufficient statistic of the consumer’s DNA for the purpose of estimating the probability of contracting the disease, conditional on the current genetic prediction technology. *G*_*f*_, the future PGI of *L*, is a sufficient statistic of the consumer’s DNA conditional on the future genetic prediction technology. The **non-genetic covariates W** are a row vector of covariates. The variables (*L, G*_*c*_, *G*_*f*_, **W**) are independently identically distributed across consumers, with joint distribution ℙ. *G*_*c*_ and *G*_*f*_ take real values. **W** takes values in Euclidean space.

#### Private information about risk

Each consumer has private information about her non-genetic factors **W** and her PGI(s) *G*. We will consider four counterfactual scenarios with different sets of consumer private information that vary as a function of the assumed genetic prediction technology: with no genetic prediction technology (Scenario 1); with the current PGI (*G* = *G*_*c*_; Scenario 2); and with the future PGI (*G* = *G*_*f*_) with lower-bound (Scenario 3L) and upper-bound (Scenario 3U) predictive power.

Denote consumers’ private information about risk as follows. The **non-genetic private risk function** is defined as the probability *π*(**w**) of the loss conditional on non-genetic factors **W** = **w**:

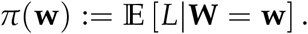

The **private risk function** is defined as the probability *ρ*(*g*, **w**) of the loss conditional on the PGI(s) observed by the consumer, *G* = *g*, and non-genetic factors **W** = **w**:

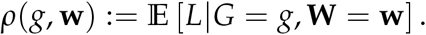

Define the random variable for **non-genetic private risk** as *P* := *π*(**W**), and the random variable for **private risk** as *R* := *ρ*(*G*, **W**). The realizations of losses as well as both non-genetic private risk and private risk are independent across consumers.

#### Measures of adverse selection

The distribution of private risk *R* across the population is informative about the degree of adverse selection when consumers have genetic information. The left panel of Figure 1 displays the distribution of the risk of contracting prostate cancer by age 65 under the different information scenarios. The blue region represents consumers in the standard risk class. In Scenario 1, where consumers only have non-genetic private risk, the distribution is concentrated around the mean. Therefore, consumers have little private information about their own risk, and consumers who purchase a single-disease CII contract for prostate cancer are not likely to be particularly high-risk, making it easier for an insurance market to perform well. By contrast, the distributions in Scenarios 3L and 3U, where consumers also observe future PGIs, are highly dispersed. Therefore, some consumers know that they have low risk and other consumers know that they have high risk. So consumers who purchase more insurance are likely to be those with particularly high risk. This makes it more likely that an insurance market will perform poorly. In standard adverse selection models, few or no consumers may end up buying insurance, because firms expect that only high-risk consumers would want to purchase and so set a high price, which further deters low-risk consumers, which leads firms to set an even higher price (Akerlof, 1970).

Adverse selection depends on the entire distribution of private risk *R*. Therefore, for the contracts we study, we will consider the key distributions with histograms like those in Figure 1. Moreover, we will also report an economic measure of selection, the **implicit tax** (Hendren, 2013). The implicit tax *t*(*r*) is defined as the extra amount a consumer with private risk *r* would have to pay if she had to buy insurance at the actuarially fair price for all consumers with private risk *r* or greater, versus what her own actuarially fair price is. The formal definition is in Appendix A. Intuitively, the implicit tax tells us how much more the marginal consumer has to pay to buy insurance due to adverse selection, and gives this number as a percentage tax that is easy to interpret.

The implicit tax measure has three advantages. First, it is intuitive. Second, Hendren (2013) proposed a no-trade theorem that shows that sufficiently high values of the minimum of the implicit tax cause insurance markets to break down under certain assumptions. Third, we can compare the level of selection we find with earlier studies. Hendren (2013) estimated the implicit tax in market segments that had unraveled (in the sense of not being served by insurers) and in market segments that had not unraveled. He reports the minimum implicit tax up to the 80th percentile of risk, which we denote *t*_80_. For market segments that had not unraveled, Hendren estimated *t*_80_ to be between 7% and 35%, and for unraveled segments, he estimated *t*_80_ to be between 43% and 83%.

### 3.2 Econometric model and identification

Our model is fully specified by ℙ, the joint distribution of (*L, G*_*c*_, *G*_*f*_, **W**).^9^ While *G*_*c*_ can be observed in the UKB data by the econometrician, *G*_*f*_ cannot. Define the distribution of the observed data as a joint distribution ℙdata over (*L, G*_*c*_, **W**). We say that the model is identified if there is a unique value of ℙ consistent with ℙdata.

It may at first appear that the model cannot be identified. Identification implies that we can use the data to estimate the degree of selection in a world in which consumers can observe their future PGIs. It turns out that the model is identified under reasonable assumptions. The key idea is that empirical regularities from quantitative genetics considerably restrict the data, making identification possible. Our assumptions are based on two empirical regularities. First, PGIs are approximately normally distributed. And second, existing heritability estimates for a disease are informative about the likely predictive power of its future PGI.

We now state the necessary assumptions. The first assumption is a regularity condition on the distribution of covariates.

#### Assumption 1.

*[Covariates’ distribution]* **W** *is distributed according to a distribution* ℙ_**W**_ *in Euclidean space. The support of* **W** *spans the entire space*.

The next two assumptions are conditional normality assumptions on *G*_*c*_ and *G*_*f*_. They are motivated by the two regularities mentioned above and by the fact that what distinguishes a current PGI from a future PGI for a disease is simply the size of the independent and approximately normally distributed error. Further, we take *G*_*c*_ to be standardized.

#### Assumption 2.

*[Gaussian future PGI] Conditional on observables, the future PGI has a Gaussian distribution. The variance is constant and the mean is a linear function of observables:*

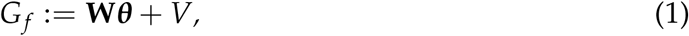

*with* 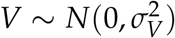 *and V ⊥* **W**.

#### Assumption 3.

*[Noisy current PGI] The observed current PGI G*_*c*_ *has unit variance*, Var[*G*_*c*_] = 1, *and is a noisy estimate of G*_*f*_ :

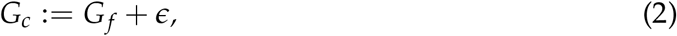

*with* 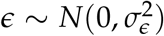 *and ϵ ⊥* (**W**, *V*).

The fourth assumption is that *L* follows a probit model, which is the standard functional form for binary diseases in genetic epidemiology. The assumption also implies that the current PGI has no predictive power beyond the future PGI.

#### Assumption 4.

*[Probit disease model] Disease probabilities follow a linear probit model:*

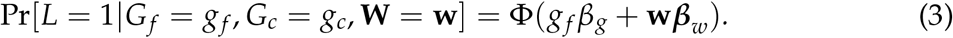

Under assumptions 1–4, the distribution ℙ has parameters

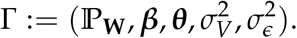

For the model to be identified, we need one more assumption. We assume that we know the predictive power of the disease’s future PGI *G*_*f*_, which we denote 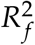, from heritability studies.

#### Assumption 5.

*[Future PGI predictive power] The pseudo R*^2^ *on the liability scale of a probit regression of L on G*_*f*_ *equals a known constant* 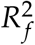.

We are now ready to state the identification result.

#### Theorem 1.

*Under assumptions 1–5, the single-disease model is identified*.

Appendix B provides the proof. The intuition is that the heritability estimates tell us how predictive the future PGI is on its own. The correlations in the current data tell us how much added value over the covariates the future PGI will bring. And the normality assumptions take us from correlations to identifying the full distribution.

The formal proof takes a different path from the intuition. The first key obsevation is that the posterior distribution of *G*_*f*_ given the data is normal and given by a standard formula from Bayesian statistics. The second key observation is that this implies that the disease model is also probit in *G*_*c*_. And that the model parameters are functions of feasible regressions, that use *G*_*c*_ instead of *G*_*f*_. This observations yield not only the identification proof, but also a statistically, computationally efficient estimator, feasible for large data.

## 4 Data and empirical specification

### 4.1 Data

We analyze data from the UK Biobank (UKB) (Bycroft et al., 2018; Sudlow et al., 2015). The UKB has collected genotypic and rich health-related data on about 500,000 people from across the United Kingdom. The participants were between 40 and 69 years at the time of recruitment in 2006–2010. The data is linked to the UK’s National Health Service (NHS), which maintains electronic health records (EHR) and with which 98% of the UK population is registered (Sudlow et al., 2015). Furthermore, the participants completed a baseline assessment, in which they provided extensive questionnaire and interview data on environmental, family history, health, lifestyle, physical, and sociodemographic factors, as well as biomedical assays from blood, saliva, and urine samples. Table 2 reports sample descriptive statistics for the seven diseases and multiple-disease contracts (Panel A) and a set of disease-general factors (Panel B; these are discussed below).

**Table 2:**
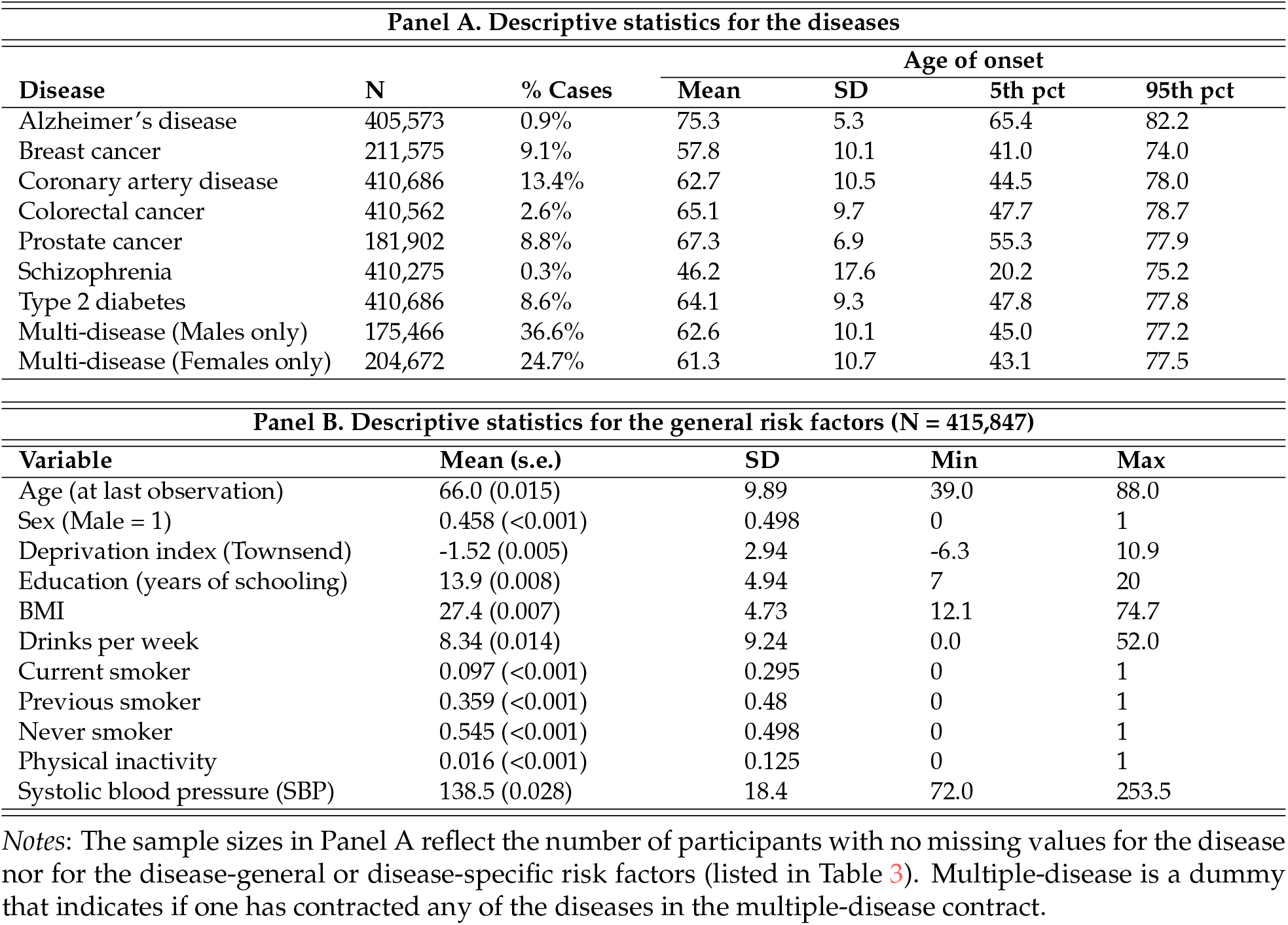
Sample descriptive statistics.

The UKB holds genotypic array data on *∼*488,000 participants. Our study sample was restricted to 446,570 participants of European-like ancestry and whose genotypic data passed a number of quality control filters. We use these genetic data and GWAS estimates for each disease to compute each disease’s current PGI. The GWAS estimates come from the largest available GWAS on the disease whose analysis sample did not include the UKB.^10^ Supplementary Information 1 provides additional details on the PGIs.

To code the diseases for statistical analysis from the unstructured EHR-data, we relied on the widely used “Phecode” mapping system (Denny et al., 2013). The Phecode system was designed specifically to handle the complexities of defining clinically meaningful case-control status from various complex EHR sources (Bastarache, 2021). The mapping uses the categories of the International Classification of Diseases (ICD9/10) as reference point. A central feature is to condense about 90,000 highly detailed ICD subcodes (e.g., C50.211: “malignant neoplasm of upper-inner quadrant of female breast”) into about 1,900 major case-control status codes (174.1: “Breast cancer [female]”).

We reviewed clinical guidelines and the epidemiological literature to identify the main disease-specific non-genetic risk factors that are used either by medical practitioners or by insurers to assess individual disease risk. It is important for our analysis to model the medical information that is typically observed by insurers during underwriting. These disease-specific risk factors are listed in Table 3, together with a set of disease-general factors. About half of the risk factors were sourced from the assessment centre data, and the other half from the linked EHR-data (e.g., ICD-codes for hypertension). These factors are the model covariates in **W**, together with a genotyping array dummy and the top ten genetic principal components (PCs). Adjusting for genetic PCs is best practice in applied genetic research (Price et al., 2006). Supplementary Information 2 provides additional details on the coding of the health data.

**Table 3:** Non-genetic risk factors for the studied diseases.

| Disease | Key risk factors |
| --- | --- |
| General risk factors | Age, sex, Townsend Deprivation index, education (years of schooling), body mass index (BMI), alcohol consumption, smoking (current, ex, or never), physical inactivity, systolic blood pressure, family history* (did each of father, mother, or siblings ever suffer from the disease?) |
| Alzheimer's disease | Air pollution, [emphAPOE risk type], [brain injury], [coronary artery disease], depression, diabetes type 1, [diabetes type 2], hearing loss, hypertension, physical inactivity, social isolation |
| Breast cancer | Diabetes type 1, [diabetes type 2], ever had breast cancer screening / mammogram, ever had hormonal replacement therapy, hormonal/oral contraceptives, age at menarche, age at menopause, age at first birth, number of live births |
| Coronary artery disease | Crohn's disease, diabetes type 1, [diabetes type 2], [diet], ever had bowel cancer screening, irritable bowel syndrome (IBS), polyp of colon, ulcerative colitis (and other non-infective gastro-enteritis and colitis) |
| Colorectal cancer | Depression, diabetes type 1, [diabetes type 2], [diet], hypercholesterolemia, hyperglycemia, hypertension, social isolation |
| Prostate cancer | Ever had prostate specific antigen (PSA) test |
| Schizophrenia | Anxiety, bipolar disorder, [cannabis use], depression, neuroticism, psychopathology**, [substance use] |
| Type 2 diabetes | [Diet], hypercholesterolemia, hyperglycemia, hypertension, social isolation |

Notes: The non-genetic risk factors were identified in a review of clinical guidelines and the epidemiological literature, and the search findings were verified by two medical doctors. References are reported in Supplementary Table 3. All the risk factors listed in the table were coded as variables in the UKB, with the exception those (i) that were not well-defined (e.g., diet), (ii) for which data were lacking (e.g., brain injury, cannabis use), (iii) that were a disease outcome in the multiple-disease contract (e.g., type 2 diabetes), or (iv) that were captured by the PGI (i.e., *APOE*). These risk factors are indicated in square brackets. For more details on the variable coding, see Supplementary Table 4.
\* For schizophrenia, family history of "severe depression" was used as a proxy.
\*\* Psychopathology was proxied by neuroticism score, bipolar disorder, and depression.

### 4.2 Contracts and specification

We consider a loss to be the occurrence of a covered disease. We consider standard contracts that simply pay a lump sum when that loss occurs, thus ignoring the possibility of non-standard contracts. Real-world contracts vary across several dimensions (see Section 2.2), so we narrow down our analysis to particular types of contracts, especially in two key contract dimensions: the set of covered diseases and the coverage period. We also specify a population of interest (i.e., a risk class) and the covariates. With this, we have a full specification to take the observable variables from our model (the loss *L*, the current PGI *G*_*c*_, and the covariates **W**) to the data.

#### Set of covered diseases

CII contracts vary in the number of diseases they cover. Most contracts cover some 20 to 50 illnesses and medical conditions, and the set of diseases has been a point of differentiation between different insurers. Nevertheless, cancers and cardiovascular diseases account for over 80% of all CII claims. Some insurers offer enhanced policies with multiple tiers of covered diseases and medical conditions. There are also less comprehensive contracts such as cancer insurance and single-disease CII contracts.

We consider the seven diseases listed in Table 1. These include three of the four most common cancers in Western countries—prostate, breast, and colorectal cancer—which by themselves account for a substantial share of CII claims.^11^ As a proxy for cardiovascular disease—the next most common source of claims after cancer—we include coronary artery disease.^12^ We include Alzheimer’s disease because it is important for long-term care insurance, so the results might be useful for future research on that market. Schizophrenia is included as an example of a highly heritable but rare condition. We include type 2 diabetes because it is of public health concern and can be included in enhanced CII plans, although it is not covered by standard CII policies.

Many of the diseases and medical conditions covered by real CII contracts but not by our analysis are rare. Some industry observers believe that insurers include rare diseases because consumers value the more comprehensive policies, perhaps due to poor statistical reasoning, even though the costs are negligible from an actuarial perspective.

We first consider single-disease contracts. Section 5.3 extends our methodology to multiple-disease CII contracts that bundle the diseases and pay the lump sum upon occurrence of any of the diseases, and then terminate. Since most CII claims are dominated by a few diseases, and since most of these are included in our seven diseases, our multiple-disease contracts approximate real CII contracts.

#### Coverage period

Typical CII contracts are guaranteed renewable at a predetermined rate up to a maximum age. It is common for the maximum age to be 65. We will restrict attention to this type of contract. The simplest way to model this contract is to assume that all consumers buy contracts at the same age and renew until age 65. Here, we will assume that all consumers buy contracts at age 35, a typical age of first CII purchase that is lower than the usual age of disease onset. Therefore, we will effectively consider one-time 30-year contracts. Ignoring interest rates, this means that insurers’ costs are proportional to the probability that the consumer will incur a covered disease by age 65. This is the approximation we will use. In the terminology of our model, we set the loss *L* = 1 if the consumer ever incurs a covered disease by age 65.

We stress that this is an approximation; it departs from reality in two important ways. First, in practice, consumers do let policies lapse. Lapsation is a secondary but important driver of insurance premiums. For example, in life insurance it has been documented that older consumers are more likely to let their policies lapse, even though they have the greatest probability of dying. This is crucial for firms to make positive profits (Gottlieb and Smetters, 2021). Second, the timing of losses also influences insurers’ costs, due to time discounting.

#### Population of interest

We are interested in a population of 35-year-olds who may purchase CII contracts. Our model considers a population of consumers who are treated similarly by insurers. We follow standard underwriting practices to define this group. Insurers often define the **standard risk class** as all consumers who are within 0.75 to 1.25 times the average population-wide risk, based on underwriting using the covariates insurers can observe. It is typical for insurers to charge the same rates to all consumers in the standard risk class. We take the standard risk class to be the population of interest in the analyses we report in the main text. This fits our model, where we measure selection within a group of consumers who are treated identically by insurers.

#### PGIs and covariates

For each disease, we construct the current PGI *G*_*c*_ based on the genetic data from the UKB and effect-size estimates from prior GWASs, as described in Section 4.1.

#### Scenarios and information sets

For each contract, we produce results under the four Scenarios 1–3U. In all scenarios, insurers effectively only observe a number of discrete consumer risk classes (including the standard class).^13^ The scenarios differ with respect to the consumers’ private information sets. In Scenario 1, consumers observe only their non-genetic covariates and thus observe no more than a moderate amount of non-genetic private risk *P*. In the other scenarios, consumers observe both their non-genetic covariates and their current or a future PGI, and thus observe their private risk *R*. In Scenario 2, consumers observe their current PGI (*G*_*c*_). In Scenarios 3L and 3U, they observe a future PGI (*G*_*f*_). In Scenario 3L, the PGI’s predictive power is equal to the disease’s SNP Heritability 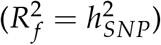, and in Scenario 3U it is equal to its twin heritability 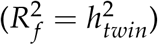.

### 4.3 Estimation for single-disease contracts

For all contracts, one minor complication is that, in the UKB, we do not have long-term follow-up data on most of the non-genetic covariates (only the EHR-data is longitudinal). If we had such follow-up data, we would estimate the model off a population of consumers that we would observe both at age 35 and then again at age 65. The non-genetic covariates would be observed at the former age and disease diagnoses at the latter age. In practice, because of this data limitation, we use data from a population of different ages to estimate the risk of developing each disease by age 65.

For each single-disease contract, estimation is based on the identification Theorem 1, and our method proceeds as follows:

#### Step 1. Estimating the econometric model

We begin by estimating our econometric model for the contract, using our study sample of UKB participants of European-like ancestry. We pool data from females and males, except for breast cancer and prostate cancer. Estimation follows the two-step estimator in Appendix C, which is based on the identification Theorem 1.

#### Step 2. Generating the private risk distributions

For all four scenarios, we generate the distribution of private risk for the disease for all the individuals in our sample. For Scenarios 3L and 3U, we do not observe *G*_*f*_, so for each individual we draw 10 values from its probability distribution conditional on *G*_*c*_ and **W** (given by Lemma B.1 in the Appendix) and use these to generate the risk distribution.

Individuals in the data range in age from 39 to 86. Our set of covariates **W** (see Table 3) includes age as well as a number of age-dependent covariates, such as BMI, blood pressure, hypertension, and whether one ever had menopause (for breast cancer for females) or a prostate specific antigen (PSA) test (for prostate cancer for males). Because age and a number of these covariates are important predictors of risk for all the diseases, we set age to 65 and adjust the most age-dependent covariates to their expected values at age 65 when using Equation 3 to estimate the risk of the disease by age 65. Supplementary Information 2 describes how some covariates were adjusted to reflect this age.

#### Step 3. Identifying the risk classes

To assign the individuals to different risk classes, we use the risk distribution we generated for Scenario 1. That distribution is based on the non-genetic covariates only, which is what insurers use to construct the risk classes. Individuals whose risk was between 0.75 − 1.25 times the average population-wide risk were assigned to the standard risk class.

#### Step 4. Estimating the implicit tax

For each scenario, we use the scenario’s risk distribution from Step 2, limited to those in the standard risk class. We compute Hendren’s implicit tax measure for all percentiles of the distribution. Our main summary statistic of the amount of selection is the minimum implicit tax up to the 80th percentile, denoted as *t*_80_. To obtain 95% confidence intervals, we use the bootstrap method, with 200 draws.

## 5 Results

### 5.1 Single-disease contracts: case study for prostate cancer

We first return to the case study of a prostate cancer CII contract from the introduction. The left panel of Figure 1 shows the risk distributions for prostate cancer under Scenarios 1–3U. Scenario 1 predicts risk using only non-genetic covariates **W**. The distribution is bimodal because family history variables are strong risk factors. There is considerable dispersion in non-genetic risk. Therefore, if insurers had to treat all consumers equally, there would be a noticeable amount of selection from the non-genetic risk factors.

In practice, however, insurers underwrite risk-rated policies and classify consumers into the aforementioned risk classes based on the non-genetic risk factors they observe. We focus on consumers in the standard risk class, shown in blue in Figure 1. Since insurers treat these equally, any variation in risk within the risk class effectively becomes private information. Thus, the blue area in Figure 1, Scenario 1, corresponds to the distribution of non-genetic private risk *P* for the standard risk class.

We now consider selection under the current genetic prediction technology. Figure 1, Scenario 2, shows in blue the distribution of private risk *R*, this time conditional on the covariates and the current PGI, for the consumers in the standard risk class. The current PGI explains 9.9% of the variance in liability for prostate cancer. It adds a considerable 8.3 pp over and above the 22.9% variance explained by the non-genetic covariates. The extra information from the PGI increases the spread of the distribution of private risk, which is no longer bimodal because the explanatory power of the coarse family history dummies is mostly overshadowed by the PGI. Crucially, *even for the consumers in the standard risk class, who are treated equally based on their non-genetic risk factors, the current PGI introduces significant spread in private risk R*. This suggests that, were these consumers to have access to their genetic predictions, there would be a non-negligible amount of selection.

The amount of selection grows considerably with the expected future improvements in genetic prediction technology. Figure 1, Scenarios 3L and 3U, plot in blue the distribution of private risk *R* for the standard risk class with future PGIs. Scenario 3L uses the lower bound for the future PGI’s *R*^2^ based on the SNP heritability, which is 18.0%. Scenario 3U uses the upper bound based on the twin heritability, which is 57.0%.

We can use the implicit tax to summarize the amount of selection. The right panel of Figure 1 plots the implicit taxes in the four scenarios for consumers in the standard risk class. Following Hendren (2013), we focus on the minimum implicit tax up to the 80th percentile, *t*_80_. In Scenario 1, with the non-genetic covariates only, *t*_80_ is equal to 6.3%. This means that a consumer at that risk percentile would have to pay 6.3% more than his actuarially fair price, if he were charged the actuarially fair price for the segment of consumers with equal or higher risk than his. Thus, after underwriting, there is a negligible amount of selection within the standard risk class, as expected.

In Scenario 2, with the current PGI, *t*_80_ is 59.8%. This result implies that there would already be noticeable selection with today’s genetic prediction technology, *if all consumers had adopted this technology*.

In Scenario 3L, with a future PGI that explains as much as prostate cancer’s SNP heritability, adverse selection increases considerably, with *t*_80_ reaching 86.8%. And in Scenario 3U, with predictive power equal to the twin heritability, the amount of selection increases dramatically, with *t*_80_ reaching 426.9%. The implicit taxes in Scenarios 3L and 3U are considerably higher than what Hendren (2013) found for market segments that had completely unraveled and had no trade. This suggests a single-disease CII contract for prostate cancer may not be viable if genetic predictions approach the heritability estimates and are widely available.

### 5.2 Single-disease contracts: main results

We now extend these results to all seven single-disease CII contracts. Though there are nuances across the diseases, the big picture is consistent with the prostate cancer case study. Figure 2 reports the minimum implicit tax up to the 80th percentile of risk (*t*_80_) for each disease and under each scenario for the standard risk class. Table 4 reports detailed results. There are three main findings.

**Table 4:** Summary of main results (standard risk class)

|  | Single-disease contracts |  |  |  |  |  |  | Multiple-disease contracts |  |
| --- | --- | --- | --- | --- | --- | --- | --- | --- | --- |
|  | ALZ | BRC | CAD | CRC | PRC | SCZ | T2D | M only | F only |
| Sex | M & F | F only | M & F | M & F | M only | M & F | M & F | M only | F only |
| Sample size | 405,573 | 211,575 | 410,686 | 410,562 | 181,902 | 410,275 | 410,686 | 175,466 | 204,672 |
| Cases | 3,529 | 18,133 | 54,395 | 10,453 | 15,470 | 1,085 | 34,667 | 58,441 | 45,427 |

| Panel A. Risk of contracting disease ($r$) by age 65, conditional on non-genetic covariates ($\mathbf{W}$) only | | | | | | | | | |
| --- | --- | --- | --- | --- | --- | --- | --- | --- | --- |
| *Full analysis sample* |  |  |  |  |  |  |  |  |  |
| Mean | 0.2% | 7.9% | 8.9% | 1.9% | 5.5% | 0.3% | 6.1% | 25.6% | 17.5% |
| SD | 0.1 pp | 2.1 pp | 8.9 pp | 1.2 pp | 2.0 pp | 1.2 pp | 8.3 pp | 13.1 pp | 8.9 pp |
| *Standard risk class ($r = 0.75$–$1.25$ of sample mean)* |  |  |  |  |  |  |  |  |  |
| Mean | 0.1% | 7.6% | 8.6% | 1.8% | 5.2% | 0.2% | 6.0% | 24.7% | 16.5% |
| SD | 0.02 pp | 1.0 pp | 1.3 pp | 0.3 pp | 0.8 pp | 0.04 pp | 0.9 pp | 3.6 pp | 2.4 pp |

| Panel B. Explanatory power of non-genetic covariates ($\mathbf{W}$) and PGIs | | | | | | | | | |
| --- | --- | --- | --- | --- | --- | --- | --- | --- | --- |
| *Scenarios 1-3* |  |  |  |  |  |  |  |  |  |
| $R^2$ of $\mathbf{W}$ | 38.5% | 6.0% | 36.8% | 11.5% | 22.9% | 20.1% | 39.7% | – | – |
| *Scenario 2* |  |  |  |  |  |  |  |  |  |
| $R^2$ of $G_c$ | 7.1% | 6.7% | 2.5% | 2.2% | 9.9% | 4.9% | 7.4% | – | – |
| $R^2$ of $G_c$ & $\mathbf{W}$ | 43.7% | 12.1% | 37.8% | 13.5% | 31.2% | 22.9% | 43.4% | – | – |
| $\Delta R^2$ of $G_c$ | 5.3 pp | 6.1 pp | 1.0 pp | 2.0 pp | 8.3 pp | 2.8 pp | 3.6 pp | – | – |
| *Scenario 3L* |  |  |  |  |  |  |  |  |  |
| $R^2$ of $G_f$ (SNP $h^2$) | 33.1% | 14.0% | 22.0% | 9.0% | 18.0% | 24.0% | 19.6% | – | – |
| $R^2$ of $G_f$ & $\mathbf{W}$ | 64.6% | 18.9% | 47.6% | 20.1% | 38.2% | 35.9% | 50.1% | – | – |
| $\Delta R^2$ of $G_f$ | 26.2 pp | 12.9 pp | 10.8 pp | 8.6 pp | 15.3 pp | 15.8 pp | 10.4 pp | – | – |
| *Scenario 3U* |  |  |  |  |  |  |  |  |  |
| $R^2$ of $G_f$ (twin $h^2$) | 58.0% | 31.0% | 40.0% | 40.2% | 57.0% | 79.0% | 72.0% | – | – |
| $R^2$ of $G_f$ & $\mathbf{W}$ | 86.8% | 35.3% | 61.5% | 55.7% | 72.9% | 85.4% | 90.7% | – | – |
| $\Delta R^2$ of $G_f$ | 48.4 pp | 29.3 pp | 24.7 pp | 44.2 pp | 50.0 pp | 65.3 pp | 51.0 pp | – | – |

| Panel C. Minimum implicit tax up to the 80th percentile of the disease risk distribution ($t_{80}$) | | | | | | | | | |
| --- | --- | --- | --- | --- | --- | --- | --- | --- | --- |
| *No individual underwriting by insurers (same premium for all)* |  |  |  |  |  |  |  |  |  |
| Scenario 1 | 65.8% | 25.4% | 98.3% | 41.9% | 27.6% | 379.4% | 120.5% | 69.1% | 65.4% |
| *Standard risk class ($r = 0.75$–$1.25$ of full sample mean)* |  |  |  |  |  |  |  |  |  |
| Scenario 1 | 7.0% | 7.2% | 6.3% | 6.3% | 6.3% | 5.8% | 5.8% | 8.9% | 9.0% |
| Scenario 2 | 117.9% | 36.9% | 17.9% | 26.5% | 59.8% | 42.1% | 37.0% | 30.8% | 26.7% |
| Scenario 3L | >1,000% | 57.5% | 64.9% | 61.5% | 86.8% | 178.3% | 71.4% | 54.4% | 45.6% |
| Scenario 3U | >1,000% | 109.8% | 134.2% | 410.1% | 426.9% | >1,000% | >1,000% | 243.9% | 145.6% |

Notes: In Panel A, the reported $R^2$ 's are pseudo- $R^2$ 's on the liability scale (defined in the text). The $R^2$ of the non-genetic covariates and of the current PGI ( $G_c$ ) are estimated in the data; in Scenarios 3L and 3U, the $R^2$ of the future PGI ( $G_f$ ) is assumed to be equal to the disease's SNP heritability and twin heritability, respectively, and the joint $R^2$ of the future PGI and the covariates is obtained from our econometric model. The $\Delta R^2$ of a PGI is its incremental predictive power for the disease, defined as the difference between the $R^2$ of the PGI and the covariates jointly and the $R^2$ of the covariates only. References for the heritability estimates are listed in Supplementary Table 1. The four scenarios are defined in the text.

**Figure 2:**
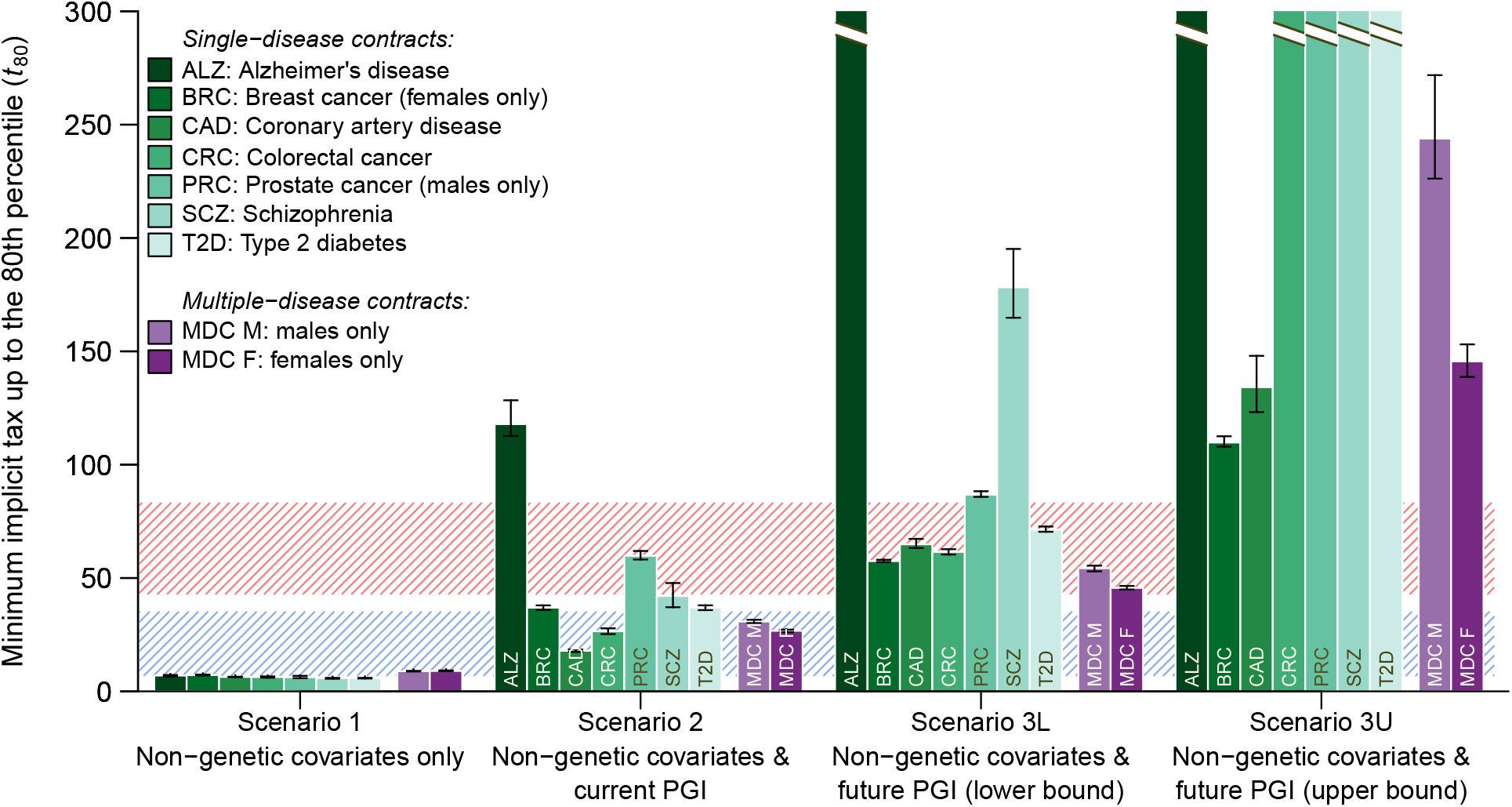
Minimum implicit taxes up to the 80th percentile (*t*_80_) *Notes*: The figure displays *t*_80_ within the standard risk class for the single-disease and multiple-disease CII contracts, in the four scenarios. The striped blue area corresponds to the range of *t*_80_ observed by Hendren (2013) in market segments that had not unraveled, and the striped red area corresponds to the range for market segments that had unraveled. The breaks at the top of the tallest bars indicate that these bars’ heights exceed the figure’s. The results in this figure are also reported in Supplementary Table 7.

First, there would be noticeable selection if the current genetic prediction technology is widely adopted (Scenario 2), in which *t*_80_ ranges from 17.9% for coronary artery disease to 117.9% for Alzheimer’s disease. In comparison with Hendren (2013), *t*_80_ for coronary artery disease (17.9%) and colorectal cancer (26.5%) fall in the middle of the range found for market segments that had not unraveled, and *t*_80_ for breast cancer (36.9%), schizophrenia (42.1%), and type 2 diabetes (37.0%) fall in between the ranges of no unraveling and unraveling. The *t*_80_ of Alzheimer’s disease and prostate cancer are within or above the range observed for segments that had unraveled. The Scenario 2 results suggest that current prediction technology is already sufficiently powerful to cause noticeable selection, and that the main question is when the technology will be adopted widely.

The second main finding is that, with the expected predictive power of future PGIs, the amount of selection could become crippling. In Scenario 3L, where the future *R*^2^’s are conservatively assumed to be equal to the diseases’ SNP heritabilities, *t*_80_ ranges from 57.5% for breast to >1,000% for Alzheimer’s. Some contracts have implicit taxes considerably higher than what Hendren (2013) found even in market segments that had unraveled. The detailed plots in Supplementary Figures 3–10 show that implicit taxes are high throughout the distribution. The histograms of private risk show considerable spread within the standard risk class. In Scenario 3U—in which future PGIs explain as much as the diseases’ twin heritabilities—*t*_80_ is even higher, exceeding 100% for all seven single-disease contracts, and exceeding 1,000% for three. These findings suggest that, with widespread adoption, single-disease CII contracts would not be a viable product.

**Figure 3:**
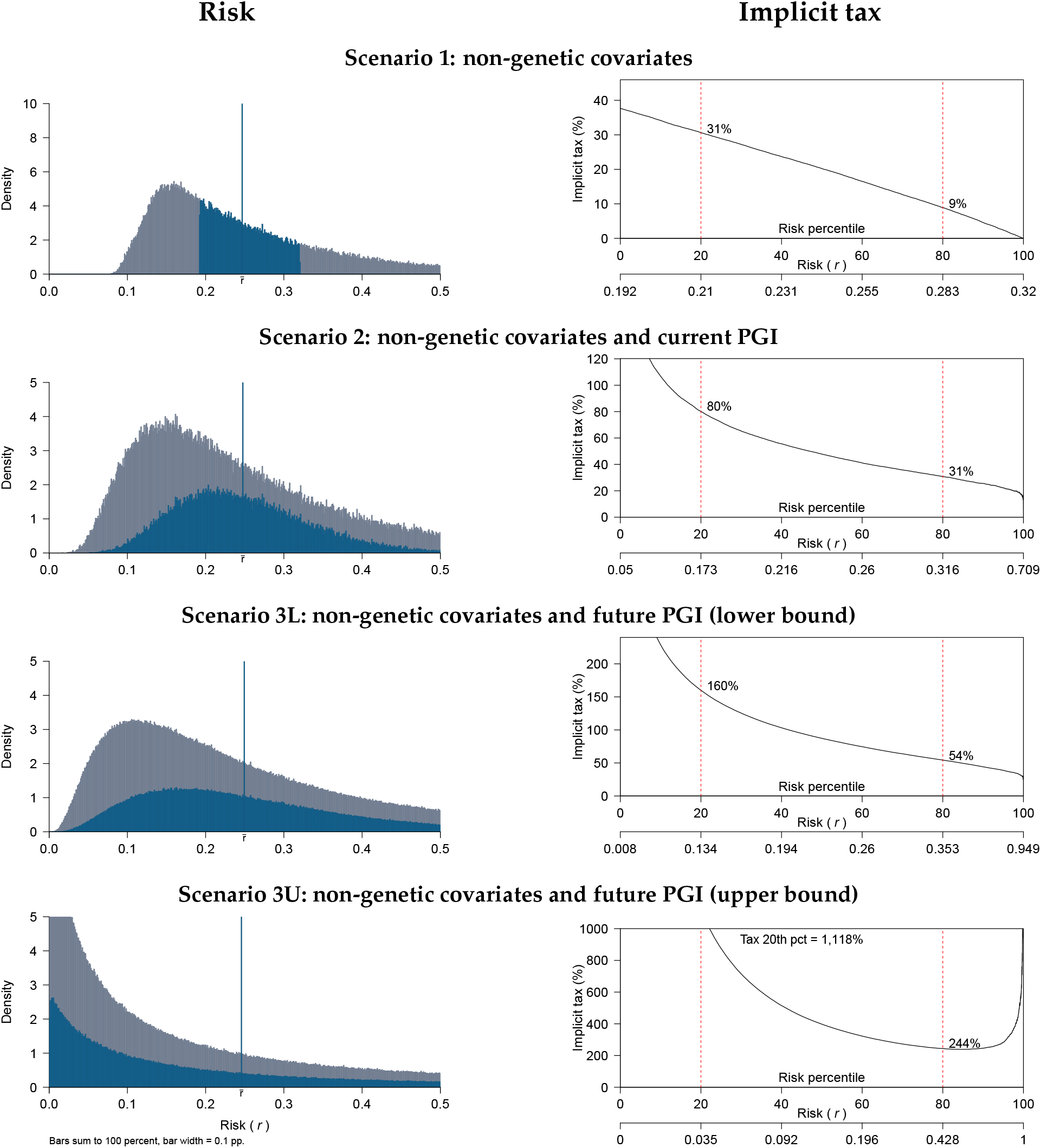
Multiple-disease contract for males: risk and implicit tax. *Notes*: Left panel: distribution of the risk of contracting any of the six diseases in the male multiple-disease contract by age 65, conditional on various information scenarios and computed in the UKB data (*N* = 175, 466). The vertical blue line marks the average risk in the standard risk class 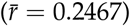. The standard risk class individuals are shown in blue; insurers treat these individuals identically, so the blue distribution corresponds to the distribution of private risk for these individuals. Right panel: implicit tax for consumers in the standard risk class as a function of their percentile private risk of any of the six diseases in the male multiple-disease contract for each scenario.

The third main finding is that there is substantial variation between the diseases. The implicit tax seems to be mainly driven by three factors: the predictive power of the PGI, how much of that predictive power is incremental over that of the non-genetic covariates, and the prevalence of the disease (Table 4). Consider the following examples in Scenario 3L. Alzheimer’s and schizophrenia have the highest SNP *h*^2^ and have low prevalence. Accordingly, they have the highest implicit taxes. At the opposite end of the spectrum, breast and colorectal cancer have the lowest SNP *h*^2^ and their future PGIs have some of the lowest incremental *R*^2^. Accordingly, these two contracts display the lowest *t*_80_.

### 5.3 Multiple-disease contracts

We now consider multiple-disease contracts. We consider separate contracts for males and females that each bundle all the diseases, except breast cancer for males and prostate cancer for females. The contracts pay a lump sum upon any of the covered diseases.

#### Model and empirical implementation

The economic model generalizes to multiple diseases defining the **loss** *L* as a random variable equal to zero or one depending on whether a consumer contracts any of the diseases listed in the contract. The econometric model also extends. This extension requires specifying and estimating the multivariate distributions across the diseases of the genetic disturbance terms *V*, of the current PGI error terms *ϵ*, and of the probit error. Supplementary Information 3 generalizes our model, identification theorem, and estimation procedure. The analysis is nearly identical, except that the formulae are replaced by multidimensional matrix counterparts.

#### Results

Figure 3 displays the risk distributions and implicit taxes for the multiple-disease contract for males in the standard risk class. The average risk is 25.6%, but there is a lot of variation in individual risks, even when conditioning only on the non-genetic covariates (Scenario 1). In Scenario 2, when individuals observe their current PGIs, the spread of the distribution of private risk *R* increases, with *t*_80_ = 30.8% for the standard risk class, implying moderate levels of selection comparable to those observed by Hendren (2013) in markets that had not unraveled.

In Scenarios 3L and 3U, when individuals observe their future PGIs, the distribution of private risk *R* becomes very spread out. In Scenario 3L, with future PGIs that explain as much as the diseases’ SNP heritabilities, consumers at the 5th and 95th percentiles of *R* face risks of 7.8% and 50.4% of contracting a covered disease; in Scenario 3U, with future PGIs explaining as much as the twin heritabilities, the corresponding figures are 0.7% and 91.3%. Thus, with future PGIs, the standard risk class contains a range of individuals— all of whom face the same price of insurance—with very low private risk and others with very high private risk, suggesting ample opportunity for selection. Consistent with this, *t*_80_ = 54.4% in Scenario 3L and 243.9% in Scenario 3U, imply elevated levels of selection that have not been observed in well-functioning market segments.

The results for the multiple-disease contract for females in the standard risk class (Supplementary Figure 3) are qualitatively very similar. Overall, the broad patterns from the single-disease CII contracts hold for the multiple-disease contracts. However, the implicit taxes are often lower in the bundled contracts.

## 6 Robustness and extensions

This section considers limitations of our results, robustness analyses, and extensions.

### 6.1 Robustness analysis in the Health and Retirement Study

The Health and Retirement Study (HRS) is a longitudinal dataset of *∼*20,000 individuals with genotypic, health, lifesyle, and socioeconomic data. Supplementary Information 4 reimplements our model in the HRS, with the goal of assessing the robustness of the results and increasing the ease of replication. The main difficulty in using the HRS is that its health and medical data are not as detailed as those of the UKB. The HRS health data are mostly based on self-reports, whereas the UKB includes EHR data as well as a comprehensive interview by a trained nurse.

We searched the HRS for high-quality data on critical illnesses. The highest quality variable is a question about diagnoses for heart conditions. The conditions include CAD, but also some less severe conditions. For the purposes of our analysis, we define the “HRS CAD” contract as a CII contract for the HRS heart conditions. We repeated our estimation procedure in this sample.

The HRS CAD results are qualitatively similar to the UKB results. We find noticeable selection with the current PRS (*t*_80_ = 18.9%), and large amounts of selection with the future PRS (*t*_80_ = 51.8% in Scenario 3L and 96.0% in Scenario 3U).

### 6.2 Robustness analysis in a calibrated equilibrium model

A major limitation of our analysis is that we consider only private information about risk, ignoring risk preferences and market equilibrium. To check the robustness of our results, Supplementary Information 5 considers a parsimonious calibrated equilibrium model of adverse selection in the CII market. We use the standard supply and demand model of Akerlof (1970) and Einav, Finkelstein, and Cullen (2010).

We model insurance demand using a parsimonious binary loss framework. A loss is assumed to reduce consumption by a fraction Δ*c*. Consumers vary along two fundamental dimensions of heterogeneity: their probability of loss (disease risk) and their relative risk aversion. The cost for firms to provide coverage equals the expected payout plus a fixed cost *F*. We apply the model to the HRS CAD contract. A key advantage of using the HRS is that Kimball, Sahm, and Shapiro (2008) estimated individual-level relative risk aversion based on HRS survey questions. We use their estimate for each consumer’s relative risk aversion. For disease risk, we use our econometric model.

This leaves two parameters to calibrate: Δ*c* and *F*. We calibrate these to match basic CII market statistics from the UK. Based on industry estimates, we set the equilibrium quantity (i.e., the fraction of individual who buy coverage) to 30% and choose fixed costs *F* that imply a loss ratio of 50%. We always restrict attention to consumers in the standard risk class, and perform the calibration in Scenario 1 where the only information is from non-genetic covariates. We refer readers to Supplementary Information 5 for the full model specification, institutional details, and calibration.

#### Results

Figure 4 displays the supply and demand graphs for each scenario. We refer readers to Einav, Finkelstein, and Cullen (2010) for a detailed discussion of these supply and demand curves. In summary, the demand curve represents the percentage of consumers who would like to purchase insurance as a function of the price. The average cost curve *AC* represents the cost of selling coverage to an average buyer, as a function of the fraction of consumers buying insurance. If there is no selection, the cost of selling insurance is independent of the set of consumers buying insurance, and the *AC* curve is flat. With adverse selection, the average cost is downward sloping because it is cheaper to sell to the average consumer in the entire market than to sell to the average consumer in a segment with higher willingness to pay (which correlates with disease risk). Einav, Finkelstein, and Cullen (2010) define the marginal cost curve *MC* as the cost of selling to a marginal consumer.

**Figure 4:**
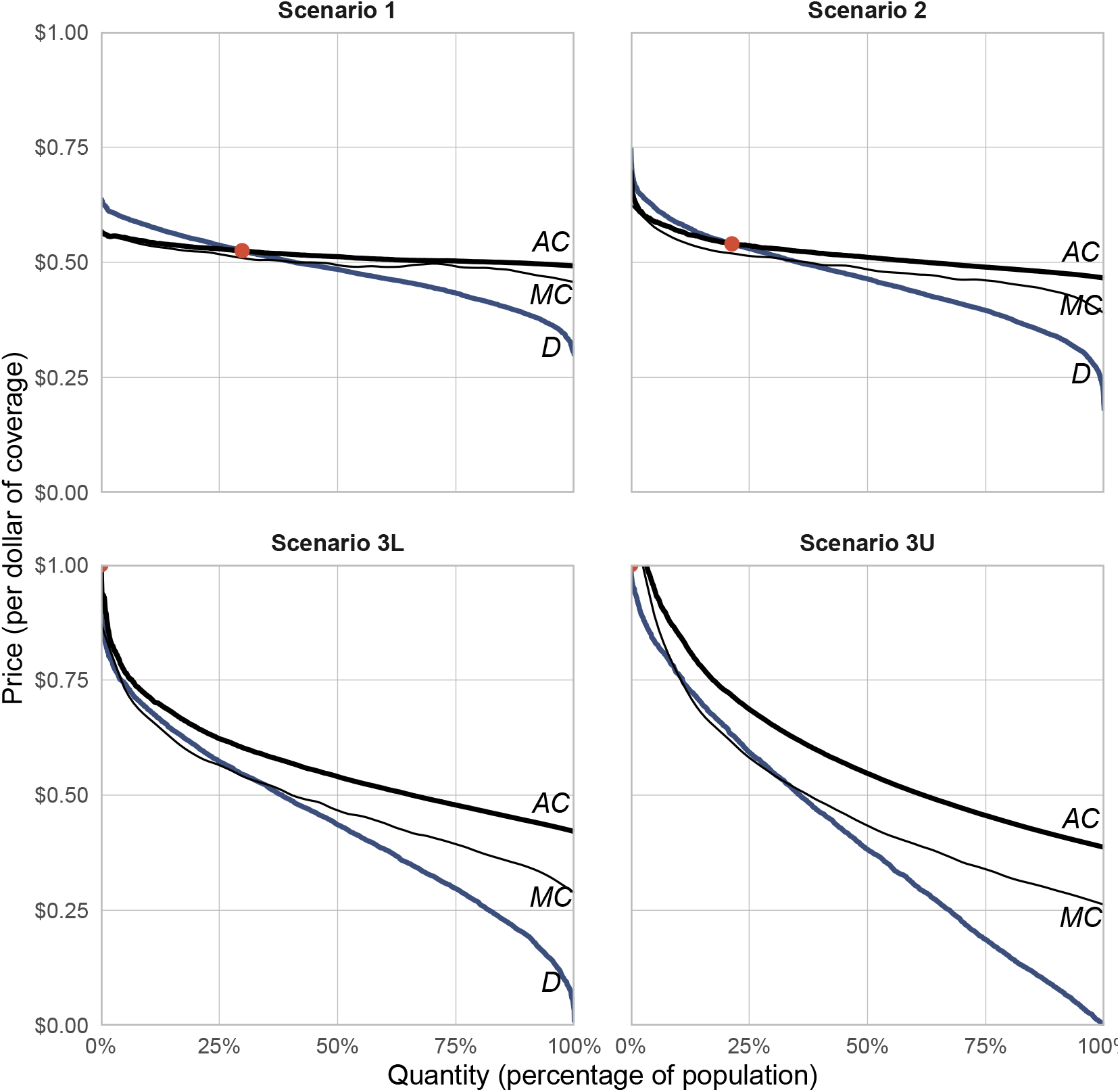
Supply, demand, and equilibria. *Notes:* This figure shows the Einav, Finkelstein, and Cullen (2010) supply (*AC*, black line), demand (*D*, blue line), and marginal cost (*MC*, thin black line) curves as well as the equilibrium point (in red) for CII under the four scenarios. The figure is based on the calibrated equilibrium model for the HRS CAD contract. The four scenarios are defined in the text.

In Scenario 1, where consumers only observe non-genetic covariates, the *AC* curve is nearly flat because there is almost no selection. This is consistent with the definition of the standard risk class. Accordingly, the demand curve is relatively flat because much of the variation in willingness to pay is due to variation in risk aversion rather than in disease risk. The equilibrium quantity is nearly identical to the calibration target of 30% of consumers buying insurance.

In Scenario 2, consumers have access to the current PGI. The *AC* and demand curves rotate and become steeper. The consumers who are most willing to buy insurance are on average more likely to have a loss. These changes affect the market equilibrium. The equilibrium quantity goes down to 21.4%. Thus, giving consumers information about the current PGI creates a noticeable amount of adverse selection.

In Scenarios 3L and 3U consumers have access to future PGIs with predictive power given by the heritability estimates. In both cases, we see a dramatic steepening of the demand curve. The PGI is highly predictive, as seen in Figure 2. This creates a large amount of adverse selection, consistent with our measures purely based on disease risk, such as the implicit tax. Accordingly, in both cases the equilibrium quantity goes to zero. The market suffers from a complete Akerlof (1970) death spiral, and there is no trade. So giving consumers information about a future PGI that reaches the heritability estimates would result in a large, potentially crippling, amount of adverse selection.

We highlight that these results are based on a parsimonious model calibrated from the data, but different equilibrium models may yield different results. For example, if we had assumed that consumers have very high willingness to pay for insurance, and that their demand does not depend on the probability of loss, then the equilibrium quantity would be 100% in all scenarios. We refer readers to Supplementary Information 5 for details on the model and a discussion of alternatives.

### 6.3 Correlation between risk and risk preferences in the UKB

Though the UKB does not include an elicited measure of risk preferences from which we can estimate relative risk aversion, it does includes a self-reported survey item on general risk tolerance. Supplementary Table 6 reports various measures of the correlation between general risk tolerance and each of the diseases we study. For all these measures— including for the genetic correlation, which is not attenuated by measurement error^14^ —we find extremely low correlations. This is consistent with the low correlation between relative risk aversion and disease risk in the HRS and with the results of the calibrated equilibrium model. This further suggests that any correlation between risk and risk preferences is unlikely to meaningfully influence our overall findings in the UKB (e.g., due to advantageous selection; Meza and Webb, 2001).

### 6.4 Partial take-up of genetic testing

Scenarios 2-3U assume that 100% of consumers take up genetic prediction technology. However, there is uncertainty about how common it will become. The modal expert opinion is that it will be common (Our Future Health, 2022). Consistent with this view, major consumer genetics companies and startups have started offering disease risk prediction services and large healthcare providers will inform millions of patients of their genetically predicted disease risk.

Nonetheless, to examine the robustness of our results to assuming lower take-up of genetic testing, we compute our results under two alternative scenarios: 10% in the low take-up scenario and 50% in the medium take-up scenario.

Supplementary Figures 11–13 show the results. As expected, the amount of selection usually goes down with lower take-up. In the low take-up scenario there would not be much selection regardless of the available prediction technology. In the moderate take-up scenario, the amount of selection remains problematically high for all the single-disease contracts in Scenarios 3L and 3U. For the multiple-disease contracts, *t*_80_ is just below Hendren’s unraveling region in Scenario 3L, and is in the unraveling range in Scenario 3U. These results suggest that selection would start being noticeable once the predictive power exceeds the lower future bound and the take-up rate exceeds 50%.

Incomplete take-up generates kinks and non-monotonicities in the implicit tax function. The main reason is that the risk distributions are no longer continuous. In Scenarios 2 and 3, the risk histograms are a mixture of the distribution of non-genetic private risk *P* (for the untested consumers) and the distribution of private risk *R* (for tested consumers). This induces kinks and non-monotone regions in the implicit tax.

### 6.5 Preventive treatment and behavioral changes after testing

One of the main potential benefits of genetic testing is personalized preventive medicine. Genetic tests could be used to identify at-risk patients and offer them preventive cures or inform them of beneficial behavioral changes to mitigate disease risk. Preventive medicine or behavioral changes could in principle decrease the future predictive power of genetic prediction, since they would lower the actual disease risk of individuals with high PGIs. This would reduce the distributional spread of private risk and thus lower the minimum implicit tax *t*_80_ and reduce selection. In practice, this effect is unlikely to substantively affect our main findings. The most common covered diseases are cancers, for which the preventive behaviors have bounded effectiveness. We find very high levels of selection for most contracts. So, given plausible assumptions about the take-up of preventive actions and the effectiveness of these actions, this is unlikely to reduce selection so much as to overturn our main findings.

### 6.6 Firms with an informational advantage

There are a few theoretical papers about insurance markets that assume insurers have private information.^15^ Genetic testing could make such theoretical assumptions plausible in some institutional settings. For example, consider a future world where insurers are allowed to underwrite based on genetics, tests are widely available, but consumers do not have the sophistication to interpret the tests. This would correspond to a model where insurers receive private information but consumers do not.

The simplest and most robust prediction of this type of model is in the competitive case. If insurers just compete their profits to zero, they will offer actuarially fair insurance to each consumer. Thus, even if consumers can’t interpret their genetic tests, they will be able to assess their risk by getting insurance quotes. In such a future world, there is effectively no private information and equilibrium is the same regardless of whether consumers can or cannot interpret their genetic tests. Equilibrium is efficient, in the sense that each consumer is charged her true cost for insurance. But equilibrium can be seen as very unfair, as a consumer who is unlucky in the genetic lottery will face high prices.

### 6.7 Policy implications

Many developed countries currently ban insurers from using genetic information (Nabholz and Rechfeld, 2017). Our findings suggest that, with future genetic prediction technology and widespread genetic testing, these bans will lead to high levels of adverse selection in CII, with the risk of market unravelling for most of the contracts we study.

There is a large amount of research and practical experience on how to regulate selection markets to address market failures due to asymmetric information (Einav, Finkelstein, and Levin, 2010; Einav, Finkelstein, and Mahoney, 2021; Einav, Finkelstein, and Fisman, 2023). The problem of regulating genetic information in insurance is similar. In a nutshell, regulation of selection markets typically seeks to balance two goals: efficiency and redistribution. Efficiency aims to maximize total economic surplus. Redistribution aims to help particular groups, in particular high risk groups who face a higher price of insurance (but also consumers with lower wealth or worse health status). The standard regulatory playbook can be divided in three approaches: laissez-faire, government provision, and managed competition. Here, laissez-faire would involve allowing insurers to observe genetic information and to use it in underwriting, without governmental intervention. This would result in the highest risk consumers facing very high premiums and may be deemed unfair. This could also lead some to forego genetic testing or share their genetic data for research, which could be detrimental to public health. As for government provision, it may not be attractive for a product such as CII that is not seen as essential for many consumers.

Empirical work similar to existing research on other selection markets will be necessary to design optimal policies for the CII market. Nonetheless, a natural policy response that falls under the rubric of managed competition could involve a mix of community rating, subsidies, and risk adjustment. Community rating are rules that limit price discrimination. The current ban on genetic underwriting is a community rating regulation. Community rating improves redistribution across consumers, but generates adverse selection. Some level of community rating may be desirable to help high-risk consumers, but a full ban on genetic price discrimination may be too much due to the large high levels of adverse selection it would cause. Likewise, subsidies might be desirable to increase efficiency and help vulnerable populations, at the cost of spending tax revenue. Finally, risk adjustment are cross subsidies to insurers based on the populations they enroll. Supplementary Information 6 further discusses these standard policy instruments.

## 7 Conclusion

We developed a methodology to measure adverse selection in insurance markets due to genetic prediction of common diseases. We make three marginal contributions. First, we develop a methodology to measure selection based on PGIs, which depend on millions of common genetic variants, as opposed to diagnostic genetic tests for rare mutations. Second, we use heritability estimates to bound the future predictive power of genetic prediction, and measure the incremental predictive power of genetic prediction over and above observable risk factors. This allows us to estimate how selection will change as prediction technology improves. And third, using the UKB’s comprehensive, large-scale micro-data with rich information on non-genetic risk factors, genotypes, and diseases, we apply our method to the CII market. We find that, if genetic testing becomes widespread, there would be noticeable selection with the current prediction technology. The amount of selection would be potentially crippling with the expected future prediction technology.

Genetic prediction is a powerful new technology that has the potential to bring about important benefits, such as personalized medicine and preventive treatments. Understanding the unintended consequences of this technology can help society mitigate its negative effects while still harvesting its benefits. This study aims to help understand some of these possible unintended consequences in insurance markets. Future work should seek to address the above limitations and to apply our methodology to larger markets like life, health, and long-term care insurance.

## Data Availability

All individual-level data analysed in this study are available online at:
https://www.ukbiobank.ac.uk/enable-your-research/apply-for-access
https://hrsdata.isr.umich.edu
GWAS summary statistics data are available online and sources are reported in the Supplementary Information.
Restricted-access results data will be archived with the UK Biobank upon publication.
Open-access results data will be archived at a public data repository upon publication (TBD).

https://www.ukbiobank.ac.uk

https://hrsdata.isr.umich.edu

## Appendix

### A Definition of the implicit tax

Consider an insurance contract that pays $1 if a loss happens, and otherwise charges a premium. For the contract to break even with a consumer with private risk *r*, the premium has to be

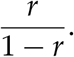

Consider now the case where this policy is purchased by all consumers with private risk *r* or greater. In that case, for the policy to break even the premium would have to be

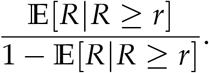

The consumer with private risk *r* would have to pay *T*(*r*) times the actuarially fair price, where

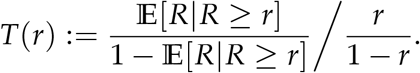

Hendren (2013) calls *T*(*r*) the “pooled price ratio”. To make exposition more intuitive, we define the **implicit tax** *t*(*r*) as

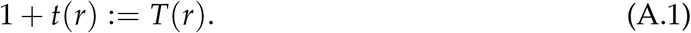

So *t*(*r*) is the implicit tax that a consumer with risk *r* would have to pay due to selection if she were charged an actuarially fair price for all higher-risk consumers.

### B Proof of Theorem 1

#### Lemma B.1.

*Conditional on G*_*c*_ = *g*_*c*_ *and* **W** = **w**, *G*_*f*_ *is normally distributed with mean*

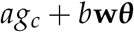

*and variance c*^2^, *where*

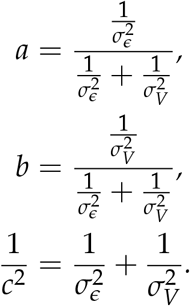

*Proof*. By Assumption 2, conditional on **W** = **w**, *G*_*f*_ = **w*θ*** + *V*, with 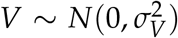. And by Assumption 3, *G*_*c*_ = *G*_*f*_ + *ϵ*, with 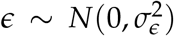. Lemma B.1 then follows from the formula in Gelman et al. (2013, p. 40, Equation 2.10). This is a standard formula from Bayesian statistics.

*Proof of Theorem 1*. We will show that the parameters 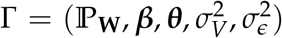 are functions of ℙdata. ℙ_**W**_ is trivial because **W** is observed. We now turn to the other parameters.

#### Part 1: *θ*

Assumption 4 implies that

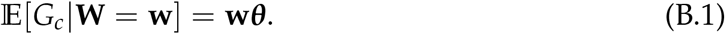

Assumption 1 implies that there is a set of **w**’s in the support of ℙ_**W**_ that span the entire space. So ***θ*** is the vector of coefficients of a regression of *G*_*c*_ on **W**.

#### Part 2: 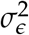

By Assumption 4, there exists a latent variable ℒ for the disease such that

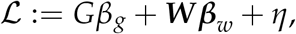

where *η ∼ N*(0, 1), *η ⊥* (***W***, *V, ϵ*), and *L* = {ℒ > 0}, and where the operator {..} is equal to 1 if its argument is true and to 0 otherwise.

Let the pseudo-*R*^2^ of a probit regression of *L* on the current PGI be 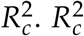 is a function of the data. 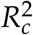 equals

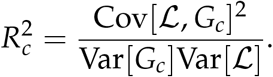

The pseudo-*R*^2^ of a probit regression of *L* on the future PGI is

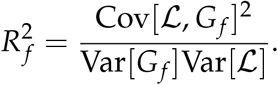

Therefore, using the fact that Cov[ℒ, *G*_*f*_] = Cov[ℒ, *G*_*c*_] and Var[*G*_*c*_] = 1,

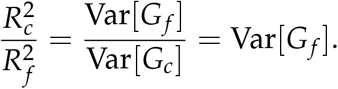

Assumption 3 implies that

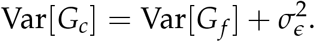

Plugging in the formula above we have

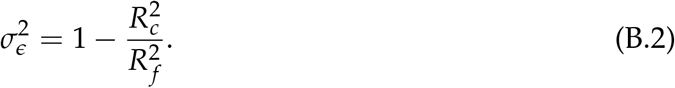

#### Part 3: 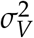

Assumptions 3 and 2 yield

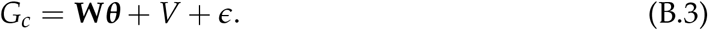

Taking the variance we find so that

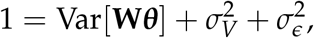

so that

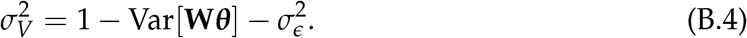

#### Part 4: *β*_*g*_

As mentioned in Part 2, by Assumption 4, we have

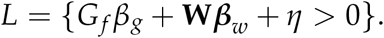

Consider the distribution of *L* conditional on *G*_*c*_ = *g*_*c*_ and **W** = **w**. Lemma B.1 implies that

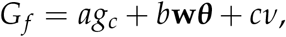

where ν is a standard normal random variable independent of *η*. Therefore, conditional on *G*_*c*_ = *g*_*c*_ and **W** = **w**,

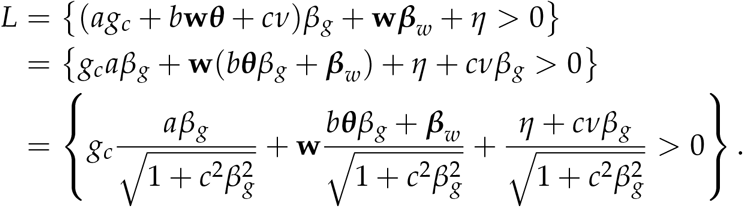

The last term is distributed as *N*(0, 1). Therefore

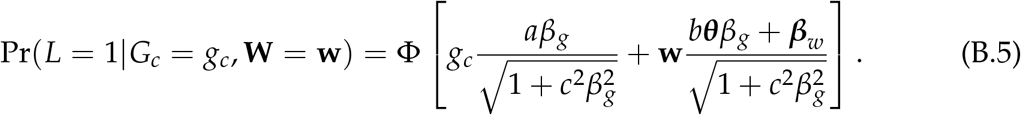

Let ***γ*** be the coefficient vector of a probit regression of *L* on *G*_*c*_ and **W**. Then

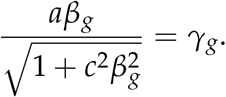

This equation implies that *β*_*g*_ has the same sign as *γ*_*g*_. Taking the square of both sides and solving, we find

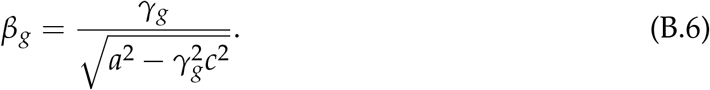

This result implies that |*γ*_*g*_| *< a*/*c*.

#### Part 5: *β*_*w*_

By Equation B.5 and using the fact that the support of **W** includes a set that spans the entire space (Assumption 1),

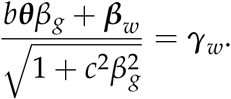

Then

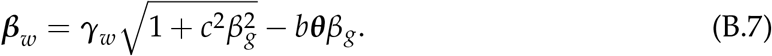

### C Estimation of the single-disease model

Section 4.3 describes how we estimate the single-disease model for Scenarios 1 and 2. For Scenarios 3L and 3U, as also described there, we first estimate 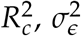, and ℙ_*W*_. For 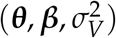, we use maximum likelihood estimation. To simplify the analysis, write the vector of parameters as 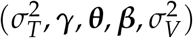, where 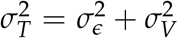 and ***γ*** are the coefficients of the probit regression of *L* on *G*_*c*_ and **W** (Equation B.5). Written this way, ***β*** is a function of the other parameters, as given by Equations B.6 and B.7, and 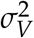 is a function of 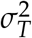.

We can decompose the likelihood function:

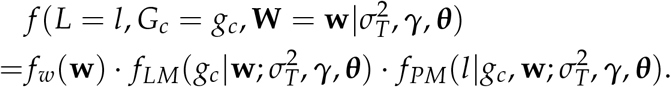

The distribution of *G*_*c*_ conditional on **W** = **w** is given by the linear model (LM) in Equation B.3 and the distribution of *L* conditional on *G*_*c*_ = *g*_*c*_ and **W** = **w** is given by the probit model (PM) in Equation B.5. Therefore, the likelihood can be simplified to

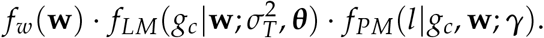

Therefore, the log-likelihood function is a constant plus

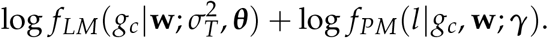

Thus, we can fit the model by maximum likelihood separately for 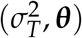 with a linear model and for ***γ*** with a probit model. ***β*** and 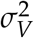 can then be calculated.

For inference, note that the log likelihood has no interaction terms between 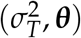 and ***γ***. Therefore, the Fisher information matrix for these parameters is block diagonal, so 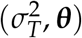 and ***γ*** are asymptotically normal and independent, with the standard errors from their separate estimation. Because ***β*** and 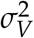 are smooth functions of the other parameters, they are also asymptotically normal. We estimate the full variance-covariance matrix for the parameters 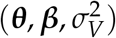 with the delta method. Because we are using maximum likelihood, the estimator is asymptotically efficient.

## Footnotes

1 For instance, in the US, the Genetic Information Nondiscrimination Act (GINA) of 2008 prohibits the use of genetic information by health insurers. Some states have also legislated bans to limit the use of genetic information by other types of insurers. In Canada, the Genetic Non-Discrimination Act (GNDA) of 2017 broadly prohibits genetic discrimination, including in all sectors of the insurance industry. Similarly, in France, the Penal Code prohibits the use of genetic information other than for medico-scientific purposes, and thus by all insurers (Bélisle-Pipon et al., 2019). Some countries—including the UK, the Netherlands, and Sweden—generally prohibit the use of genetic information but permit it for exceptionally large insured amounts or certain disorders, e.g., Huntington’s disease (Nabholz and Rechfeld, 2017).

2 Commonly covered conditions include different types of cancers, coronary artery disease, stroke, multiple sclerosis, paralysis, blindness, and unspecified terminal illness (Gatzert and Maegebier, 2015).

3 Alzheimer’s disease and breast cancer are also polygenic, but in addition to being affected by a large number of variants with tiny effects, they are affected by one common variant (in the *APOE* gene, for Alzheimer’s) or by rare variants (in the *BRCA* genes, for breast cancer) with large effects. Still, the bulk of the genetic variation for Alzheimer’s and breast cancer is due to many variants with small effects.

4 Genotyping is the process of identifying a subset of an individual’s DNA that tags much of the genome’s variation; sequencing is a costlier process to identify every single element of a person’s DNA.

5 A GWAS is a large-scale genetic study in which a trait of interest is regressed on each of millions of genetic variants, separately. As we explain in Supplementary Information 1, the method we use to construct the PGIs, PRS-CS (Ge et al., 2019), computes the estimates 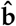 by adjusting the GWAS regression estimates to account for the effects of nearby, correlated SNPs.

6 The GWAS and 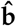 estimates are measures of the variants’ *associations* with the trait, rather than of their effects on the trait (below, for simplicity, we at times nonetheless refer to 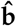 as effects). The PGIs we construct do not therefore have a clear causal interpretation. They are, however, useful for prediction, which is what we need here.

7 The expression SNP heritability comes from the fact that the variants used are typically single nucleotide polymorphisms (SNPs). The SNP heritability estimates in Table 1 come from different sources (listed in Supplementary Table 1, available at https://osf.io/9ndw6/files/osfstorage) and were obtained using different estimation pipelines and sets of common SNPs. They must be seen as ballpark figures rather than precise point estimates.

8 In Canada, between 2006 and 2011, 72% and 20% of claims were due to cancers and cardiovascular diseases, respectively (Canadian Institute of Actuaries, 2014). Cancers and cardiovascular diseases together accounted for between 70% and 87% of claims in the US in 2022 (Gen Re Research Center, 2023) and account for over 80% of claims in the UK (Association of British Insurers, 2022).

9 Our model is similar in spirit to that developed by Becker et al. (2021) for continuous outcomes.

10 To ensure that *G*^*\**^ *⊥ ϵ*, the GWAS sample must not overlap with the UKB.

11 These three cancers account for a little less than half of all cancers in Western countries. For instance, they accounted for 40% of all cancers in the UK between 2016 and 2018 (Cancer Research UK, n.d.).

12 Most vasculatory-disease-related CII claims are due to heart attacks (cardiovascular), followed by strokes (cerebrovascular). Heart attacks are fully captured by our cardiovascular disease variable.

13 Though insurers observe all the non-genetic covariates, they only use these to classify consumers across the risk classes, and so they effectively only observe the risk classes. Because the risk classes are somewhat coarse, there is still non-genetic private risk within each class, but not enough to lead to high levels of selection (that’s why insurers are content to use these coarse risk classes to set prices).

14 We estimate genetic correlations with LD Score regressions (Bulik-Sullivan et al., 2015), which yields estimates that are not attenuated by measurement error.

15 See Brunnermeier, Lamba, and Segura-Rodriguez (2021) and the references therein.

