## Supplementary Information for "Genetic prediction and adverse selection"

First version: October, 2024
This version: May 28, 2025

*The Wharton School, University of Pennsylvania,

†Interdisciplinary Center for Economic Science and Department of Economics, George Mason University,

‡Department of Economics, Leiden University,
School of Business and Economics, Vrije Universiteit Amsterdam,

Supplementary Information files are available at: https://osf.io/9ndw6/files/osfstorage

### 1 Background on genetics, GWAS, and PGIs

#### 1.1 DNA and single-nucleiotide polymorphisms (SNPs)

This study investigates common **complex** (or **non-Mendelian**) **diseases**—i.e., common diseases that are influenced by many genetic variants (and by the environment) (Schork, 1997). Research has established that complex diseases are typically polygenic—i.e., a very large number of genetic variants are involved, most or all of which have only a tiny individual effect (Visscher, Loic Yengo, et al., 2021). Thus, the risk of a complex disease cannot be accurately predicted from only one or a few genetic variants. Even though each variant typically has a tiny effect, the overall genetic effect across all the variants can be considerable. The heritability—defined as the share of the variation in disease liability that is attributable to genetic factors—is often substantial, reaching $\sim$50% on average across complex diseases (Polderman et al., 2015).

The human genome consists of two roughly 3-billion-long sequences of base pairs encoded in DNA (NHGRI, 2023). One of the two sequences is inherited from the mother and the other is inherited from the father. Each sequence consists of four possible nucleotide bases: adenine ("A"), cytosine ("C"), guanine ("G"), and thymine ("T"). Two complementary strands—the "forward" and the "reverse" strand—make up the DNA molecule. They are connected by bonding base pairs like the rungs on a ladder; due to a property called complementarity, A always bonds with T, C with G, and vice versa. This property means that the information content of both strands can be recorded perfectly from a single strand.

The maternal and paternal sequences are each packaged into 23 chromosomes, thus yielding 23 *pairs* of chromosomes in the offspring. One of these pairs contains the sex chromosomes (Chromosome 23). We exclude the sex chromosomes from our analysis because most large genetic association studies omit these (Sun et al., 2023). This decision will not impact our results much because the sex chromosomes generally contribute little to the heritability of complex traits and disease (<5%) (Visscher, Medland, et al., 2006). Each genomic position on the (non-sex) chromosome pairs consists of two base pairs, one that gets passed on by the mother (e.g., A on the forward strand and T on the reverse strand) and one by the father (e.g., G and C on the forward and reverse strand, respectively).

The nucleotides at most positions in the genome are identical across all (or nearly all) humans (NHGRI, 2023). The small fraction of nucleotides that do vary are called single-nucleotide polymorphisms (SNPs), which can be viewed as the smallest unit of genetic variation.[1] At most SNPs, only two possible nucleotide bases have ever been observed among humans on the forward (or reverse) strand. Such SNPs are called "bi-allelic". SNPs with more than two possible nucleotide bases—called "multi-allelic"—do exist but are

[1] Other types of genetic variants exist, including copy number variants (CNVs), insertions, and deletions. Genes are protein-coding sequences of nucleotides. Here, we only focus on SNPs, because they are correlated with much of the variation due to these other types of variants (Sudmant et al., 2015) and capture a large fraction of the heritability of complex traits (Loïc Yengo et al., 2022).

rare and are excluded from our analysis. To code a variable for a given SNP, geneticists arbitrarily select one of the two possible nucleotides, label it as the reference nucleotide, and then sum the number of non-reference (or "effect-coded") nucleotides. The resulting SNP variable thus takes a value of either 0, 1, or 2. For more details, see Beauchamp et al. (2011), Benjamin et al. (2012), or Uffelmann et al. (2021).

#### 1.2 Modeling the genetics of polygenic diseases

As indicated in the main text, we follow standard practice in epidemiological genetics and model a disease $D$ with a liability threshold model, according to which one contracts the disease if one's liability $\mathcal{L}$ is positive. We also assume the additive genetic model (Falconer and Mackay, 1996):

$$
\begin{aligned}
&D = \{\mathcal{L} > 0\}, \\
&\mathcal{L} = k + G^* + \psi = k + \mathbf{X}\,\mathbf{b} + \psi,
\end{aligned}
$$

where $k$ is a constant; $\mathbf{X}$ is a row vector that contains the measured variants, and $\mathbf{b}$ is a column vector that contains their true effect sizes on $D$; $G^* = \mathbf{X}\,\mathbf{b}$ is the individual's true additive genetic factor for $D$; $\psi$ is the disturbance term; and the operator $\{..\}$ is equal to 1 if its argument is true and to 0 otherwise. Thus, $\mathcal{L}$ is the weighted sum of an individual's genetic variants, where the weights are the variants' effect sizes on $\mathcal{L}$, plus a disturbance term that captures all other influences on $D$. It is commonly assumed that $\psi$ follows a normal (or logistic) distribution, in which case the liability threshold model becomes a probit (or logistic) regression of the disease on the liability.

#### 1.3 Polygenic indexes (PGIs)

As explained in the main text, in practice neither $\mathbf{b}$ nor $G^*$ are observed. Instead, one can construct estimates $\hat{\mathbf{b}}$ using estimates from a previously published genome-wide association study (GWAS) of the disease and use them to construct a polygenic index (PGI) $G$.[2] A GWAS is a large-scale genetic study in which a trait of interest is regressed on each of millions of genetic variants, separately. When the trait of interest is a dichotomous disease, probit or logit regressions are used. This relatively simple approach has proven successful in discovering replicable SNP associations. Large-scale GWASs are now discovering novel, replicable SNP associations at an unprecedented rate (Tam et al., 2019). To achieve sufficient sample size, the largest published GWASs are usually meta-analyses of results derived in multiple biobanks. The next section discusses the GWASs from which we obtained the regression estimates needed to construct the estimates $\hat{\mathbf{b}}$

For each individual in the data that passes our sample quality-control procedure (described below), the PGI $G$ for a given disease is the weighted sum of the SNP variables,

[2] PGIs are also commonly called "polygenic risk scores" ("PRSs").

with each SNP variable weighted by its corresponding effect size estimate for the disease:

$$G = \mathbf{X}\,\hat{\mathbf{b}}.$$

Observe that

$$G = \mathbf{X}\,\hat{\mathbf{b}} = \mathbf{X}\,(\mathbf{b} + \boldsymbol{\epsilon}) = \mathbf{X}\,\mathbf{b} + \mathbf{X}\,\boldsymbol{\epsilon} = G^* + \epsilon,$$

where $\boldsymbol{\epsilon}$ is the measurement error in $\hat{\mathbf{b}}$ and $\epsilon = \mathbf{X}\,\boldsymbol{\epsilon}$ is the error in the PGI $G$. As mentioned, complex diseases are influenced by a large number of variants with tiny effects. Thus, by a simple application of the central limit theorem, both $G^* = \mathbf{X}\,\mathbf{b}$ and $\epsilon = \mathbf{X}\,\boldsymbol{\epsilon}$ are approximately normally distributed. Further, because $\boldsymbol{\epsilon}$ (the error in $\hat{\mathbf{b}}$) arises because of sampling variation in the GWAS, and because the GWAS is conducted in a sample that is independent from the analysis sample, $\mathbf{X} \perp \boldsymbol{\epsilon}$ and $G^* \perp \epsilon$.

A given SNP's GWAS estimate comes from a regression of the trait or disease on the SNP (and some baseline controls); that regression usually does not control for the SNPs that are located near the focal SNP in the genome and that are typically correlated with it. Thus, the focal SNP's GWAS estimate captures both that SNP's effect as well as those of nearby, correlated SNPs (such SNPs are said to be in "linkage disequilibrium" with the focal SNP). We computed the PGIs using the software PRS-CS (Ge et al., 2019; Choi, Mak, and O'Reilly, 2020), which models patterns of correlation across SNPs and use these to transform GWAS regression estimates into estimates of $\mathbf{b}$ (i.e., into $\hat{\mathbf{b}}$). PRS-CS also aims to improve the PGIs' signal-to-noise ratio by applying a continuous shrinkage prior in penalized regression to downweight the regression estimates of correlated and/or weakly associated SNPs.

PRS-CS has been shown to perform well for several of our diseases of interest, and about as well as alternative methods like LDpred2 (Privé, Arbel, and Vilhjálmsson, 2020). By default, PRS-CS restricts the set of SNPs to those covered by the reference map of the International HapMap 3 Consortium[3] (Altshuler et al., 2010), and then removes any SNPs with MAF $< 1\%$ in the 1000 Genomes reference panel. This set of SNPs was used to compute all our PGIs except that for Alzheimer's disease. As we further discuss in Supplementary Information 1.5, to tag the well-known *APOE* risk types, our PGI for Alzheimer's had to be augmented by a single additional SNP—rs429358—that is not covered by the otherwise comprehensive HapMap3 set. We ran PRS-CS using its default parameters.

After PRS-CS, we used the PLINK2 software (Chang et al., 2015) to compute the PGIs in the UK Biobank data, while using the PRC-CS adjusted GWAS coefficients as the weights ($\hat{\mathbf{b}}$). Our protocol for computing PGIs is similar to those of recent studies (e.g., Becker et al. (2021) or Ge et al. (2019)) and it follows recent scientific guidelines (see, e.g., Figs 1–2. in Choi, Mak, and O'Reilly, 2020).

It is important to note that the GWAS regression estimates and the transformed estimates $\hat{\mathbf{b}}$ are measures of the SNPs' *associations* with the trait, rather than of their effects

[3] The HapMap 3 SNPs (Altshuler et al., 2010) are a standard set of SNPs used in the PGI literature (Becker et al., 2021), and have been shown to capture the bulk of the heritability attributable to common SNPs (MAF $<1\%$) in populations of European ancestry (Ge et al., 2019; Loïc Yengo et al., 2022).

on the trait. The PGIs we construct do not therefore have a clear causal interpretation. Nonetheless, they are useful predictive tools, which is exactly what is needed for this study. For simplicity, we sometimes refer to $\hat{\mathbf{b}}$ as effects, even though there is no clear causal interpretation.

#### 1.4 GWAS, quality control (QC), and meta-analysis

This section describes how we searched and gathered GWAS "summary statistics", which include the GWAS regression coefficients used as weights to compute $\mathbf{X}\hat{\mathbf{b}}$. Importantly, to avoid overfitting, these summary statistics and GWAS regression coefficients must be estimated in datasets that are independent of the UK Biobank (Wray et al., 2013).

In summary, we computed PGIs using large-sample GWAS summary statistics for our selection of diseases. To quantify the sample size of a GWAS, we rely on the effective sample size metric, "$N_{eff}$", which penalizes the total sample size of an imbalanced case-control regression so that it matches the expected statistical power of a balanced analysis with 50% cases/controls (Grotzinger et al., 2022). The $N_{eff}$ of the GWASs from which we obtained the summary statistics for the seven diseases are shown in Table 1.

Table 1: Summary of the GWAS of the seven diseases of interest

| **Disease** | **Abbreviation** | **N cases** | **N controls** | $Neff$ | **Primary reference** |
|---|---|---|---|---|---|
| Alzheimer's disease | ALZ | 41,197 | 445,030 | 126,275 | J.-C. Lambert et al. (2013) |
| Breast cancer | BRC | 133,384 | 113,789 | 236,094 | Zhang et al. (2020) |
| Coronary artery disease | CAD | 56,424 | 357,721 | 117,486 | Nikpay et al. (2015) |
| Colorectal cancer | CRC | 60,801 | 123,504 | 162,973 | Fernandez-Rozadilla et al. (2023) |
| Prostate cancer | PRC | 79,148 | 61,106 | 137,933 | Schumacher et al. (2018) |
| Schizophrenia | SCZ | 67,390 | 94,015 | 157,013 | Trubetskoy et al. (2022) |
| Type 2 diabetes | T2D | 55,005 | 400,308 | 193,440 | Mahajan et al. (2018) |

*Notes*: Additional details are reported in Supplementary Table 1. Some of the listed GWAS were meta-analyzed with publicly available results derived in the FinnGen Biobank (Kurki et al., 2023). Additional study references and acknowledgments are listed in the Acknowledgements (Supplementary Information 7).

To find the GWAS summary statistics, we searched four prominent public databases hosted by the scientific community. The four databases are the GWAS Catalog (Buniello et al., 2018), the GWAS Atlas (Watanabe et al., 2019), the Polygenic Score Catalog (S. A. Lambert et al., 2021), and the PGI Repository (Becker et al., 2021). The objective was to find the largest published GWAS of each of our diseases of interest. Because the UK Biobank is a standard dataset to include in GWAS meta-analyses, it has become common practice for authors to share, alongside their main results, a hold-out version of their GWAS meta-analysis that excludes the UK Biobank estimates from the meta-analysis. We aimed to find such a hold-out version whenever possible. The latest search was done on January 19, 2024.

For breast cancer, coronary artery disease, prostate cancer, schizophrenia, and type 2 diabetes, we were able to identify publicly available hold-out versions of the largest published GWAS of each of these diseases. However, for Alzheimer's disease and colorectal cancer, we could not find any public hold-out summary statistics, and not all cohort-level files were publicly available. Therefore, for these two diseases, we meta-analyzed the subset of cohort-level files that we could find in the databases, and compensated for the loss of sample size by including recent summary statistics derived by the FinnGen Biobank (Kurki et al., 2023). After the quality control described next, we combined the cohort-level files by fixed-effect meta-analysis using the METAL software (Willer, Li, and Abecasis, 2010).

We applied GWAS quality control (QC) to each downloaded set of summary statistics, as is standard practice (Winkler et al., 2014). (In Supplementary Information 1.6, we also describe how we applied QC to the individual-level genetic data in the UKB.) Our GWAS QC-protocol was based on the often-cited protocol developed by the Social Science Genetic Association Consortium (SSGAC) (Karlsson Linnér, Biroli, et al., 2019; Okbay et al., 2022), which is a continuation of the older "industry-standard" protocol of the GIANT consortium (Winkler et al., 2014). The GWAS QC-protocol serves two main purposes: (1) to remove SNPs that for technical reasons are likely to worsen the signal-to-noise ratio of the PGI (e.g., rare SNPs); and (2) to ensure that SNP coordinates, reference nucleotides, and other per-SNP statistics get aligned across files (e.g., to align them all with the forward strand in the genome reference data).

The protocol was applied using the EasyQC software package (Winkler et al., 2014) and it removed (i) multi-allelic or non-SNP variants (e.g., indels), (ii) SNPs on the sex chromosomes, (iii) SNPs with MAF $<0.5\%$ (the PRS-CS software additionally removes SNPs with MAF $<1\%$ in the reference data), (iv) SNPs that could not be matched with the genome reference data from the Haplotype Reference Consortium (HRC) (McCarthy et al., 2016), and (v) SNPs whose MAF differs by more than 0.2 from the reference frequency in the HRC data. Next, we visually inspected a series of diagnostic plots that were produced by EasyQC, and found no conspicuous issues. Because the imputation quality metric was missing from most of the public summary statistics files, we did not filter on this metric. However, we note that most of the downloaded files were already filtered on imputation quality by the original studies, and that the vast majority of HapMap 3 SNPs are well imputed.

#### 1.5 Alzheimer's disease and the *APOE* region

The *APOE* region on chromosome 19 (coordinates 45,384,477–45,432,606, human reference genome build 37/hg 19) has a well-established large effect on the risk of Alzheimer's disease (Scheltens et al., 2021; J.-C. Lambert et al., 2013). There are six primary risk types of the gene product apolipoprotein E (*ApoE*), called $\varepsilon 2/\varepsilon 2$ through to $\varepsilon 4/\varepsilon 4$. These risk types can be assayed by measuring only two "tag-SNPs" (rs7412 and rs429358) (Faul et al., 2021). The greatest disease risk is conferred by the C nucleotide of rs429358, which has a considerably larger effect than rs7412. The SNP rs429358 is not covered by the comprehensive HapMap3 set, and we therefore augmented the PGI for Alzheimer's disease by including this SNP (see below).

Genetic testing for *APOE* status has been available for years and is commonly included in consumer genetic health reports (Ryan et al., 2021). We model *APOE* status as a component of the PGI (observed by the consumer) rather than as a covariate observable to insurers because insurers typically cannot use that information in underwriting.[4]

Because the tag-SNP rs429358 is missing from the HapMap3 reference set of SNPs, and because the *APOE* region has extensive correlation structure, we modeled *APOE* status as follows. During the GWAS QC, we excluded the genomic region chr19:43,000,000–48,000,000 from the summary statistics for Alzheimer's disease. This 5 megabase (Mb) exclusion region, which covers ∼8.1% of chr 19, was identified by first identifying the furthest SNPs on each side of rs7412 and rs429358 with a pairwise squared correlation of $r^2 >= 0.01$ (computed with the HRC reference data). Thereafter, to be cautious, we expanded the window on each side by one additional Mb, followed by rounding to the nearest preceding/succeeding megabase (i.e., 43Mb and 48Mb). In genetic terminology, we excluded from the PGI for Alzheimer's disease the entire "linkage-disequilibrium region" surrounding the *APOE* region plus a safety margin. Next, we temporarily scored the two tag SNPs separately by their beta's from a probit regression in the UKB, followed by merging this two-SNP PGI with the remaining non-*APOE* genome-wide PGI that fully excluded the *APOE* region, to make our final "augmented" PGI for Alzheimer's disease.

We benchmarked the augmented PGI against the standard approach of modeling the *APOE* risk types using dummy variables. Supplementary Figure 1 shows that the standard dummy-variables approach successfully predicts the risk of disease according to expectation. For our benchmark, we ran a series of probit regressions to evaluate the incremental McKelvey & Zavoina pseudo-$R^2$ from adding each of the following to a specification with only the non-genetic covariates $\boldsymbol{W}$: (i) the dummies, (ii) the dummies and the non-*APOE* genome-wide PGI, or (iii) the augmented PGI. A saturated model that included both the dummies and the augmented PGI was also evaluated. The benchmark showed that the best incremental $R^2$ (5.3 pp) was achieved by the augmented PGI (iii). The saturated model performed marginally worse. Thus, the augmented PGI was selected for use.

#### 1.6 Genotype data, ancestry, and sample quality control (QC)

The analysis reported in the paper was conducted with the harmonized genetic data resource collected, maintained, and distributed by the UK Biobank (Data Category 100319). This resource is described in detail by Bycroft et al. (2018). Imputed genetic data are

[4]Genetic testing to determine *APOE* status is normally considered a predictive genetic test, so genetic information bans typically forbid insurers from observing this test result (Nabholz and Rechfeld, 2017; Dixon et al., 2024), even when that information is included in medical records that would otherwise be permissible for insurers to observe during risk classification.

available for about 487,000 participants. The total number of imputed genetic variants is about 97 million, but the vast majority of these are typically not tested in GWASs because they are rare. There are about 10.3 million imputed variants with MAF $> 1\%$ in the UK Biobank.

We followed standard practice in applied genetic research and restricted our analysis to individuals of European ancestry. There is an unfortunate lack of well-powered GWAS in non-European populations (Ruan et al., 2022), and PGIs constructed using GWAS estimates obtained in samples of European ancestry don't perform well in samples of different ancestries (Martin et al., 2017). In addition to this, to control for population stratification, we include the first 10 principal components (PCs) of the genetic relatedness matrix as control variables (Price et al., 2006; Choi, Mak, and O'Reilly, 2020) when estimating our econometric model.[5] We used the standard set of genetic PCs that are distributed with the UKB genetic data release (Data Category 22009).

To restrict the UK Biobank sample to participants of European ancestry, we relied on the approach described in Karlsson Linnér, Biroli, et al. (2019). Specifically, participants were considered to be of European ancestry if they (i) self-reported their ethnic background to be "White", "White British", "White Irish", or "Any other white background" (Data Field 21000), and (ii) have a value on the first genetic PC that is $\leq 0$. This approach identifies about 448,000 participants that form a tight cluster on the genetic PCs. That cluster is distributed distinctly from the other major population genetic groups, pointing to shared European genetic ancestry Bycroft et al. (2018).

Next, we applied the following sample-level quality control (QC) filters, using the precomputed quality metrics distributed by the UKB with the imputed genetic data resource (Data Category 100313). Participants were removed when (1) there was a mismatch between self-reported and genetic sex (Data Fields 31, 22001), (2) there was evidence of sex-chromosome aneuploidy (Data Field 22019), (3) they were classified as outliers on either genotype heterozygosity or missingness rates based on directly genotyped SNPs (Data Field 22027).

#### 1.7 Inspection of PGI distributions

Supplementary Figure 2 plots the distribution of the PGIs for the seven diseases. We inspected the distributions for (1) departure from normality and (2) sex differences. All PGIs were found to be approximately normally distributed, except for the PGI for Alzheimer's disease. Because of the large risk conferred by the two tag-SNPs for *APOE*, the distribution of this PGI has a noticeable bump to the right. We could not identify any meaningful differences in the PGI distributions across sexes. Nevertheless, we standardized the PGIs so that the have mean zero and unit variance separately by sex.

[5] Population stratification can occur when cultural or environmental differences across populations that impact a disease correlate with genetic differences that are typically non-causal for the disease. This can introduce bias in GWASs and other applied genetic research. For this study, since we are interested in genetic *prediction* rather than *causation*, population stratification is not a major concern. We nonetheless follow standard practice in applied genetic research and control for the top 10 PCs.

#### 2 Health data and risk factors

The UKB has collected self-reported information on a large number of health, psychological, and socioeconomic variables via a touchscreen questionnaire administered at the baseline assessment. This resource is described in detail by Sudlow et al. (2015). The baseline visit occurred in 2006–2010. The touchscreen procedure was followed by a structured follow-up interview by a trained nurse to verify and map the self-reported medical conditions and other health outcome to standardized diagnostic categories, and to run various physiological measurements and lab assays. The most recent version of the study data were refreshed on the UK Biobank Research Analysis Platform on October 22, 2024.

Since the baseline assessment took place, the UKB has been enriched through linkage with several electronic health record data sources. Most of the healthcare in the UK is delivered by the publicly-funded National Health Service (NHS) (Kelly and Stoye, 2020), with which 98% of the population is registered and tracked (Sudlow et al., 2015). Only 7% of the population is covered by private health insurance (Anderson and Mossialos, 2022). Thus, the vast majority of healthcare in the UK is provided and tracked by the NHS (or its subsidiaries). However, the UK has no centralized system specifically for the primary care records from general practices, meaning that the primary care records has worse coverage than the other linked sources described below. For living participants, new information is still being recorded in their electronic health records. The main electronic health record sources linked to the UKB are:

1. Primary care records (Category 3000)
2. Hospital inpatient records (Category 2000)
3. National cancer registries (Category 100092)
4. National death registries (Category 100093).

Some of these data are still in the process of being linked. Unfortunately, because of the UK's decentralized system for primary care records, primary care records are not yet linked for all participants; only about 45% of the participants have been linked thus far. Nevertheless, because the disease outcomes we study are fairly serious, and because the linkage of the cancer and death registries are largely complete, we consider it unlikely that we are missing a substantial number of disease cases because of missing primary care records. However, the UKB participants have been found to be healthier than the general population (Schoeler et al., 2023), so disease cases may be fewer in our data than in the general population.

The data availability of the linked electronic health record sources varies over time. The primary care records go as far back as 1938 for some participants. The hospital inpatient records only go back to 1981 for Scotland, 1991 for Wales, and 1997 for England. The cancer registry goes back to 1957 for Scotland and to 1971 for England and Wales. The death registry captures all deaths among UKB participants after they joined the biobank. Lastly, the self-reported data have the best lifetime coverage because all participants were asked to recall their lifetime medical histories, but this source may suffer from imperfect recall and other such errors. Nevertheless, the combination of self-reported medical history with several distinct electronic health record sources provide good overall coverage of the disease and medical histories of the UKB participants

#### 2.1 Coding the disease outcomes and comorbidities

This section details how we coded the disease outcomes and comorbidities. Our starting point is the "First occurrence of health outcomes" resource (Category 1712), which is maintained and distributed by the UK Biobank. This resource is the result of a harmonization of the self-reported medical data with the linked electronic health record sources (i–iv). However, because the "First occurrence" resource excludes cancer codes, and because it is updated at periodic intervals that differ from the periodic updates of the underlying sources (i–iv), we merged the "First occurrence" resource with the most recent data from all electronic health record sources (i–iv).

The disease outcomes are recorded in the data using the International Classification of Disease (ICD) version 10, censored to first three letters (e.g., C50 "Malignant neoplasm of breast"). To re-code the ICD10 codes into disease outcomes suitable for statistical analysis, we relied on the established and widely used "Phecode" mapping system (Denny et al., 2013; Bastarache, 2021). The Phecode system merges the many subcategories of the ICD-code system into a single general disease outcome. For example, the primary ICD10 code for Alzheimer's disease is G30, while there are sub-codes to distinguish early from late age of onset. The Phecode system merges these sub-types into a single code, 290.11 "Alzheimer's disease". Secondly, it combines redundant or alternative codes spread across ICD chapters. For example, although the official chapter for breast cancer is C50, there are alternative codes that some doctor's may use, such as "D05 Carcinoma in situ of breast". In the latest version, the Phecode mapping system condenses some 90,000 ICD-10 codes into about 1,900 Phecodes (Bastarache, 2021).

The mapping of ICD10 codes to Phecodes for our seven diseases of interest is reported in Supplementary Table 2. With the exception of coronary artery disease, our diseases of interest could all be captured by a single Phecode. To code coronary artery disease, which is defined as a collection of many underlying heart and circulatory conditions, we relied on the approach of the previous literature and considered the entire ICD10 Chapter I20–I25 "Ischaemic heart diseases" (Aragam et al., 2022), which maps to a total of six different Phecodes.

The next section describes the selection of control variables, some of which are comorbidities, e.g., asthma or bipolar disorder. The comorbidities were coded analogously using the Phecode system.

#### 2.2 Coding the epidemiological risk factors and comorbidities

An important contribution of this study is to control for observable information that may correlate with the PGIs. We carefully reviewed the epidemiological literature and clinical guidelines of public health organizations (e.g., the National Cancer Institute or the Centre for Disease Control) to identify the main observable risk factors that are used in clinical practice for screening or diagnostic purposes. This information is typically accessible to insurance companies during medical underwriting. Supplementary Table 3 reports the disease-specific risk factors that were identified. Our resulting list of epidemiological risk factors was verified independently by two medical doctors.

The UKB data are rich enough for us to code almost all the risk factors and comorbidities identified by the review. A handful of risk factors were eventually omitted or proxied (see Panel B in Supplementary Table 3), either because they were unspecific (e.g., diet) or because of limited data availability (e.g., substance use). Instead, the effects of diet were proxied by relevant anthropometric measures or biomarkers, such as BMI, hypertension, systolic blood pressure, and hypercholesterolemia (which are also observable risk factors in and of themselves). Similarly, general psychopathology was proxied by bipolar disorder, depression, and a neuroticism score. Only three risk factors were omitted completely: brain injury, cannabis use, and substance use. Notably, we were able to code family history (father, mother, or siblings) for six of the seven diseases. The exception is schizophrenia, for which family history data is not available and for which we instead used family history of depression as a proxy.

The data fields used to code each risk factor are reported in Supplementary Table 4. We defined a set of nine disease-general risk factors that were included in all the analyses: age (at the most recent observation), sex (omitted from any sex-specific analysis), Townsend's deprivation index, education (years of schooling), BMI, alcohol consumption (drinks per week), smoking (never vs. former vs. current smokers), a dummy variable indicating physical inactivity, and systolic blood pressure. These nine risk factors were always accompanied by the top 10 genetic PCs and by a genotyping array dummy (to control for the fact that part of the sample was genotyped on a different genotyping array).

In addition to these disease-general covariates, we coded risk factors and comorbidities specific to each of the seven disease. These disease-specific covariates were coded either as continuous variables or as dummies, with the exception of BMI and alcohol consumption (drinks per week), which were coded as percentile ranks. The reason is that percentile ranks remain more stable as people age (for more information, see the next section 2.3). Also, because not all female participants have yet undergone menopause, we coded the variable indicating age at menopause (or hysterectomy) as zero for women who had not yet undergone menopause, and then also included a dummy indicating menopause status. Supplementary Table 4 lists the baseline and disease-specific covariates and Supplementary Table 5 reports sample descriptive statistics.

#### 2.3 Adjustment of age-dependent covariate values

An objective of this study is to predict the risk of disease by a particular age (e.g., by age $a = 65$). The data is informative of the age of onset for the medical conditions and comorbidities we study, but for most participants, we only observe their covariate values once (at the baseline assessment in 2006–2010). Therefore, after estimating our econometric model, when predicting disease risk by a particular age, the values of the age-dependent covariates were adjusted for age before being inputted in the prediction model. We did not adjust any time-fixed covariates (e.g., genotyping array dummy) or covariates that are mostly stable in older populations, such as education (years of schooling) or age at first menstruation, nor the handful of geographical variables based on the home address at the time of recruitment, such as the Townsend's deprivation index or air pollution. Supplementary Table 4 lists which covariates were age-adjusted.

To determinate whether each potentially age-dependent covariate indeed depended on age and should thus be adjusted, we regressed each of the covariates separately on a fourth-degree polynomial of age. Whenever the polynomial explained more than 1% of the variation, the covariate was adjusted in the prediction step. Otherwise, we used the observed value both in the estimation and the prediction steps. Also, to be consistent across our seven diseases of interest, the family history variables were always age-adjusted, though we found that it did not depend on age for some diseases.

The age-adjustment procedure was done separately by sex. For continuous covariates, with the exception of "age at menopause (or hysterectomy)", we first ran a linear regression of each covariate $W$ on the age polynomial to model the covariate as a function of age $a$: $W(a) = a_1 \times a + a_2 \times a^2 + a_3 \times a^3 + a_4 \times a^4 + e$. The estimated regression coefficients were then used to predict the values of the covariate $\bar{w}(a)$ for each year of age $a$ observed in the data (i.e., 39–86 years). Then, for each covariate, we adjusted the value at age 65 for participant $i$ observed at age $a_i$ by adding the predicted difference between $\bar{w}(65)$ and $\bar{w}(a_i)$:

$$w_i(65) := w_i(a) + [\bar{w}(65) - \bar{w}(a_i)].$$

For binary covariates, with the exception of "ever menopause (or hysterectomy)", we proceeded analogously, but estimated probit rather than linear regressions and projected probabilities. We only adjusted probabilities among participants with age $a_i < 65$ who had not experienced the event of the covariate, as well as among individuals with age $a_i > 65$ who had experienced the event but for which we could not observe the age of the event. We did not adjust the covariate values of participants with age $a_i < 65$ who had already experienced the event before age 65, nor of participants of $a_i > 65$ and who had not (yet) experienced the event.

To illustrate the procedure for binary covariates, consider a woman of age $a_i = 40$ who had not yet experienced hypertension. For that woman, the observed covariate value is $x_i(40) = 0$. This was adjusted to $w_i(65) = 0 + [0.20 - 0.02]$, where $[0.20 - 0.02)] = 0.18$ is the difference between the probabilities of having hypertension at ages 65 and 40, which we assume to be the probability of developing hypertension between ages 40 and 65 conditional on not having hypertension at age 40. Similarly, the covariate for a woman of age 80 and who had experienced hypertension was adjusted to $w_i(80) = 1 + [0.2 - 0.6] = 0.6$, to reflect the fact that the woman may not have had hypertension at age 65.

For the covariates "age at menopause (or hysterectomy)" and "ever menopause (or hysterectomy)", all female participants older than age 65 who had not yet experienced the event were recoded as having experienced the event at age 65. The reason is that almost all women have either reached menopause (or had a hysterectomy) before this age.

We found that none of the age-dependent covariates in this study decreases as a function of age. Instead, in most cases, the age polynomial predicted a monotonic increase in the covariate. However, for a few covariates, the polynomial predicted slight non-monotonicity at the top and bottom of the age distribution, where there are much fewer observations. Therefore, we forced monotonicity over the entire age range by fitting a cubic smoothing spline as implemented in the R package "Shape Constrained Additive Models". The smoothing had little to no effect on most age groups in the data.

Because all covariates were observed to increases as a function of age, the age-adjustment procedure effectively increased the values of the age-dependent covariates for participants below 65, and decreased the values for those older than 65.

### 3 Generalized econometric model for multiple-disease contracts

#### 3.1 Models for multiple-disease contracts

We now discuss how we model multiple-disease CII contracts. The theoretical model in Section 3.1 of the main text accommodates this case, as the loss can be defined a the occurrence of any of the diseases. There are two basic ways of implementing this model empirically. The first is to use a PGI for the bundle and proceed as in the single-disease case. The second is to formally model the co-occurrence of the multiple diseases. Here, we pursue this second route, by extending the econometric model to multiple diseases.

For ease of exposition, in this section only, we modify our convention to write vectors in bold. We follow that convention everywhere else, but here we write matrices rather than vectors in bold.

#### 3.2 The model

We now generalize our model to the case where there are $\mathcal{D} > 1$ diseases. Each agent in now characterized by a tuple

$$(D, G_c, G_f, W).$$

The only difference is that $D$, $G_c$ and $G_f$ are now column vectors with coordinates indexed by disease $d = 1, \dots, \mathcal{D}$. The model is fully specified by the joint distribution $\mathbb{P}$ of all variables. We assume that, for each dimension $d$, Assumptions 1-5 from the main text hold.

We begin by defining the key equations of the model in matrix notation. All vectors are column vectors. Main text Equation 1 becomes

$$G_f = \boldsymbol{\theta} W + V, \tag{1}$$

where

$$G_f = (G_{f,1}, \ldots, G_{f,\mathcal{D}})^T,$$

$$\boldsymbol{\theta} = \begin{bmatrix} - & \theta_{w,1}^\top & - \\ - & \theta_{w,2}^\top & - \\ & \vdots & \\ - & \theta_{w,\mathcal{D}}^\top & - \end{bmatrix},$$

$$W = (W_1, \ldots, W_p)^T,$$

and

$$V = (V_1, \ldots, V_{\mathcal{D}})^T,$$

and where $p$ is the cardinality of the set of covariates. In accordance with our notational convention for this section only, we here bold matrices rather than vectors. $G_f$ is a $\mathcal{D} \times 1$ vector; $\theta$ is a $\mathcal{D} \times p$ matrix; $W$ is a $\mathcal{D} \times 1$ vector of covariates.

Main text Equation 2 becomes

$$G_c = G_f + \epsilon, \tag{2}$$

where

$$G_c = (G_{c,1}, \ldots, G_{c,\mathcal{D}})^T$$

$$\epsilon = (\epsilon_1, \ldots, \epsilon_{\mathcal{D}})^T.$$

Because Assumptions 1-5 hold for each dimension, there exist latent variables $\mathcal{L}_d$, $d = 1, \ldots, \mathcal{D}$, such that:

$$\mathcal{L}_d = \beta_{g,d} G_{f,d} + \beta_{w,d} W + \eta_d, \tag{3}$$

$$D_d = \{\mathcal{L}_d > 0\}.$$

We can write

$$\mathcal{L} = \beta_g G_f + \beta_w W + \eta,$$

$$D = \{\mathcal{L} > 0\},$$

where

$$\mathcal{L} = (\mathcal{L}_1, \ldots, \mathcal{L}_{\mathcal{D}})^T,$$

$$D = (D_1, \ldots, D_{\mathcal{D}}),$$

$$\boldsymbol{\beta}_g = \text{diag}(\beta_{g,1}, \ldots, \beta_{g,\mathcal{D}}),$$

$$\beta_{w,d} = (\beta_{w,d,1}, \ldots, \beta_{w,d,p})^T,$$

$$\boldsymbol{\beta}_w = \begin{bmatrix} - & \beta_{w,1}^\top & - \\ - & \beta_{w,2}^\top & - \\ & \vdots & \\ - & \beta_{w,\mathcal{D}}^\top & - \end{bmatrix},$$

$$\eta = (\eta_1, \ldots, \eta_{\mathcal{D}})^T.$$

Note that the set of relevant covariates differs across disease; when a covariate $W_k$ is not used for a disease $d$, the corresponding coefficient $W_{w,d,k}$ is 0.

We need the following natural extension of the normality and independence assumptions from the one-disease case.

**Assumption 1** (Multidimensional Assumptions)**.** *We have that*

- $(\eta, \epsilon, V)$ *are multivariate normal.*
- $\mathrm{Cov}[\epsilon]$ *is diagonal.*
- $\eta$*,* $\epsilon$*, and* $V$ *are orthogonal from each other.*

We allow the covariance matrices $\mathrm{Cov}[\eta]$ and $\mathrm{Cov}[V]$ to be non-diagonal, so that the genetic shocks $V$ and non-genetic shocks $\eta$ can have correlations across diseases. Since the PGIs for the different diseases were constructed using summary statistics from GWASs that were conducted in mostly independent datasets, it is reasonable to assume that $\mathrm{Cov}[\epsilon]$ is diagonal.

#### 3.3 Identification Theorem

We now prove that the model is identified. Theorem 1 in the main text implies that we can identify almost all parameters in the model by considering each disease separately. The only parameters for which identification still needs to be established are the off-diagonal terms in $\mathrm{Cov}[\eta]$ and $\mathrm{Cov}[V]$. Estimating $\mathrm{Cov}[V]$ is simple because, by equations (1) and (2),

$$G_c = \boldsymbol{\theta} W + V + \epsilon$$

so that

$$\mathrm{Cov}[V] = \mathrm{Cov}[G_c - \boldsymbol{\theta} W] - \mathrm{Cov}[\epsilon]. \tag{4}$$

To see how $\mathrm{Cov}[\eta]$ is identified, we use the multivariate version of the Bayesian updating lemma:

**Supplementary Information Lemma 1.** *Conditional on* $G_c = g_c$ *and* $W = w$*,* $G_f$ *is normally distributed with mean*

$$\boldsymbol{A} g_c + \boldsymbol{B\theta} w$$

*and variance*

$$\boldsymbol{C}\boldsymbol{C}^T.$$

*The constants are given by the precision matrices*

$$\begin{aligned}\mathbf{\Lambda}_\epsilon &:= \mathrm{Cov}[\epsilon]^{-1}\\ \mathbf{\Lambda}_V &:= \mathrm{Cov}[V]^{-1}\\ \mathbf{\Lambda} &:= \mathbf{\Lambda}_\epsilon + \mathbf{\Lambda}_V.\end{aligned}$$

*as*

$$\begin{aligned}\boldsymbol{A} &= \mathbf{\Lambda}^{-1}\mathbf{\Lambda}_\epsilon\\ \boldsymbol{B} &= \mathbf{\Lambda}^{-1}\mathbf{\Lambda}_V\\ \boldsymbol{C}\boldsymbol{C}^T &= \mathbf{\Lambda}^{-1}.\end{aligned}$$

*Proof.* A proof is in Section 1.7.2 of Soch et al. (2024). Our formula corresponds to the particular case here their $X$ is a $\mathcal{D} \times \mathcal{D}$ identity matrix. The covariance matrix ($\boldsymbol{C}\boldsymbol{C}^T$) can be decomposed in this form by the Cholesky decomposition. □

Therefore, conditional on $G = g_c$ and $W = w$, $G_f$ is distributed as

$$\boldsymbol{A}g_c + \boldsymbol{B}\boldsymbol{\theta}w + \boldsymbol{C}\nu, \tag{5}$$

where $\nu$ is a standard normal $\mathcal{D} \times 1$ vector. Therefore, the conditional distribution of the latent variable $\mathcal{L}$ is

$$\begin{aligned}\mathcal{L} =& \boldsymbol{\beta}_g G_f + \boldsymbol{\beta}_w w + \eta\\ \mathcal{L} =& \boldsymbol{\beta}_g(\boldsymbol{A}g_c + \boldsymbol{B}\boldsymbol{\theta}w + \boldsymbol{C}\nu) + \boldsymbol{\beta}_w w + \eta\\ \mathcal{L} =& \boldsymbol{\beta}_g \boldsymbol{A}g_c + (\boldsymbol{\beta}_g \boldsymbol{B}\boldsymbol{\theta} + \boldsymbol{\beta}_w)w + (\boldsymbol{\beta}_g \boldsymbol{C}\nu + \eta).\end{aligned} \tag{6}$$

Therefore, conditional on $g_c$ and $w$, the covariance matrix of $\mathcal{L}$ is

$$\mathrm{Cov}[\mathcal{L}|G = g_c, W = w] = \boldsymbol{\beta}_g \boldsymbol{C}\boldsymbol{C}^T \boldsymbol{\beta}_g + \mathrm{Cov}[\eta]. \tag{7}$$

With this observation, we can extend the identification theorem to the multiple-disease model. The definition of identification is identical to that for the one-disease case.

**Theorem 1.** *Under assumptions (1)-(6), the multiple diseases model is identified.*

*Proof.* The argument above shows identification of all parameters except for the off-diagonal terms of $\mathrm{Cov}[\eta]$. Choose $w$ in the support of $\mathbb{P}_W$ and $g_c$ and let

$$m := \boldsymbol{\beta}_g \boldsymbol{A}g_c + (\boldsymbol{\beta}_g \boldsymbol{B}\boldsymbol{\theta} + \boldsymbol{\beta}_w)w.$$

Then, conditional on $G_c = g_c$ and $W = w$, $\mathcal{L}$ is multivariate Gaussian with mean $m$ and covariance given by Equation 7. Therefore, the probability that $D_1 = D_2 = 1$ is the probability that the projection to the first two coordinates of a bivariate Gaussian with mean $m$ and covariance matrix given by Equation(7) is in the first quadrant. This probability is increasing in the $\mathrm{Cov}[\eta]_{12}$.[6] Therefore, $\mathrm{Cov}[\eta]_{12}$ is identified. The same argument also implies identification of all the other off-diagonal terms. □

##### 3.4 Estimation of the econometric model

The estimation of the multiple-disease econometric model proceeds as follows. First, we estimate the single-disease model for each disease. This yields estimates for all parameters except for the off-diagonal terms in $\mathrm{Cov}[V]$ and $\mathrm{Cov}[\eta]$. We estimate $\mathrm{Cov}[V]$ with Equation 4, where we use the sample analogue of $\mathrm{Cov}[G_c - \boldsymbol{\theta} W]$. To estimate $\mathrm{Cov}[\eta]$, we use Equation 7. The joint distribution of the data given $\mathrm{Cov}[\eta]$ and known parameters is given by the multivariate probit model. We fit $\mathrm{Cov}[\eta]$ by maximum likelihood.

##### 3.5 Generating the private risk distributions

To generate the private risk distribution for each of the four scenarios we consider, we generalize the approach for the single-disease contracts (see Section 4.3 of the main text) to the case of a multiple-disease contract. We consider a contract that pays out in case any of the diseases occurs, and calculate risks accordingly.

We start from a dataset that includes the covariates $W$ and the vector of current PGIs $G_c$. We then use equation (5) to draw values of $G_f$ according to its conditional expectation given observables. We draw 10 simulated observations per original observation. We then calculate the genetic and non-genetic risks based on the covariates, current PGIs, and the simulated future PGIs.

#### 4 Robustness analysis with the Health and Retirement Study

Our main analysis uses the UKB, which is an ideal dataset that combines large sample size, genetic information, and detailed health information from exams, surveys, and the NHS health records. In this section, we reproduce our main results using the Health and Retirement Study (HRS) dataset. The HRS is a longitudinal study of older adults in the United States. The HRS is a well-known dataset that has been used in many studies of insurance and health. The goal is to assess the robustness of our findings in a different setting and with a widely used dataset. While any interested researcher can apply to use

[6] We can show that this probability is increasing as follows. Formula 26.3.19 of Abramowitz and Stegun (1948) shows that the probability of the first quadrant for a standard bivariate Gaussian is

$$\frac{1}{4} + \frac{\arcsin \rho}{2\pi}.$$

This is increasing in the correlation coefficient $\rho$.

the UK Biobank, the process is more involved than for the HRS. Thus, the HRS analysis makes it easier for other researchers to replicate our results.

In our analysis, the main difficulty in using the HRS is the lack of electronic health records information. This makes it more difficult to reliably measure insurance losses that are relevant for critical illness insurance. We searched the HRS for proxy variables for the relevant insurance losses and performed basic quality checks. The most relevant proxies are built from questions about whether a doctor has told the respondent that they have a certain disease. Out of these variables, the one about heart problems was the most reliable in our quality checks. This variable is `RwHEART` in the RAND longitudinal files, where `w` is the wave number. The documentation for the RAND longitudinal file 2020v2 indicates (at p. 431) that `RwHEART` captures the following conditions: "heart attack, coronary heart disease, angina, congestive heart failure, or other heart problems". This definition is broader than CAD, but it is the closest available high-quality proxy in the HRS.

#### 4.1 The HRS CAD contract and data construction

We use the `RwHEART` variable to define the loss for a critical illness contract that we term the "HRS CAD" contract. Because the HRS's `RwHEART` definition is broader just CAD, we expect it to have a higher probability of occurrence and to be broader than the definition in real critical illness policies.

The genetic predictor we use is the CAD polygenic index from Karlsson Linnér and Koellinger (2022). We estimate its $R^2$, controlling for gender and age, to be 3.5% in the HRS. We also experimented with using the most up-to-date CAD PGI available in the HRS, `E5_MI_CARDIOGRAM15` (CARDIoGRAM 2015). This is available as part of the HRS's sensitive health information. `E5_MI_CARDIOGRAM15` is, however, a considerably older and less powerful PGI. We estimate its $R^2$ to be about half that of the newer PGI we use. Nevertheless, using `E5_MI_CARDIOGRAM15` instead of our newer PGI yields similar results for the amount of selection with the future PGI (although less selection with the current PGI, as expected). The noticeable difference in performance between `E5_MI_CARDIOGRAM15` and the newer PGI we use from Karlsson Linnér and Koellinger (2022) illustrates the rapid speed of advances in the quality of available PGIs.

For non-genetic covariates, we use the HRS variables that correspond as closely as possible to our UKB covariates. We use age, gender, BMI, smoking status, alcohol consumption, physical activity, cholesterol, and earnings income. See the replication code for the corresponding HRS variable names. For the SNP and twin heritability assumptions for Scenarios 3L and 3U, we used the same values as in the UKB analysis.

The final estimation sample includes 10,460 individuals and 39,137 individual-wave combinations. We conduct the same analysis as in the main text. After estimating the model, we simulate the risk distribution under the same informational assumptions as in the main text. For the simulations, we used a subsample of the 7,183 individuals that also had answered questions on risk preferences. We used risk preference information in our robustness analysis in Section 5. We fix this sample to facilitate comparison between both sections.

##### 4.2 Results

Figure 1 displays the risk distribution for the HRS CAD contract under the different information scenarios. The results are qualitatively similar to the UKB results for CAD. The main quantitative difference is that mean risk is higher than in the UKB CAD contract, which is as expected given the broader definition of the HRS variable. The right panel displays the implicit tax for the HRS CAD contract. These are similar to those in the UKB. As before, we find noticeable selection in the scenario where the current PGI is widely known to consumers ($t_{80}$ = 18.9%). With future genetic prediction technology, we find selection in Hendren's unraveling range ($t_{80}$ = 51.8% in Scenario 3L and 96.0% in Scenario 3U). This is at the lower end of our findings across the different contracts we study in the UKB, but still within Hendren's unraveling range.

#### 5 Robustness analysis in an equilibrium model

Equilibrium in most adverse selection models depends both on the distribution of risk and on risk preferences. However, the main text studies the distribution of risk given different sets of information, without considering risk preferences. This section checks whether the main results are robust to considering risk preferences in a calibrated equilibrium model. We check the robustness of the main qualitative conclusions: that selection due to genetics would be noticeable with the current PGI and would be high with future PGIs.

We consider calibrated equilibrium models of adverse selection in the CII market. To reduce researcher degrees of freedom, we use a parsimonious calibrated model, similar in spirit to the exercise in Brown and Finkelstein (2008). The model can be calibrated both for the contracts our main UK Biobank analysis focuses on and for the HRS CAD contract. We report the results for the HRS CAD contract for two reasons. First, the HRS data includes questions eliciting relative risk aversion. Thus, we can use the data to inform the joint distribution of risk and risk preferences, which is known to impact the degree of selection problems (Fang, Keane, and Silverman, 2008). Second, the HRS CAD contract is the least adversely selected in our analysis, so this is a more stringent robustness check.

##### 5.1 Model

We consider a simple binary loss insurance model. We use the standard competitive equilibrium concept from Akerlof (1970) and Einav, Finkelstein, and Cullen (2010). The model uses its own notation, independent of the other sections. The model is as follows.

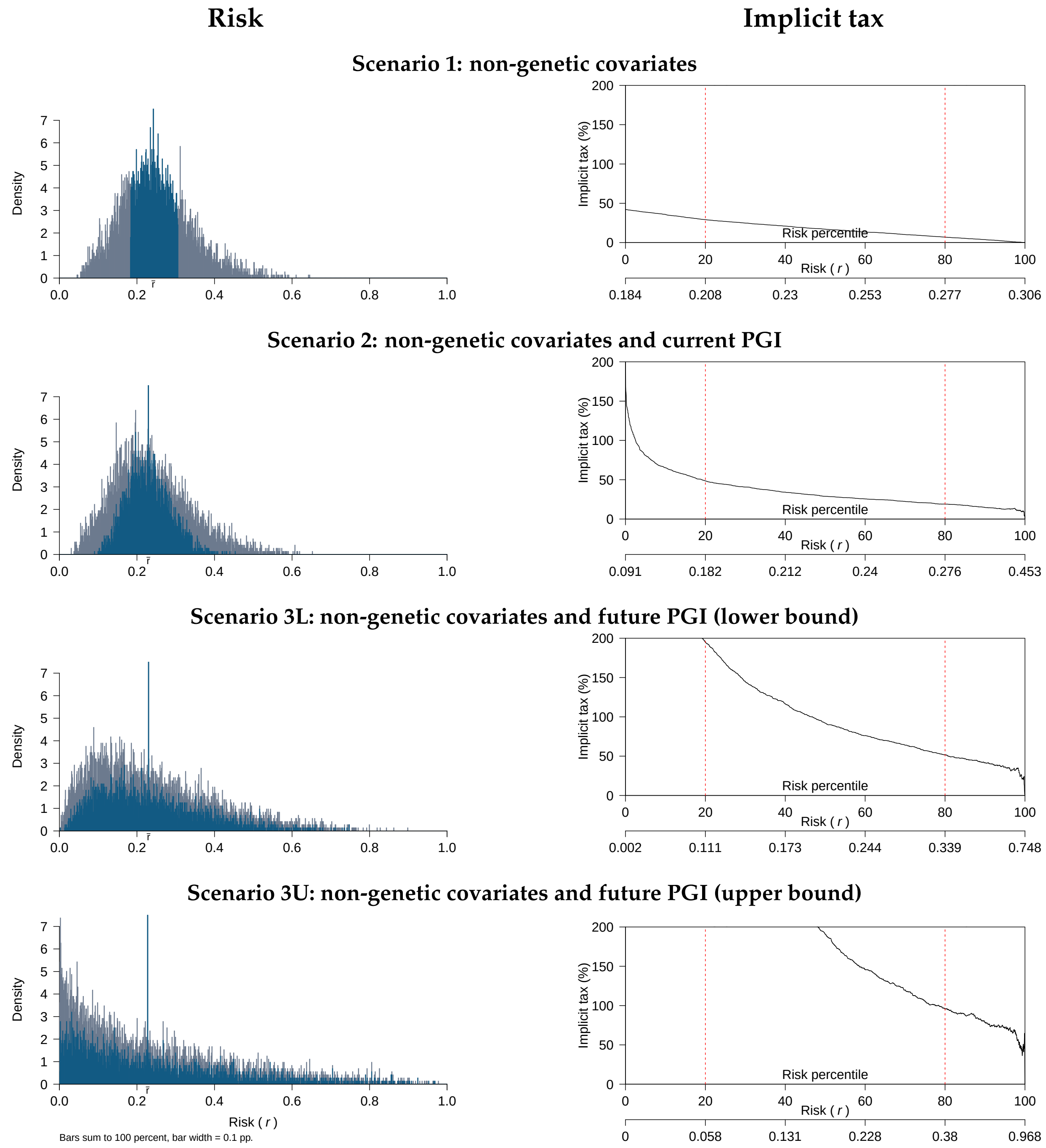


Figure 1: HRS CAD contract: risk and implicit tax

*Notes*: Left panel: distribution of the risk of a loss based on the HRS CAD variable. Right panel: implicit tax for consumers in the standard risk class as a function of their percentile private risk for each scenario.

**Consumers** $i$ in $I$ are heterogeneous in their **probability of loss** $\pi_i$ and **relative risk aversion** $\gamma_i$. The consumer's income equals $Y$ if no loss is incurred, and $(1-\Delta c)Y$ if a loss is incurred. $i$ is uniformly distributed over the unit interval. The consumer can purchase an insurance contract that pays \$1 if a loss is incurred, and \$0 otherwise. The **price** of the contract is $p$.

**Demand.** We assume that the policy payout is considerably smaller than the consumer's income. So the consumer's net gain from purchasing the contract is given by the difference in marginal utility between the two states. That is, the net gain is

$$\pi_i \, \mathrm{MU}_{\mathrm{low},i} \cdot (1-p) + (1-\pi_i) \, \mathrm{MU}_{\mathrm{high},i} \cdot (-p), \tag{8}$$

where $\mathrm{MU}_{\mathrm{low},i} := [(1-\Delta c)Y]^{-\gamma_i}$ and $\mathrm{MU}_{\mathrm{high},i} := Y^{-\gamma_i}$ denote the marginal utilities of wealth in the low and high states, respectively. The consumer's **willingness to pay** for the contract, $u_i$, is defined as the price $p$ that sets this net gain to zero.

**Supply.** The **cost** to a firm of selling the contract is the average payout plus a fixed cost: $c_i := \pi_i + F$. We include the fixed cost as in Meza and Webb (2001). The fixed cost is both realistic and a parsimonious way to calibrate the model with less than 100% of consumers buying insurance in the case without selection. We could have instead assumed other frictions such as monopoly power or moral hazard.

**Equilibrium.** Given the joint distribution of the willingness to pay $u_i$ and cost $c_i$, we use the standard competitive selection model of Akerlof (1970) and Einav, Finkelstein, and Cullen (2010). Namely, Einav, Finkelstein, and Cullen (2010) define the **demand curve** as

$$D(p) := \int_{u_i > p} 1 \, di.$$

That is, the demand at a price $p$ is the share of consumers who are willing to pay at least $p$.

Einav, Finkelstein, and Cullen (2010) define the **average cost curve** as

$$AC(p) := \int_{u_i > p} c_i \, di / D(p).$$

The $AC$ curve is the inverse supply curve. $AC(q)$ is the average cost of selling insurance when the quantity of consumers covered is $q$. The defining feature of selection models is that the $AC$ curve depends on quantity, because due to selection the probability of loss depends on which consumers are buying insurance. Einav, Finkelstein, and Cullen (2010) define an **equilibrium** as a price and quantity pair $(p^*, q^*)$ in the intersection of the demand and supply curves.

#### 5.2 Willingness to pay

We can gain some intuition for the demand curve by solving for the willingness to pay. Setting Equation (8) to zero and substituting $u_i$ for $p$ yields

$$\frac{u_i}{1-u_i} = \frac{\pi_i}{1-\pi_i} \cdot \frac{\text{MU}_{\text{low},i}}{\text{MU}_{\text{high},i}}.$$

Under the CRRA assumption,

$$\frac{u_i}{1-u_i} = \frac{\pi_i}{1-\pi_i} \cdot (1-\Delta c)^{-\gamma_i}.$$

Or, solving for $u_i$,

$$u_i = \underbrace{\pi_i}_{\text{probability of loss}} \cdot \underbrace{\frac{(1-\Delta c)^{-\gamma_i}}{(1-\pi_i) + \pi_i(1-\Delta c)^{-\gamma_i}}}_{\text{risk premium}}.$$

That is, willingness to pay equals the probability of loss times the risk premium, with the latter being equal to the ratio of marginal utility in the loss state to average marginal utility. This clarifies that demand and equilibrium depend on the joint distribution of risk ($\pi_i$) and risk preferences ($\gamma_i$). Our analysis in the main text measures selection solely based on the distribution of risk, without considering risk preferences. We now proceed to calibrate the model, using the HRS data to inform the joint distribution of risk and risk preferences.

##### 5.2.1 Calibration

We calibrate the model to the HRS CAD contract from Section 4. We focus on consumers in the standard risk class. The probability of loss $\pi_i$ is the probability of having a heart problem as defined by the CAD HRS variable. We consider the same four information scenarios as in the main text for predicting $\pi_i$: the non-genetic covariates only (Scenario 1), the non-genetic covariates and the current PGI (Scenario 2), and the non-genetic covariates and the future PGI (lower and upper bounds; Scenarios 3L and 3U). All results are reported for the standard risk class, so this is a set of consumers that firms cannot price discriminate against.

To measure $\gamma_i$, we follow Kimball, Sahm, and Shapiro (2008). They developed methods to impute a relative risk aversion coefficient from the HRS questions. The estimated coefficient is based on questions about how respondents would choose between a job with certain earnings and a job with uncertain earnings. Their method yields, for each subject, an estimated coefficient of relative risk aversion $\gamma_i$. We note that the Kimball, Sahm, and Shapiro (2008) risk aversion coefficient estimates are relatively high, with the bulk of the distribution between $\gamma = 6$ and $\gamma = 10$. Naturally, the level of demand depends on both $\gamma$ and $\Delta c$. Thus, we can still accommodate a realistic overall level of demand, as long as we calibrate the model with a modest value of $\Delta c$. With these two pieces of data, we have a joint distribution of $\pi_i$ and $\gamma_i$.

This leaves two parameters to be calibrated: the fixed cost $F$ and the loss in income in the low state, $\Delta c$. We calibrate them to match the loss ratio and market size in the critical illness insurance market. We base the numbers on the UK, which is one of the countries with the most developed critical illness insurance markets. Swiss Re (2022) reports the number of policies sold in the UK in 2022. There were 484,110 critical illness policies sold as additional benefits to term life insurance policies. These correspond to the bulk of critical illness insurance sold, with an additional 94,426 standalone critical illness policies. The total number of term life insurance policies sold was 1,698,301. Given these figures, we perform the following back-of-the-envelope calculation. First, we assume that the potential market are the 1,698,301 consumers who purchase a term life insurance policy. This follows from the details of how these products are marketed, and that they are overwhelmingly sold as additional benefits to term life insurance buyers. If we consider only the 484,110 add-on critical illness policies sold, this implies a demand of 28.5% of the market. If we consider also the 94,426 standalone critical illness policies, this implies a demand of 34%. Thus, we set our calibration target to 30% of the market. Loss ratios in the industry are widely known to be high (the loss ratio equals claims paid divided by premiums). For critical illness, loss ratios in the ballpark of 50% are often cited, indicating a substantial amount of frictions (Reuters, 2020). We set 50% as our calibration target for the loss ratio.

The calibration matches the target loss ratio and market size exactly. The calibrated parameter values are a consumption loss $\Delta c$ of 12.5% and fixed cost $F$ of \$ 0.26 per dollar of coverage.

#### 5.3 Results

Figure 2 shows the willingness to pay for insurance as a function of disease risk and risk aversion. The distribution of relative risk aversion is the same in all scenarios. But the distribution of disease risk varies considerably depending on how much information consumers have. In Scenario 1, the distribution is concentrated around the average loss; this is as expected, since these consumers are in the standard risk class and only have access to information on their non-genetic covariates, which is what insurers use to define the standard risk class. In Scenario 2, with the non-genetic covariates and the current PGI, the distribution becomes more dispersed since consumers have access to more information. Scenarios 3L and 3U display a wide dispersion in risks, reflecting our main findings that future PGIs will be highly predictive.

Willingness to pay varies predictably with risk aversion and disease risk. The least risk averse consumers are willing to pay slightly more than their expected loss—e.g., a low-risk-aversion consumer with disease risk of 50% is willing to pay slightly more than 50 cents on the dollar of coverage. The most risk averse consumers are willing to pay larger premiums. While both disease risk and risk aversion affect the willingness to pay,

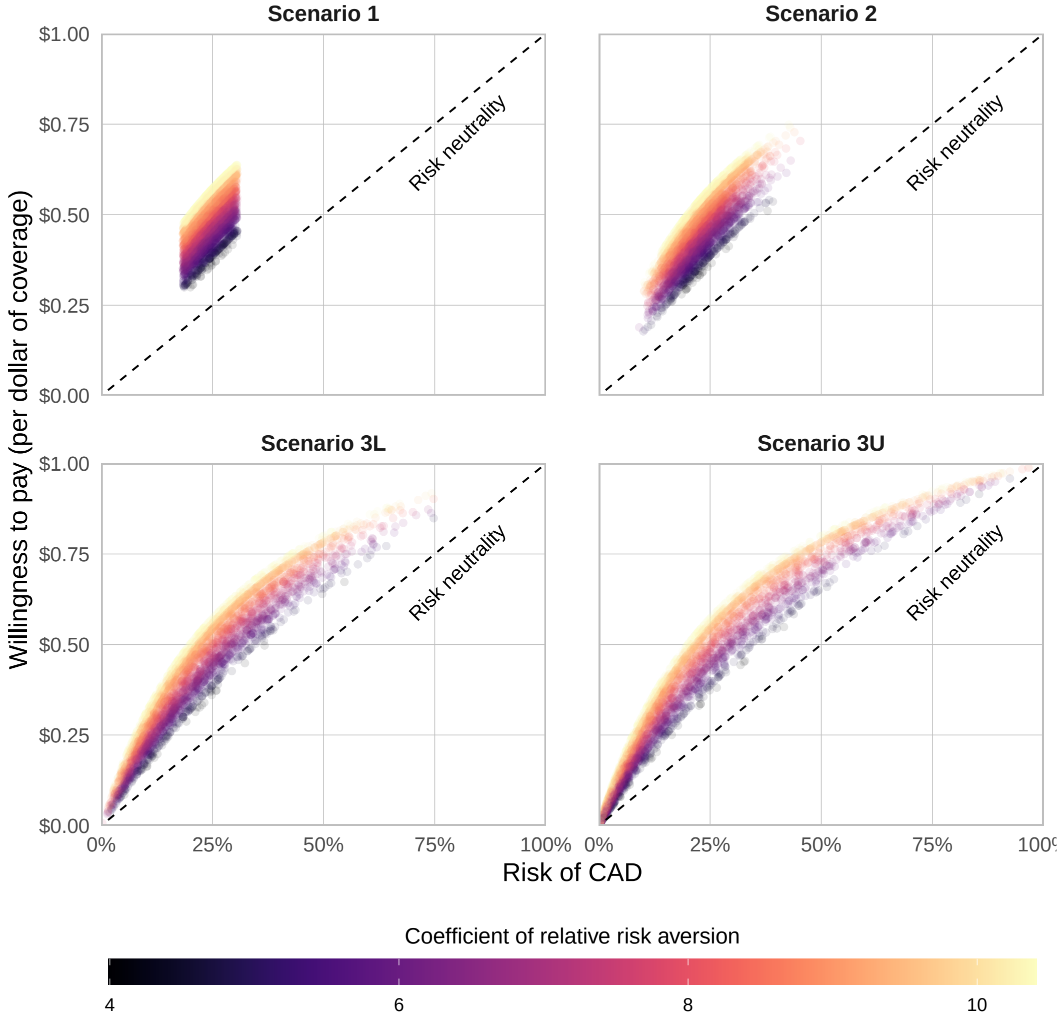


Figure 2: Willingness to pay for insurance as a function of disease risk and risk aversion

*Notes:* Willingness to pay is calculated in the calibrated equilibrium model for the HRS CAD contract using the calibrated consumption loss $\Delta c$ of 12.5%. Each point represents an individual, with the color indicating the individual's relative risk aversion. The four scenarios are defined in the text.

the impact of disease risk is much more substantial.

We now turn to equilibrium. Figure 4 in the main text plots supply, demand, and equilibria under our different information scenarios. In Scenario 1, consumers only have information from the non-genetic covariates. There is nearly no selection almost by definition, since we are considering the standard risk class, which insurers define using the non-genetic covariates. The average cost curve is accordingly nearly flat. This means that the average riskiness of buyers is about the same, regardless of whether a small or large fraction of the population buys insurance.

There is a unique equilibrium in Scenario 1. Quantity is positive, matching almost exactly our calibration target of 30% of the market buying insurance. This also follows from our assumptions, since we calibrated the fixed cost $F$ and consumption loss $\Delta c$ to match the market size and loss ratio.

In Scenario 2, the current PGI is added to the information set. The demand and average cost curves noticeably rotate, becoming steeper. Consumers now have a bit more information. This both creates more variation in willingness to pay, rotating demand, and also makes higher demand more correlated with higher risk. This creates some selection, making the AC curve noticeably steeper. The equilibrium quantity goes down to 21.4%.

Scenarios 3L and 3U use the future PGI based on the heritability assumptions. As previously noted, the future PGI greatly improves predictive power. Accordingly, the demand and average cost curves become much steeper. In both cases, we find that the only equilibrium is the zero quantity. That is, we have a complete death spiral, with no insurance sold.

The findings of the calibrated equilibrium model for the HRS CAD contract are consistent with the qualitative findings from our main UKB analysis in the main text. The UKB analysis considers only the distribution of risk, ignoring risk preferences. That analysis finds that selection is noticeable with the current PGI and would be high and potentially crippling with the future PGI. The results in the calibrated equilibrium model are consistent with this view. The calibrated model takes into account the correlation between risk and risk preferences in the HRS, and is estimated for the least selected of all the contracts we analyze, suggesting that the results of the UKB analysis are also robust to risk preferences and equilibrium considerations (as we further discuss in main text Section 6.3).

We caveat that this conclusion may not hold in different equilibrium models. For example, one could consider a model that assumes that consumers have very high willingness to pay for insurance and behavioral demand for insurance that does not depend on risk. In that model, we could have 100% of consumers buying insurance regardless of the predictability of risk. The goal of this section is simply to consider one parsimonious data-driven calibration and examine the robustness of our main findings, but it would be interesting to consider further robustness checks.

### 6 Overview of policy responses

To complement the discussion in the main text, we now give a brief overview of the literature on the regulation of selection markets. We also discuss how the ideas relate to genetic private information. As mentioned in the main text, regulation of selection markets typically seeks to balance the goals of efficiency and redistribution. The former aims to maximize total economic surplus, while the latter tries to help particular groups. Redistribution includes solidarity towards consumers with lower wealth, worse health status, or genes that are predictive of serious illnesses. The standard regulatory playbook contains three main approaches.

The first approach is **laissez-faire regulation**. That is, allowing firms to set prices as they wish, with minimal regulation focused on the financial solvency of insurers. The standard example is life insurance. Life insurance costs have a large predictable component because age and health status are highly predictive of mortality. In most countries, insurers are free to use these variables when underwriting policies. The laissez-faire approach minimizes adverse selection. In life insurance, it is thought that there is relatively little adverse selection, and that insurers often predict mortality better than consumers (Gottlieb and Smetters, 2021). However, the laissez-faire approach ignores redistribution. The elderly and other consumers with high mortality pay higher premiums. This is deemed acceptable in life insurance because premiums are a small part of most consumers' income, and redistribution is not a first-order issue.

The second, and opposite, approach is **government provision**. In many developed countries, basic health insurance is publicly provided. One reason is that redistributive issues are central in health insurance. Insurance costs are usually considerable relative to personal income because healthcare represents a substantial share of GDP (18% in the United States). In most countries, there is widespread agreement that the destitutes should be granted some level of care, and that high-cost consumers such as the elderly should be subsidized (Einav, Finkelstein, and Fisman, 2023). For these reasons, basic health insurance coverage is publicly provided in most developed countries. Public provision of any kind of insurance has three main potential drawbacks. First, the public sector may be less efficient. Second, innovation can be curbed, especially for innovative products like critical illness insurance. Third, the quality and quantity of public coverage might be inefficiently too high or too low.

The third approach is **managed competition**. Managed competition combines private provision with regulations to improve market performance. In the United States, examples include individual health insurance exchanges and insurance options sponsored by large employers. In most other developed countries, the main example is complementary health insurance. Managed competition involves four main kinds of regulations: community rating, mandates, subsidies, and risk adjustment. Community rating imposes restrictions on what variables insurers can use to set prices. Community rating is used for redistribution reasons (such as lowering premiums for the elderly) but has the cost of increasing adverse selection (Handel, Hendel, and Whinston, 2015). Mandates impose penalties on consumers who do not purchase insurance, and subsidies lower the prices of policies; these are important for increasing the number of consumers who purchase policies, which otherwise tends to be inefficiently too low due to adverse selection (Geruso et al., 2021; Chade et al., 2022; Veiga and Levy, 2022). Risk adjustment is a type of subsidy to insurers who cover sicker patients. Risk adjustments and other subsidies are thought to be important to guarantee that firms don't offer contracts that provide inefficiently low coverage (Glazer and McGuire, 2000).

Consider how these standard approaches would work in the case of genetic information and CII. The laissez-faire approach would be to allow companies to underwrite based on genetic information. It is likely that—at least at some point in the future when genetic prediction technology has further improved—companies would require genetic testing, much like they require medical exams today. This would lead to little adverse selection. The downside of this policy is that critical illness insurance would be very expensive for consumers with high risk. For example, consider a consumer with a 25% risk of developing prostate cancer. This consumer would have to pay $25,000 in lifetime premiums for a $100,000 CII contract for prostate cancer. Our results suggest that, with improved prediction technology, many consumers would be in this situation, which might create pushback against the laissez-faire approach.

The public provision approach would be for governments to publicly provide CII for all consumers. This strikes us as being unlikely to be popular. Most people do not hold CII, and providing it to a broad segment of the population is unlikely to be a priority for the government.

The managed competition approach includes a broad menu of options. In fact, the current policy (in most developed countries) of genetic information bans falls within managed competition. A genetic information ban is simply a community rating regulation. Our results suggest it is likely that this type of community rating will lead to high levels of selection in the future. In that case, industry participants and regulators may want to address selection with additional policies like risk adjustment and subsidies. The design of efficient policies would depend on empirical work, along the lines of existing research on current selection markets.

### 7 Acknowledgments

This research was reviewed and approved by the Committee Ethics and Data of the Leiden Law School of Leiden University (application number 2023-24). The use of the UK Biobank data for this research has also been reviewed by the George Mason University Institutional Review Board (IRB) Office, which has determined it to be exempt from IRB review. The use the UK Biobank data was also reviewed by and granted exemptions by the University of Southern California University Park Institutional Review Board and the Vrije Universiteit Amsterdam Research Ethics Committee. This research has been conducted using the UK Biobank Resource under application numbers 217716, 99086, and 11425. This publication is part of the project "The Economic Consequences of New Genetic Testing Technologies" (project number Vl.Veni.221E.080) of the NWO Talent Programme Veni – Social Sciences and Humanities, which is (partly) financed by the Dutch Research Council (NWO). This work used the Dutch national e-infrastructure with the support of the SURF Cooperative using grant no. EINF-8891. This research was also supported by a Faculty Research and Development Award (FRDA) at George Mason University and by the National Science Foundation (NSF award nos. 2343735 and 2343736).

#### 7.1 Additional acknowledgments and GWAS data availability

We thank the participants and investigators of the FinnGen study for providing GWAS summary statistics for these analyses (Kurki et al., 2023). The GWAS summary statistics for Alzheimer's disease and colorectal cancer derived in the FinnGen biobank can be accessed here: https://www.finngen.fi/en/access_results. We thank the International Genomics of Alzheimer's Project (IGAP) for providing summary statistics for these analyses (J.-C. Lambert et al., 2013). The GWAS summary statistics for Alzheimer's disease and an extended acknowledgment of the relevant funding sources can be accessed here: https://www.niagads.org/datasets/ng00036. We thank the participants and investigators of the Breast Cancer Association Consortium for providing the summary statistics for these analyses (Michailidou et al., 2017; Zhang et al., 2020). The GWAS summary statistics for breast cancer and an extended acknowledgment of the relevant funding sources can be accessed here: https://bcac.ccge.medschl.cam.ac.uk/bcacdata/. We thank the participants and investigators of the CARDIoGRAMplusC4D Consortium (Nikpay et al., 2015). The GWAS summary statistics for coronary artery disease can be accessed here: http://www.cardiogramplusc4d.org/. We thank the participants and investigators that contributed to producing the following GWAS summary statistics that were downloaded from the NHGRI-EBI GWAS Catalog (Sollis et al., 2022): Colorectal cancer study ID(s) GCST012876, GCST012877, GCST012878, GCST012879, GCST012880 (Huyghe et al., 2019; Fernandez-Rozadilla et al., 2023); Prostate cancer study ID(s) GCST006085 (Schumacher et al., 2018). The GWAS summary statistics on colorectal cancer and prostate cancer downloaded from the NHGRI-EBI GWAS Catalog can be accessed here: https://www.ebi.ac.uk/gwas/downloads/summary-statistics. We thank the participants and investigators of the Psychiatric Genomics Consortium (Trubetskoy et al., 2022). The GWAS summary statistics for schizophrenia can be accessed here: https://pgc.unc.edu/for-researchers/download-results/. We thank the participants and investigators of the DIAGRAM Consortium (Mahajan et al., 2018). The GWAS summary statistics for type 2 diabetes can be accessed here: http://diagram-consortium.org/downloads.html.
