## Supplementary Figures for "Genetic prediction and adverse selection"

First version: October, 2024
This version: May 28, 2025

*The Wharton School, University of Pennsylvania,
†Interdisciplinary Center for Economic Science and Department of Economics, George Mason University,
‡Department of Economics, Leiden University,
School of Business and Economics, Vrije Universiteit Amsterdam,
Supplementary Information files are available at: `https://osf.io/9ndw6/files/osfstorage`

### 1 Other supporting figures

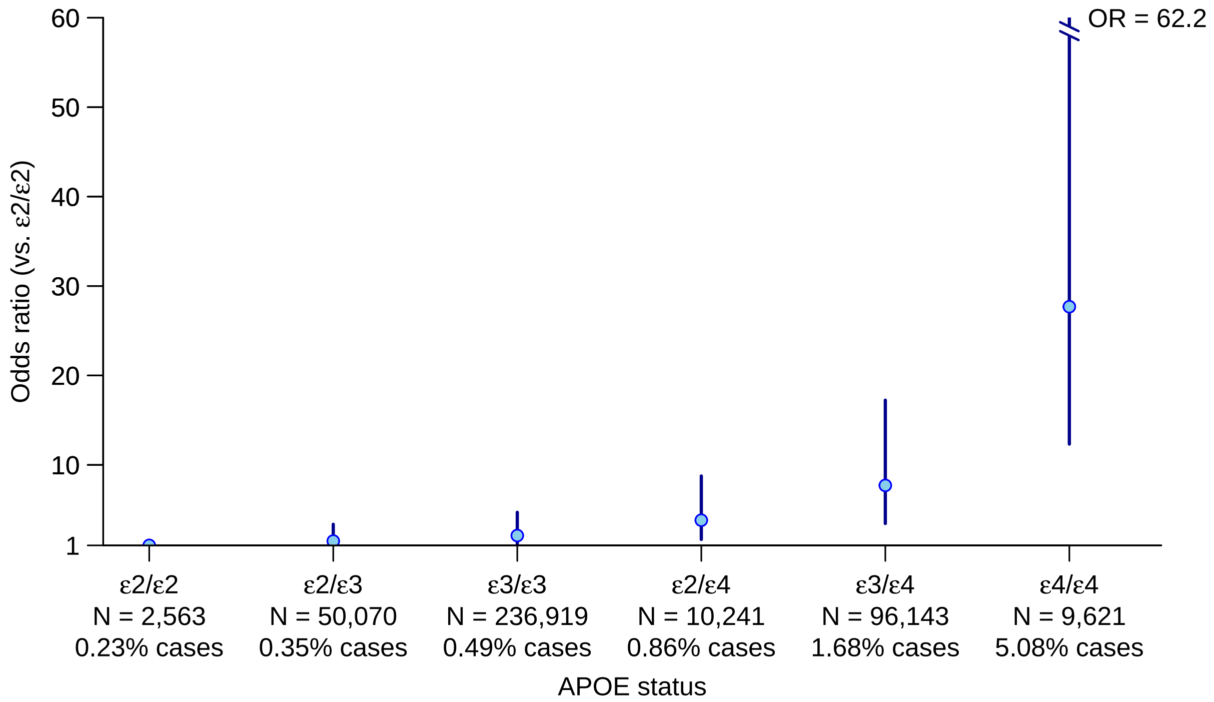


Figure 1: The risk of Alzheimer's disease by *APOE* risk type

*Notes*: The dots report (on the y-axis) the odds ratio of contracting Alzheimer's for each *APOE* risk type, and the error bars represent 95% confidence intervals (for the odds ratio). The odds ratios were estimated among 405,573 UK Biobank participants with logistic regression controlling for age, sex, and the first ten genetic PCs. "N = " and "% cases" report, respectively, the number of individuals and the percentage with Alzheimer's disease for each risk type. These percentages are lower than the corresponding population lifetime risks because the average age in our UKB sample is 65 years old and Alzheimer's is more common after that age. The *APOE* status dummy variables explain $\sim$5.4% of the variance on the latent disease scale.

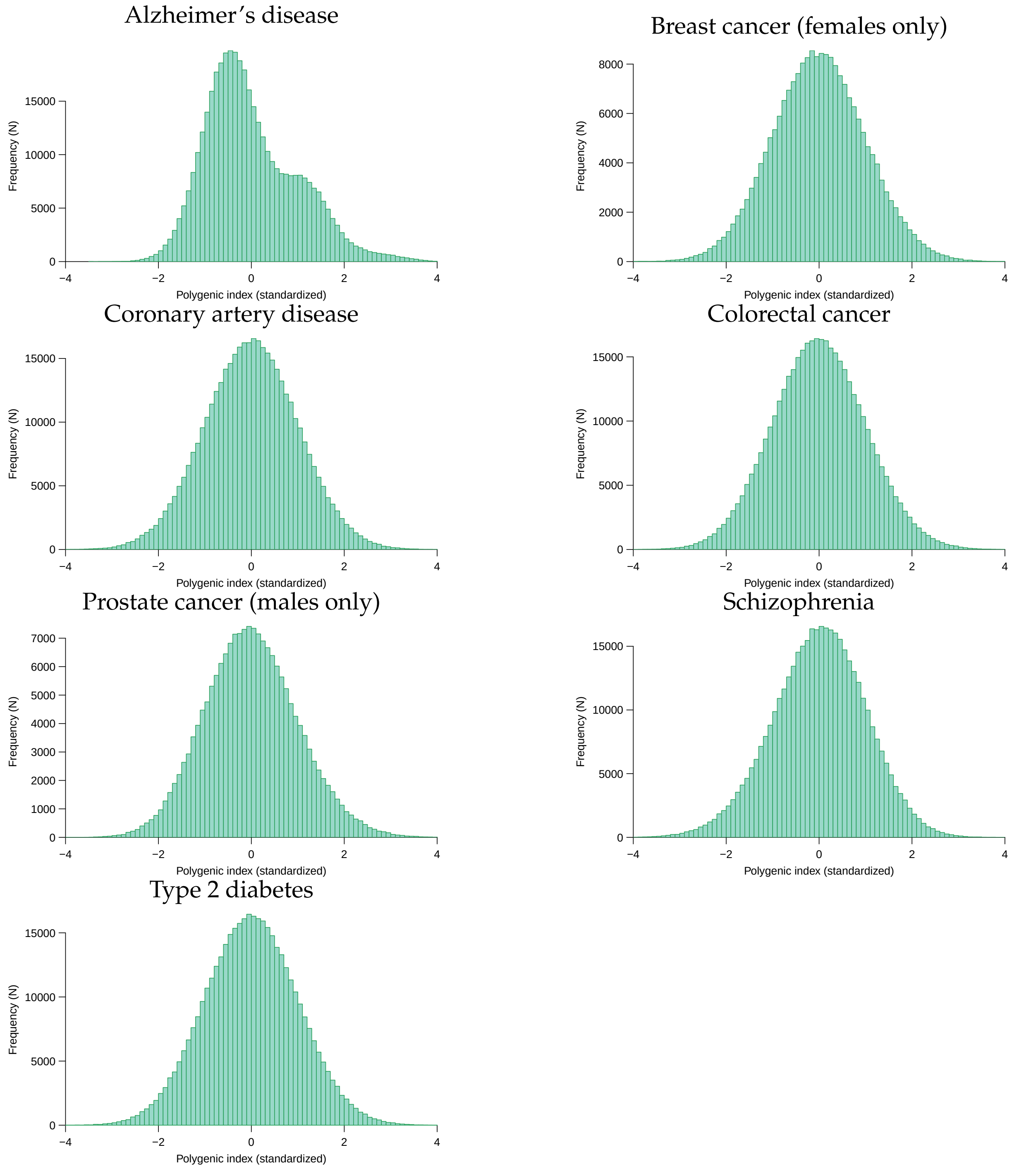


Figure 2: Histograms of the PGI distributions

*Notes*: The histograms display the distributions of the PGIs for our seven diseases of interest in the UKB. The PGIs were standardized to have mean zero and unit variance separately by sex. The PGI for Alzheimers disease has three modes because it captures the *APOE* risk types, as discussed in Supplemental Appendix 1.5.

### 2 Supplementary results for the standard risk class

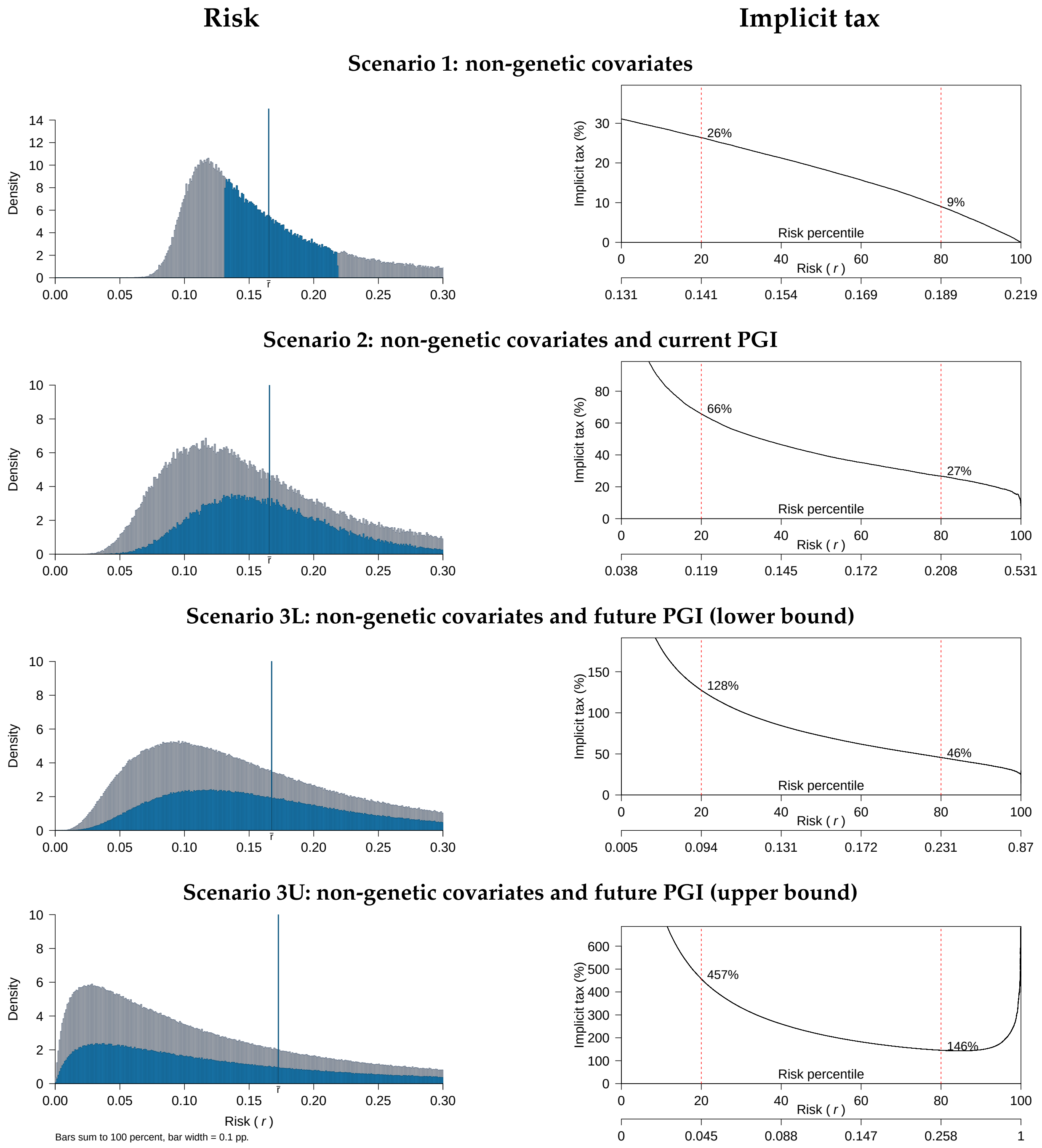


Figure 3: Multiple-disease contract for females: risk and implicit tax

*Notes*: Left panel: distribution of the risk of contracting any of the six diseases in the female multiple-disease contract by age 65, conditional on various information scenarios and computed in the UKB data ($N = 204,672$). The vertical blue line marks the average risk in the standard risk class ($\bar{r} = 0.1652$). The standard risk class individuals are shown in blue; insurers treat these individuals identically, so the blue distribution corresponds to the distribution of private risk for these individuals. Right panel: implicit tax for consumers in the standard risk class as a function of their percentile private risk of any of the six diseases in the female multiple-disease contract for each scenario.

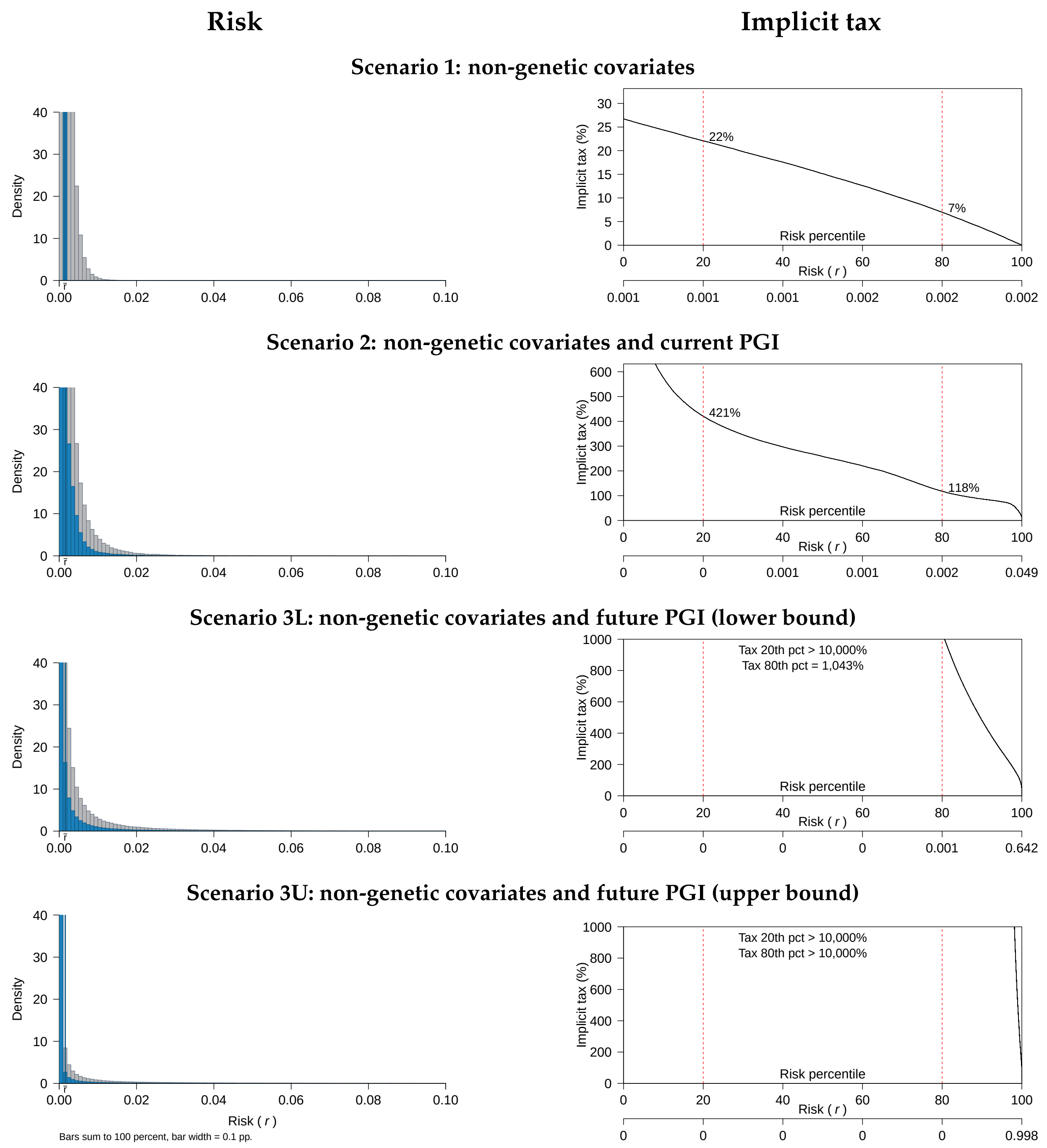


Figure 4: Single-disease CII contract for Alzheimer's disease: risk and implicit tax

*Notes*: Left panel: distribution of the risk of contracting Alzheimer's disease by age 65, conditional on various information scenarios and computed in the UKB data ($N = 405,573$). The vertical blue line marks the average risk in the standard risk class ($\bar{r} = 0.0015$). The standard risk class individuals are shown in blue; insurers treat these individuals identically, so the blue distribution corresponds to the distribution of private risk for these individuals. Right panel: implicit tax for consumers in the standard risk class as a function of their percentile private risk of Alzheimer's disease for each scenario.

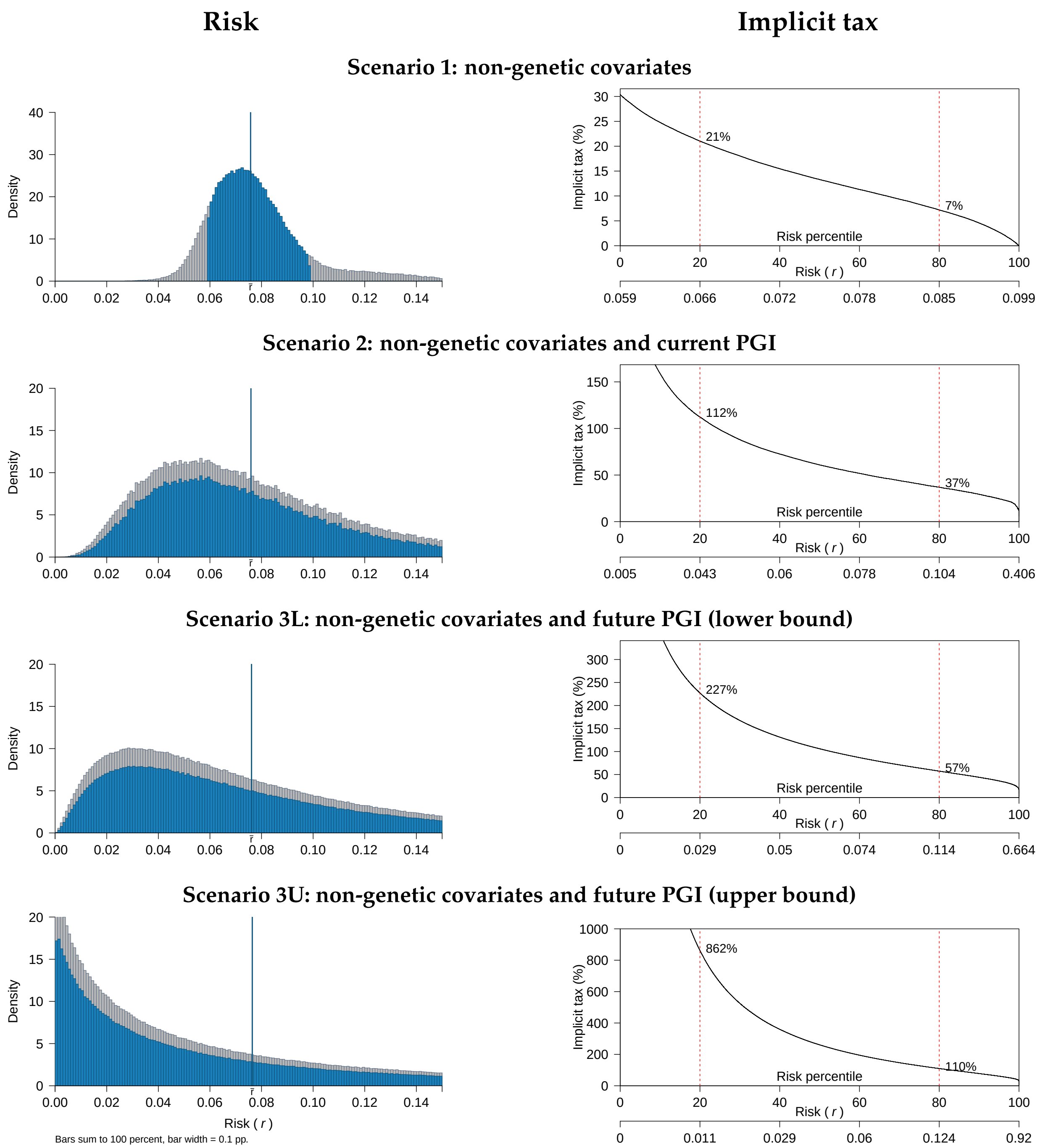


Figure 5: Single-disease CII contract for breast cancer: risk and implicit tax

*Notes*: Left panel: distribution of the risk of contracting breast cancer by age 65, conditional on various information scenarios and computed in the UKB data ($N = 211,575$). The vertical blue line marks the average risk in the standard risk class ($\bar{r} = 0.0758$). The standard risk class individuals are shown in blue; insurers treat these individuals identically, so the blue distribution corresponds to the distribution of private risk for these individuals. Right panel: implicit tax for consumers in the standard risk class as a function of their percentile private risk of breast cancer for each scenario.

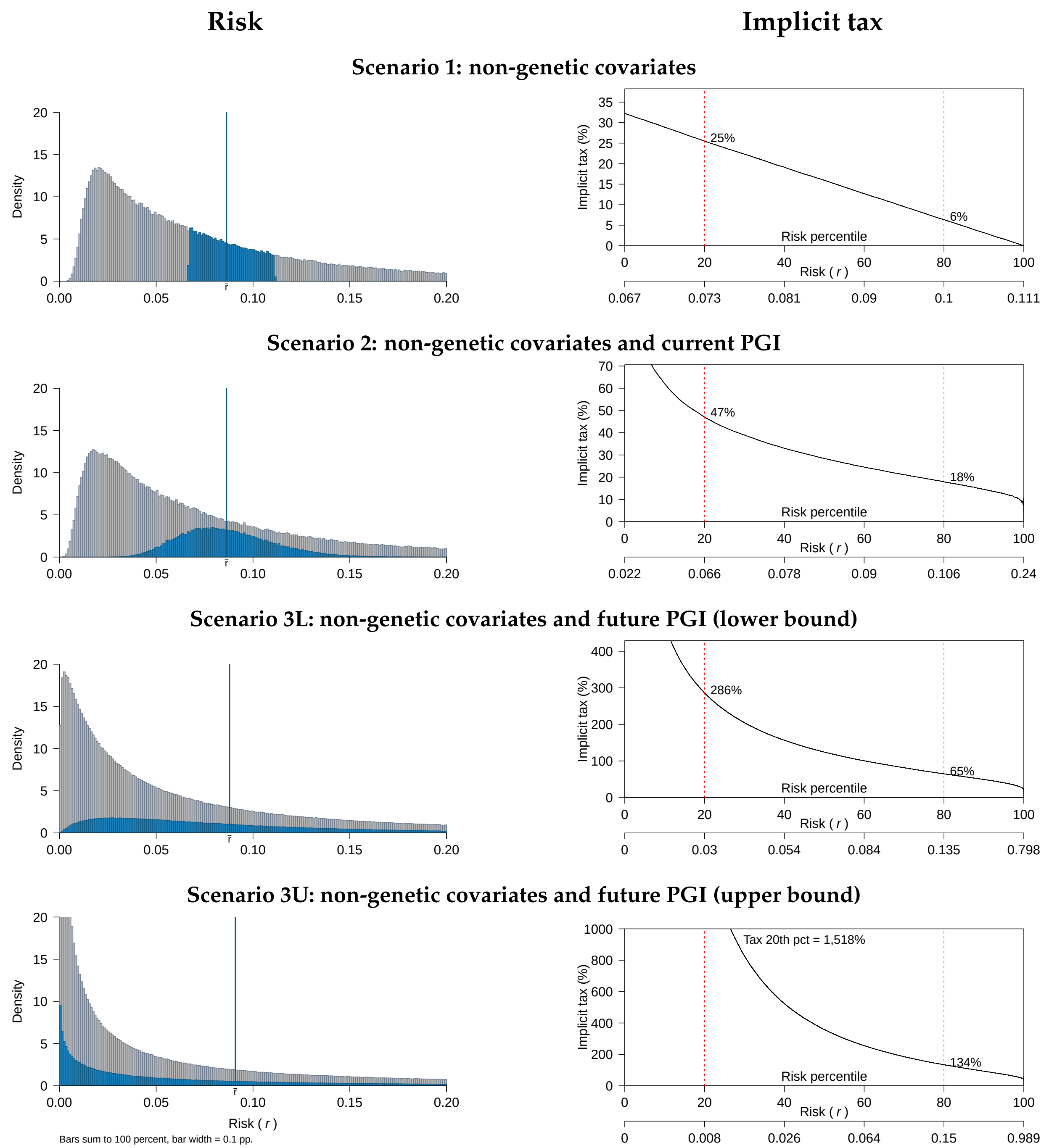


Figure 6: Single-disease CII contract for coronary artery disease: risk and implicit tax

*Notes*: Left panel: distribution of the risk of contracting coronary artery disease by age 65, conditional on various information scenarios and computed in the UKB data ($N = 410,686$). The vertical blue line marks the average risk in the standard risk class ($\bar{r} = 0.0863$). The standard risk class individuals are shown in blue; insurers treat these individuals identically, so the blue distribution corresponds to the distribution of private risk for these individuals. Right panel: implicit tax for consumers in the standard risk class as a function of their percentile private risk of coronary artery disease for each scenario.

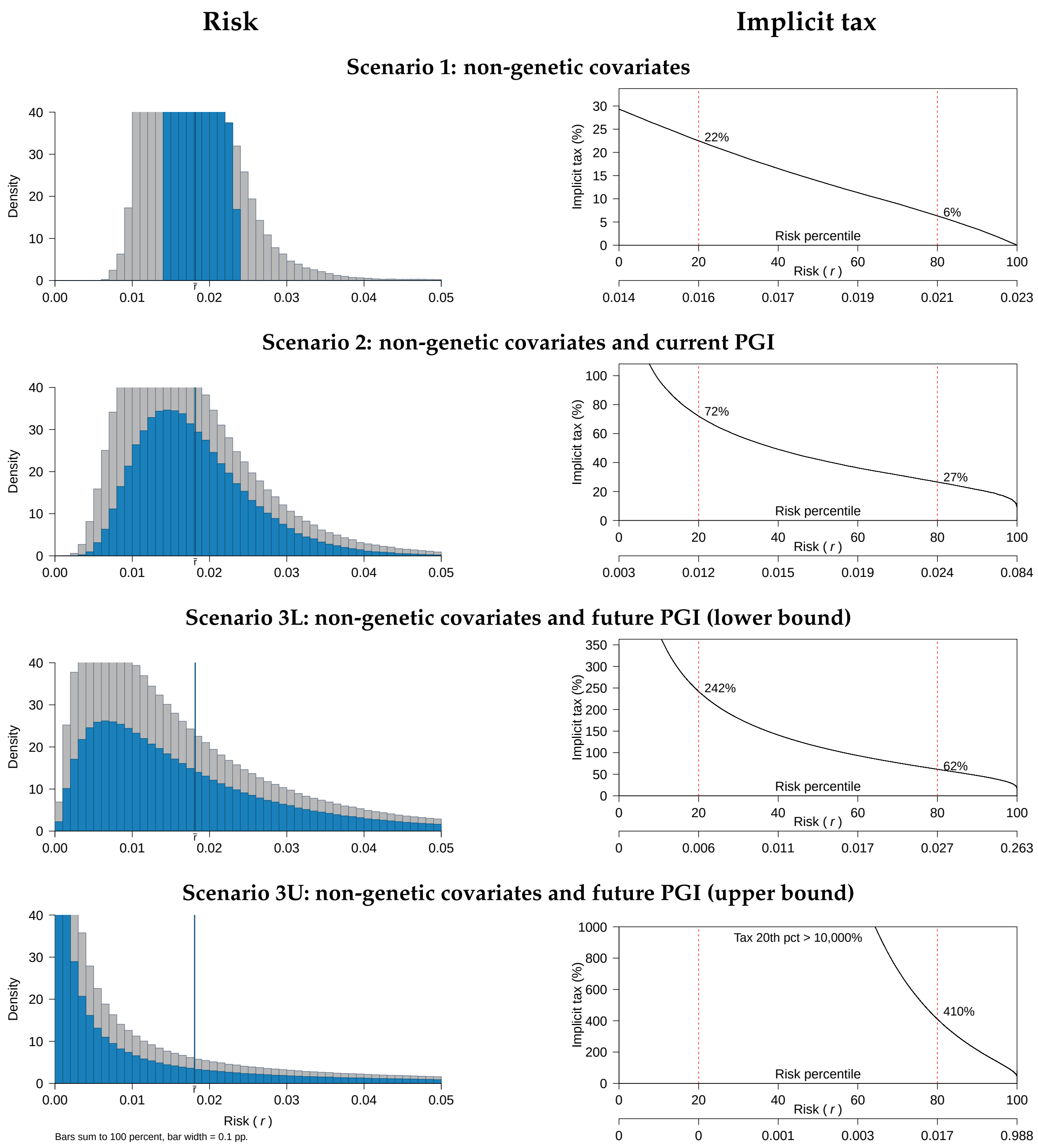


Figure 7: Single-disease CII contract for colorectal cancer: risk and implicit tax

*Notes*: Left panel: distribution of the risk of contracting colorectal cancer by age 65, conditional on various information scenarios and computed in the UKB data ($N = 410,562$). The vertical blue line marks the average risk in the standard risk class ($\bar{r} = 0.0182$). The standard risk class individuals are shown in blue; insurers treat these individuals identically, so the blue distribution corresponds to the distribution of private risk for these individuals. Right panel: implicit tax for consumers in the standard risk class as a function of their percentile private risk of colorectal cancer for each scenario.

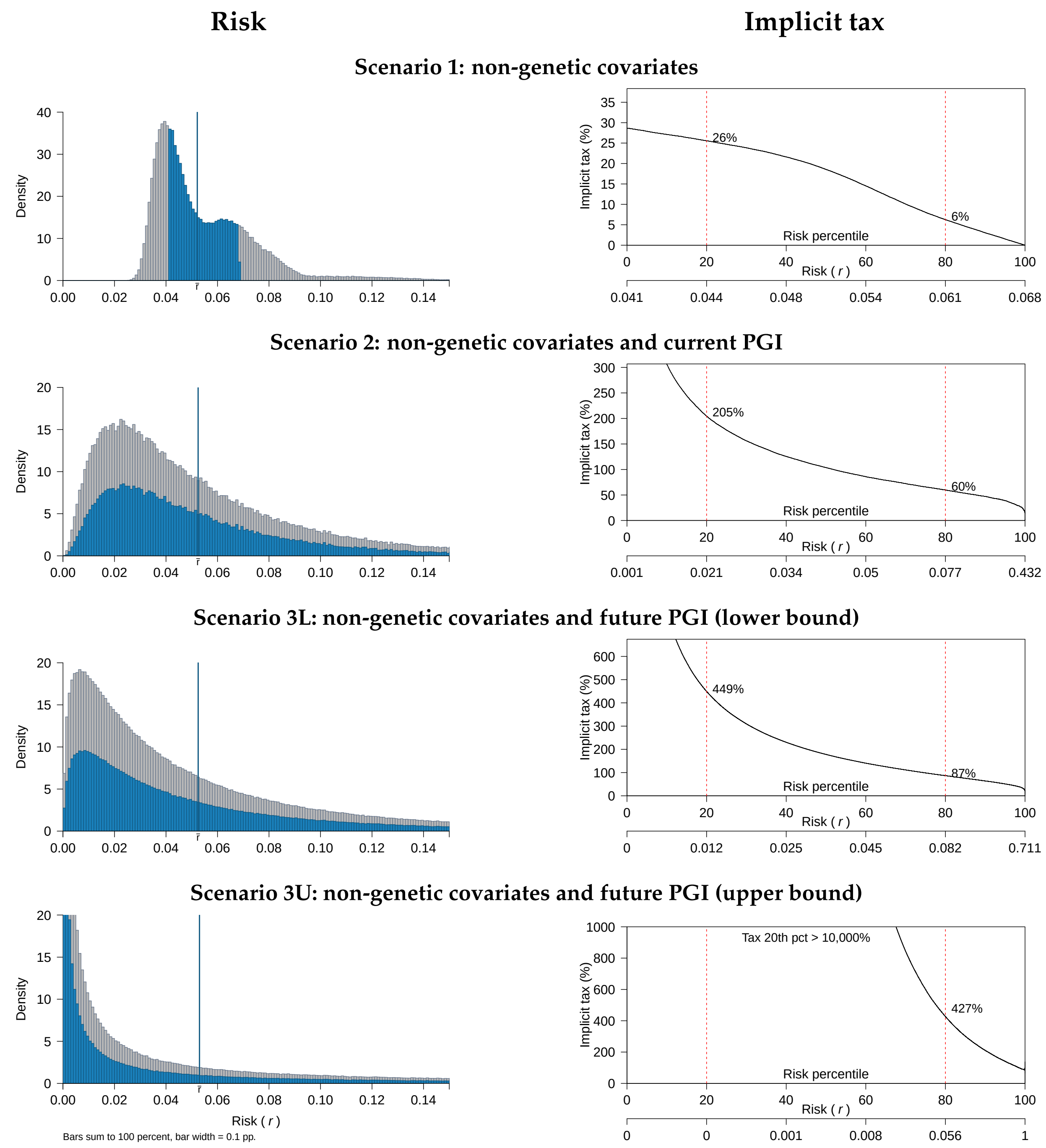


Figure 8: Single-disease CII contract for prostate: risk and implicit tax

*Notes*: Left panel: distribution of the risk of contracting prostate cancer by age 65, conditional on various information scenarios and computed in the UKB data ($N = 181,902$). The vertical blue line marks the average risk in the standard risk class ($\bar{r} = 0.0521$). The standard risk class individuals are shown in blue; insurers treat these individuals identically, so the blue distribution corresponds to the distribution of private risk for these individuals. Right panel: implicit tax for consumers in the standard risk class as a function of their percentile private risk of prostate cancer for each scenario.

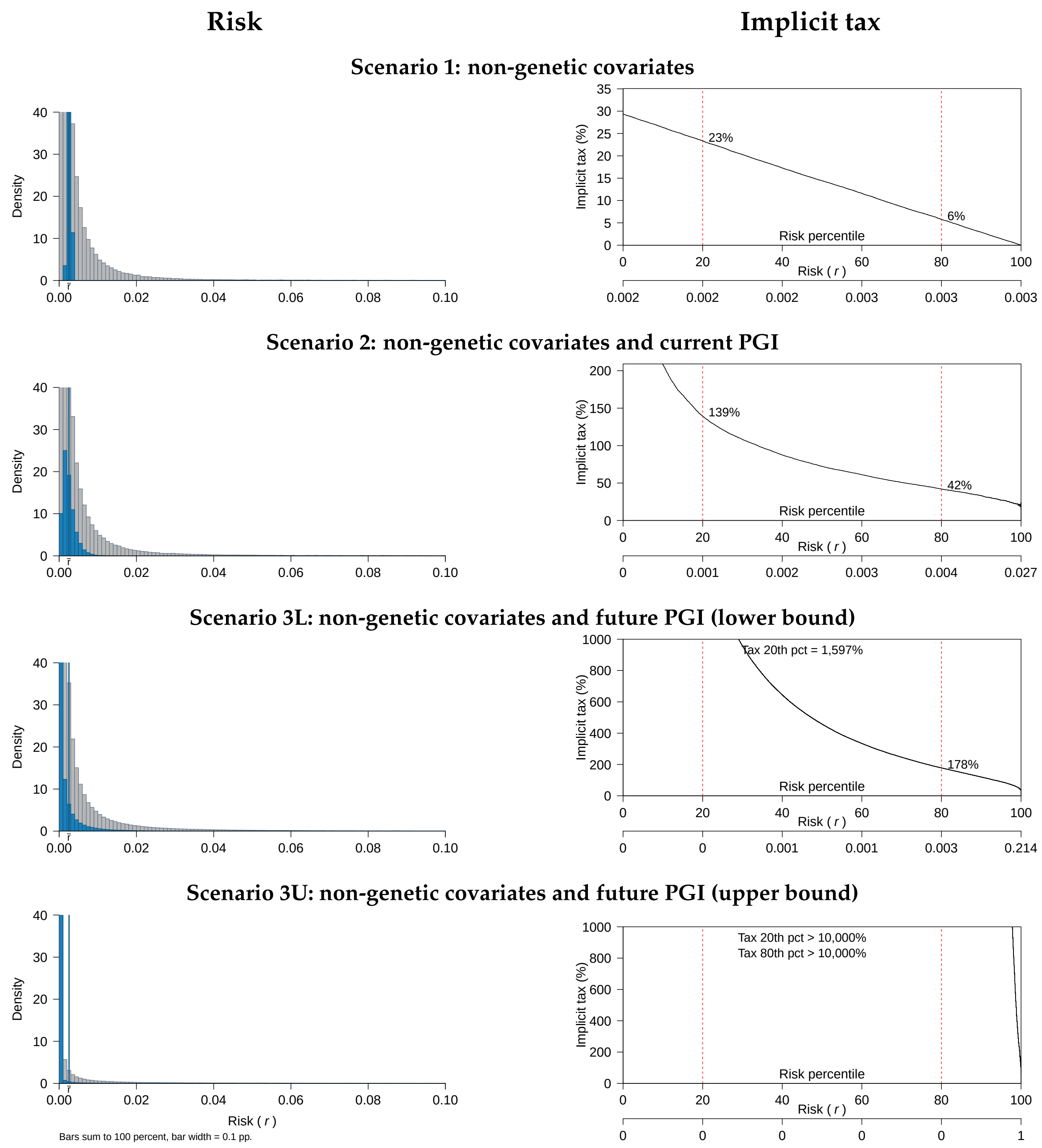


Figure 9: Single-disease CII contract for schizophrenia: risk and implicit tax

*Notes*: Left panel: distribution of the risk of contracting schizophrenia by age 65, conditional on various information scenarios and computed in the UKB data ($N = 410,275$). The vertical blue line marks the average risk in the standard risk class ($\bar{r} = 0.0025$). The standard risk class individuals are shown in blue; insurers treat these individuals identically, so the blue distribution corresponds to the distribution of private risk for these individuals. Right panel: implicit tax for consumers in the standard risk class as a function of their percentile private risk of schizophrenia for each scenario.

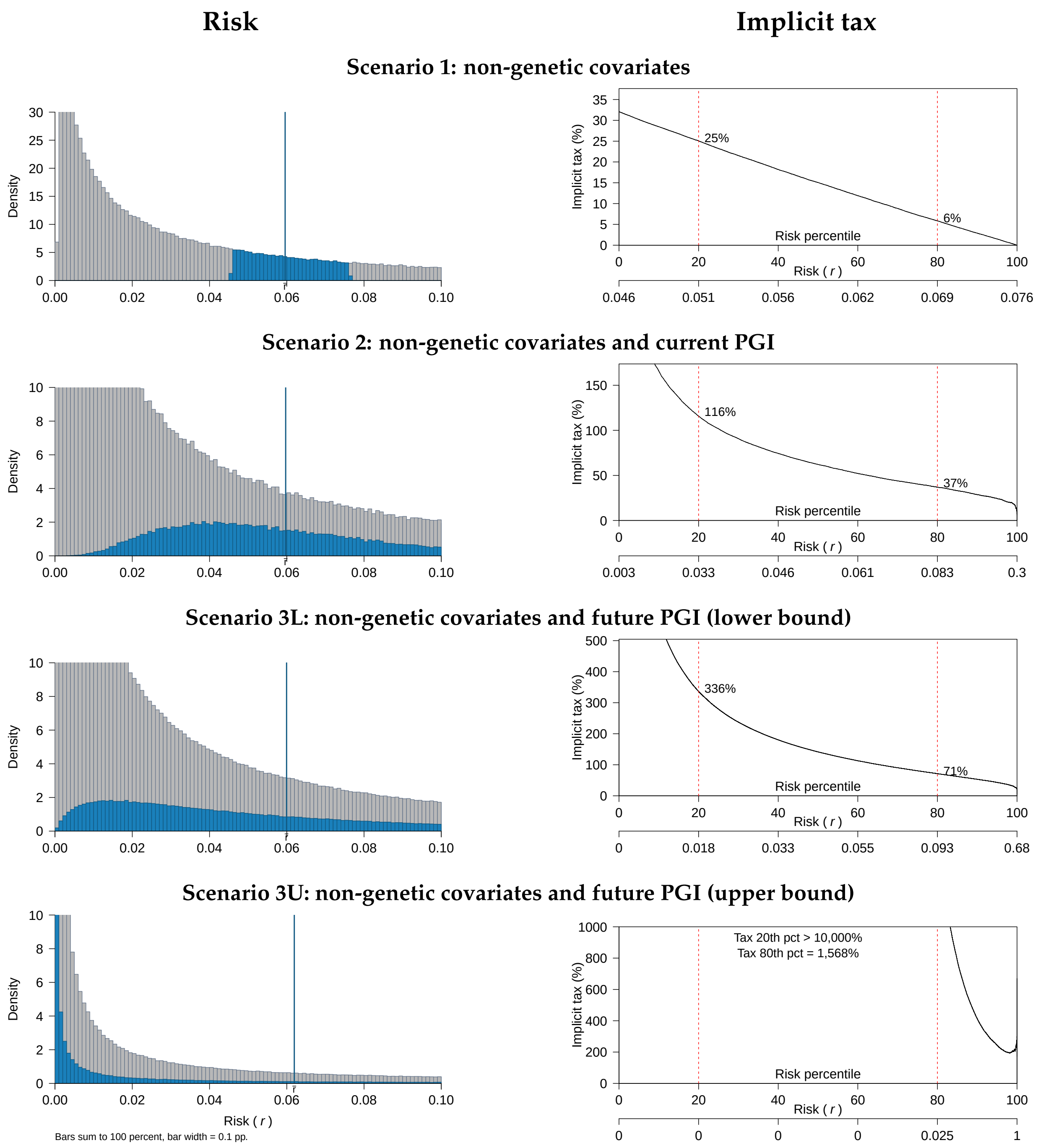


Figure 10: Single-disease CII contract for type 2 diabetes: risk and implicit tax

*Notes*: Left panel: distribution of the risk of contracting type 2 diabetes by age 65, conditional on various information scenarios and computed in the UKB data ($N = 410,686$). The vertical blue line marks the average risk in the standard risk class ($\bar{r} = 0.0596$). The standard risk class individuals are shown in blue; insurers treat these individuals identically, so the blue distribution corresponds to the distribution of private risk for these individuals. Right panel: implicit tax for consumers in the standard risk class as a function of their percentile private risk of type 2 diabetes for each scenario.

### 3 Selected results for the standard risk class under the assumption of partial take-up of genetic testing

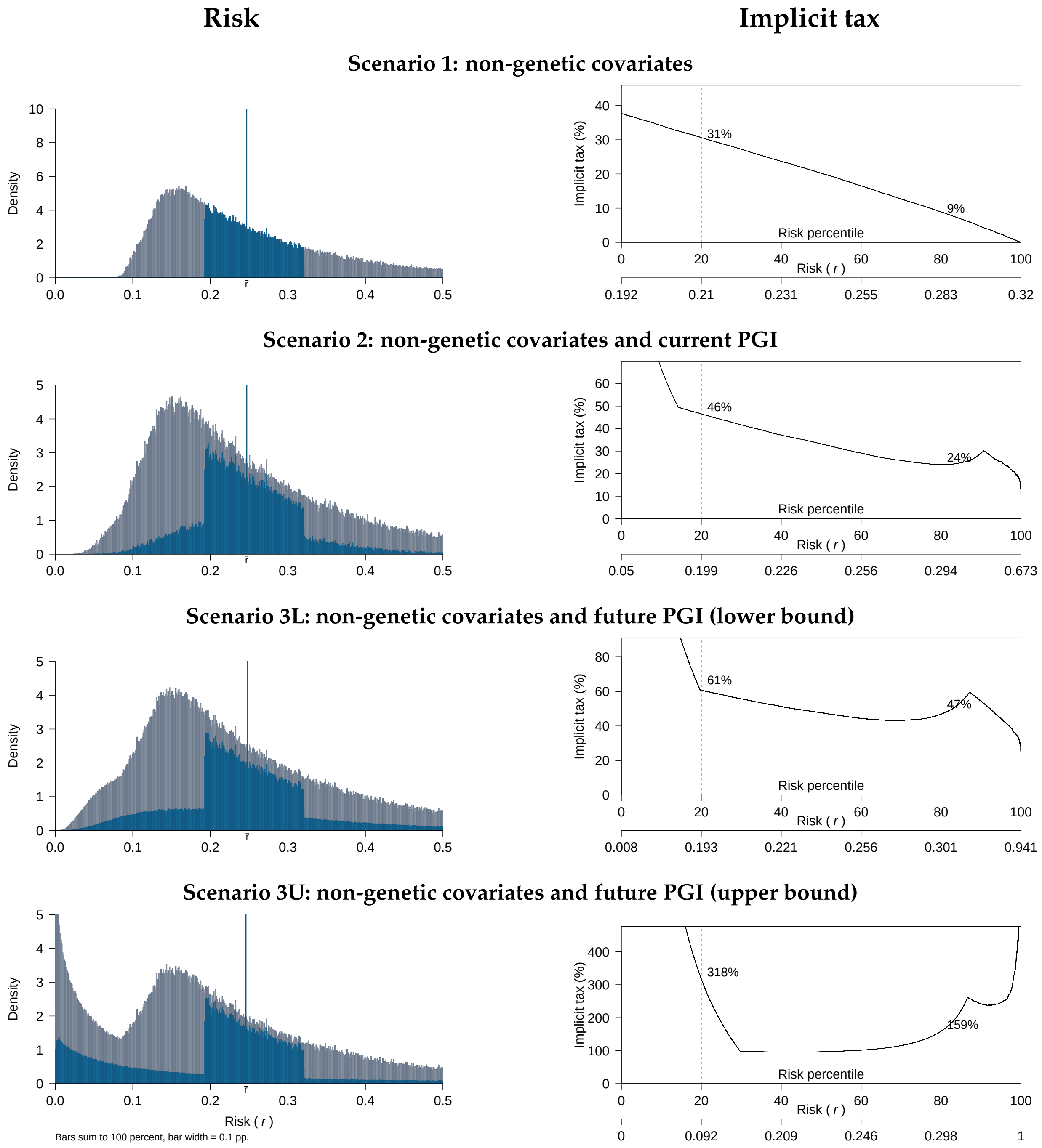


Figure 11: Multiple-disease contract for males: risk and implicit tax

*Notes*: Left panel: distribution of the risk of contracting any of the six diseases in the male multiple-disease contract by age 65, conditional on various information scenarios and computed in the UKB data ($N = 175,466$). The vertical blue line marks the average risk in the standard risk class ($\bar{r} = 0.2467$). The standard risk class individuals are shown in blue; insurers treat these individuals identically, so the blue distribution corresponds to the distribution of private risk for these individuals. Right panel: implicit tax for consumers in the standard risk class as a function of their percentile private risk of any of the six diseases in the male multiple-disease contract for each scenario.

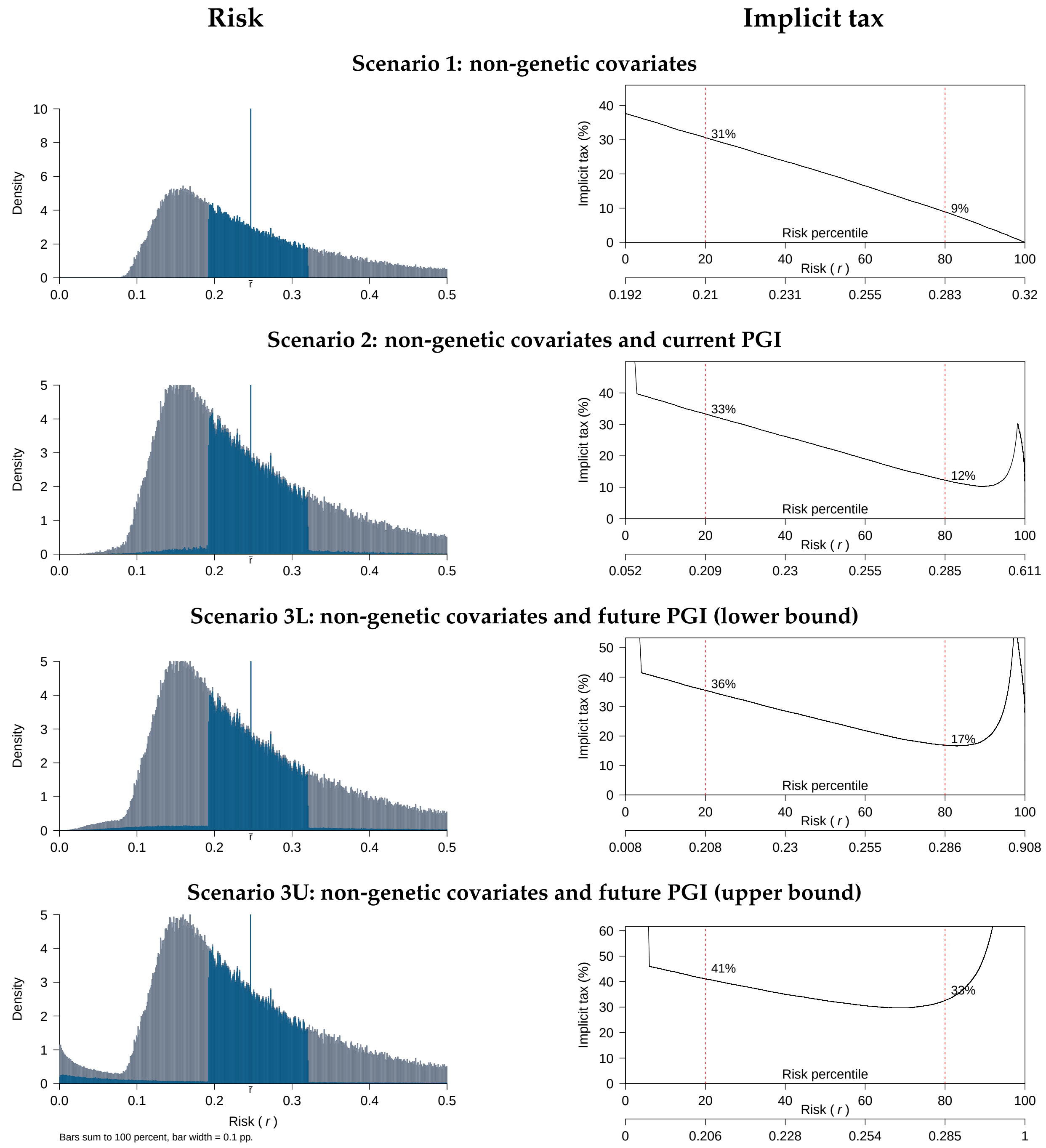


Figure 12: Multiple-disease contract for males: risk and implicit tax

*Notes*: Left panel: distribution of the risk of contracting any of the six diseases in the male multiple-disease contract by age 65, conditional on various information scenarios and computed in the UKB data ($N = 175,466$). The vertical blue line marks the average risk in the standard risk class ($\bar{r} = 0.2467$). The standard risk class individuals are shown in blue; insurers treat these individuals identically, so the blue distribution corresponds to the distribution of private risk for these individuals. Right panel: implicit tax for consumers in the standard risk class as a function of their percentile private risk of any of the six diseases in the male multiple-disease contract for each scenario.

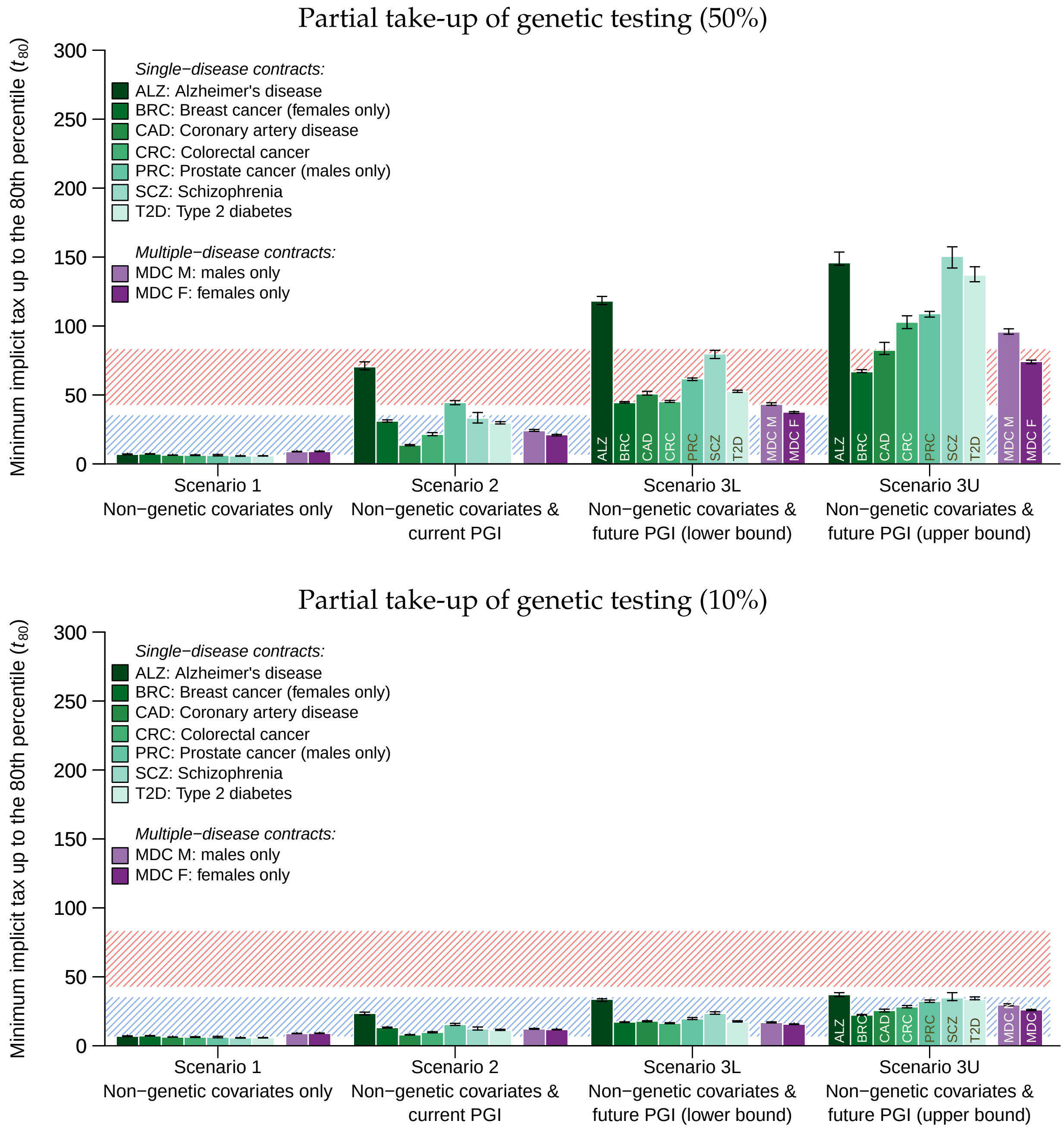


Figure 13: Minimum implicit taxes up to the 80th percentile ($t_{80}$) assuming partial take-up
*Notes*: The figure displays $t_{80}$ within the standard risk class for the single-disease and multi-disease CII contracts, in the four scenarios. The top and bottom panels show results assuming only 50% and 10% of consumers perform genetic tests. The striped blue area corresponds to the range of $t_{80}$ observed by Hendren (2013) in market segments that had not unraveled, and the striped red area corresponds to the range for market segments that had unraveled. The results in this figure are also reported in Supplementary Table 7.
